## Supplementary figures and images for "Quantifying non-communicable diseases’ burden in Egypt using State-Space model"

### algaeex.png

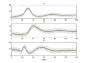

### algaeex_01.png

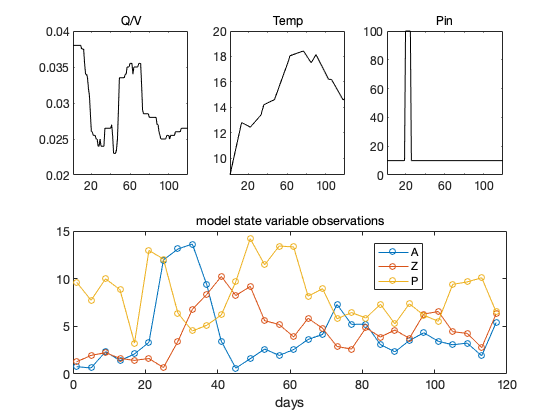

### algaeex_02.png

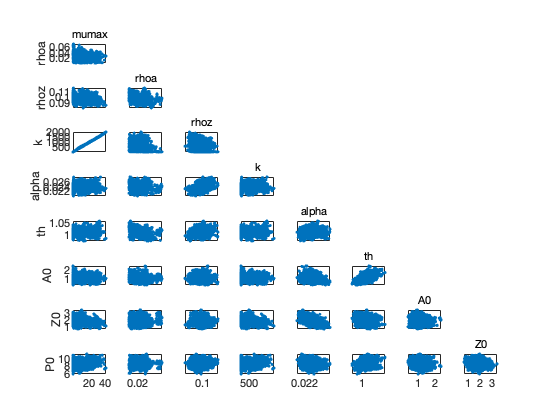

### algaeex_03.png

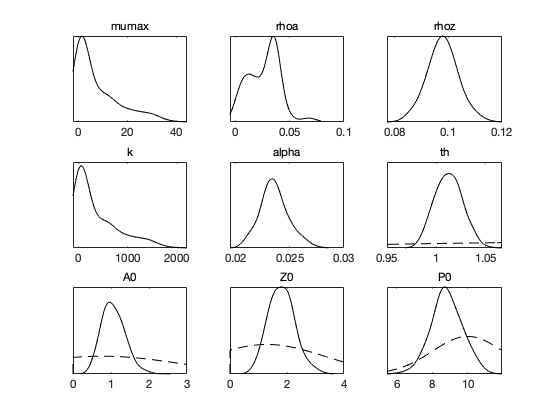

### algaeex_04.png

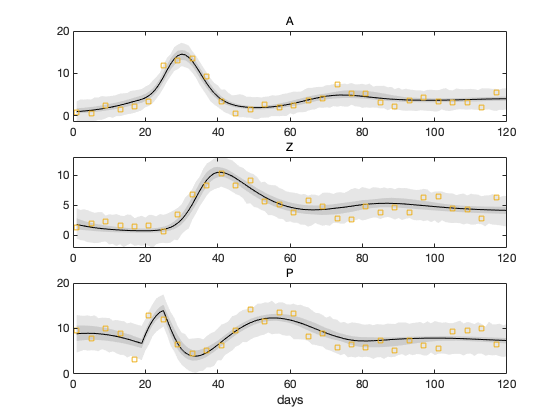

### algaeweb.png

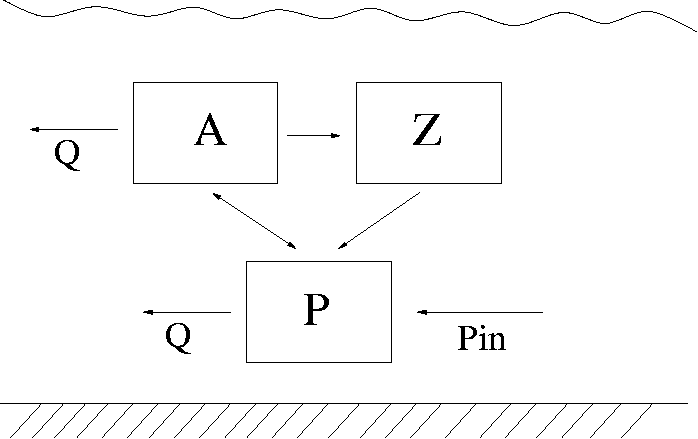

### bananaex.png

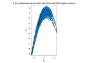

### bananaex_01.png

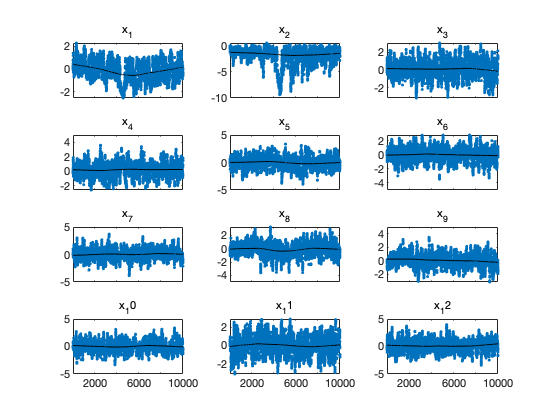

### bananaex_02.png

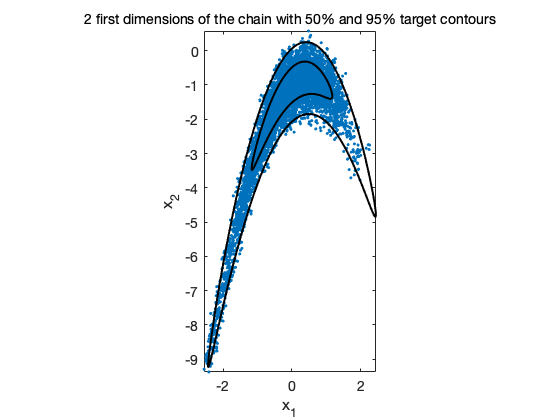

### beetleex.png

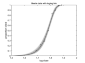

### beetleex_01.png

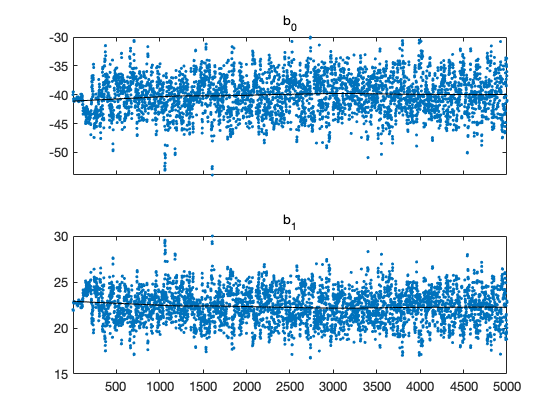

### beetleex_02.png

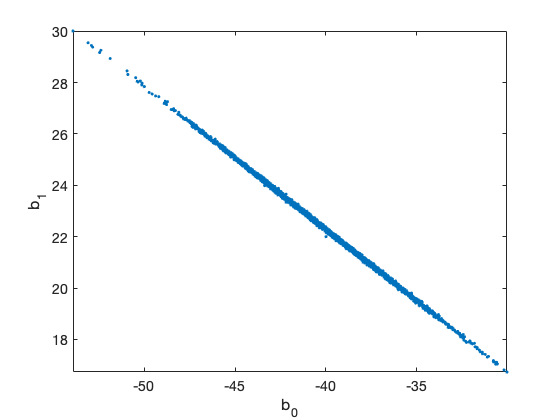

### beetleex_03.png

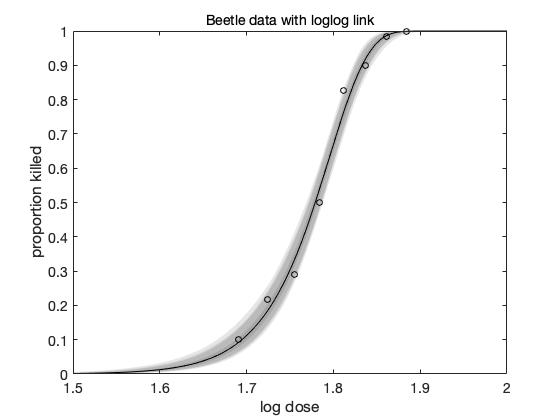

### boxoex.png

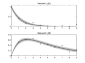

### boxoex_01.png

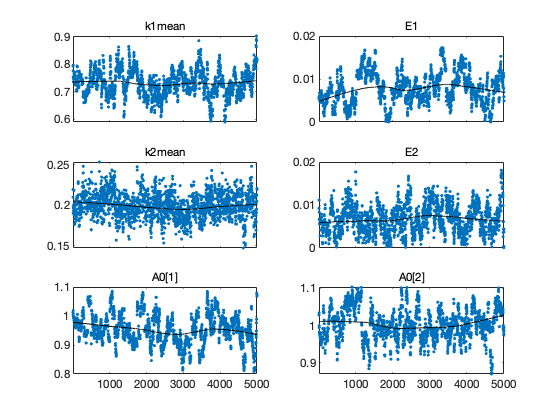

### boxoex_02.png

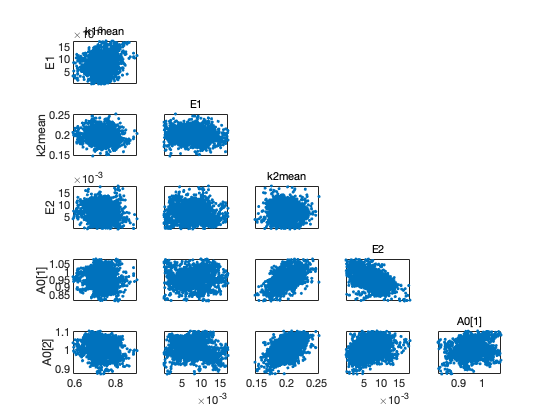

### boxoex_03.png

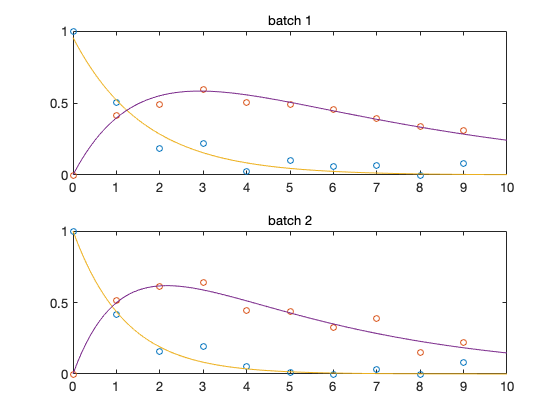

### boxoex_04.png

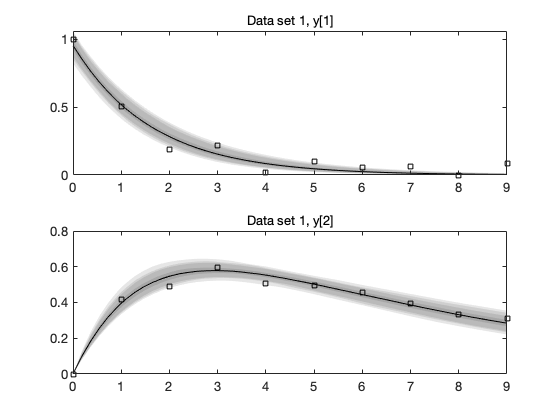

### boxoex_05.png

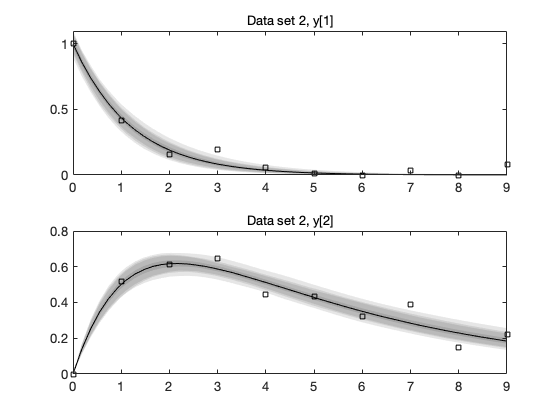

### cauchyex.png

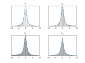

### cauchyex_01.png

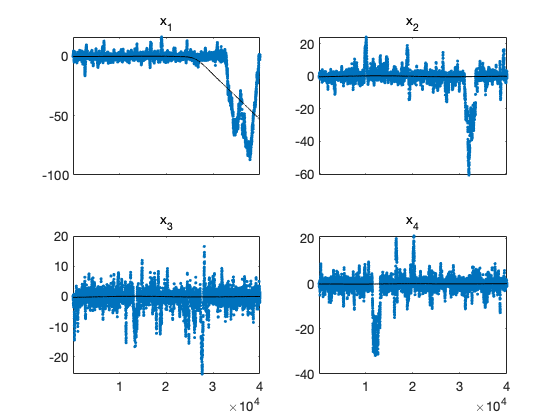

### cauchyex_02.png

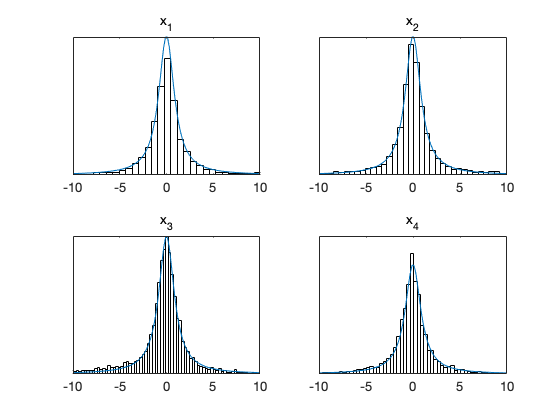

### himmelex.png

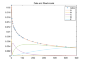

### himmelex_01.png

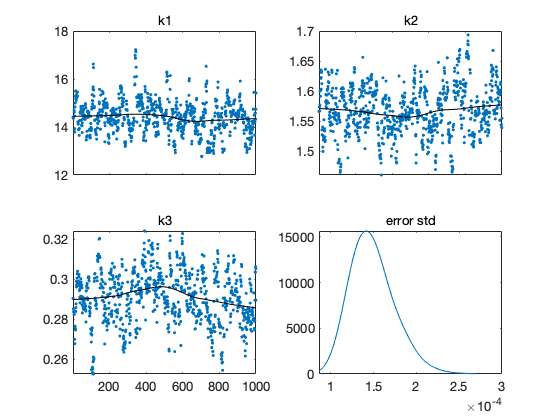

### himmelex_02.png

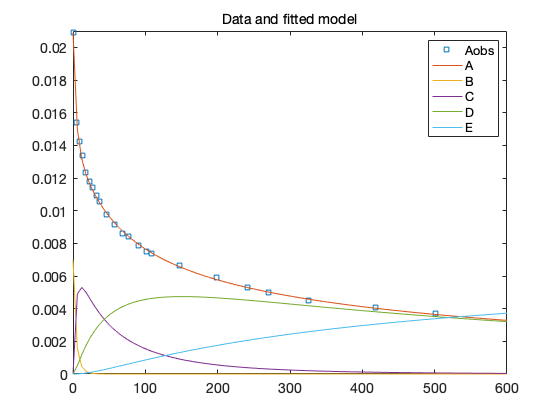

### monodex.png

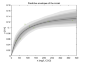

### monodex_01.png

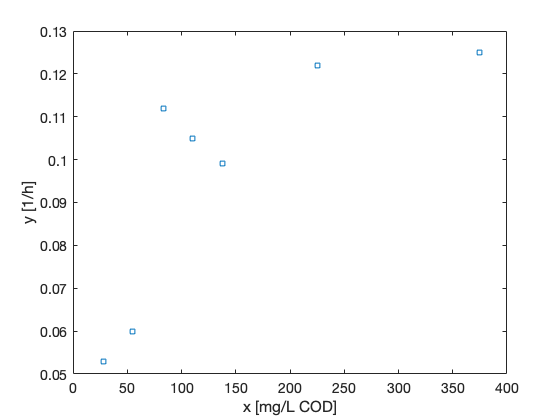

### monodex_02.png

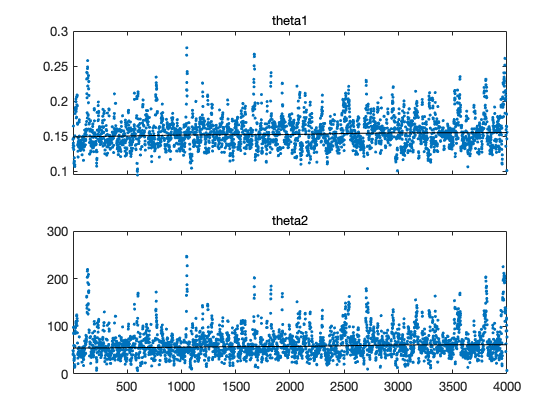

### monodex_03.png

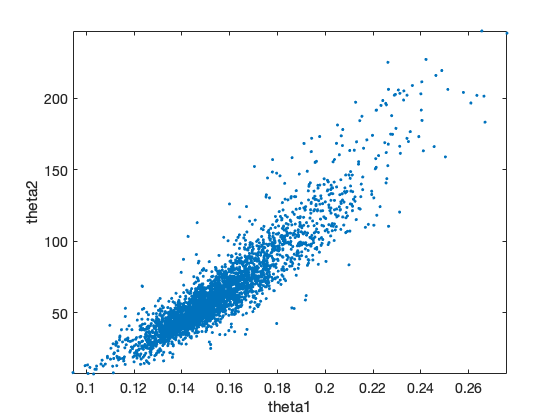

### monodex_04.png

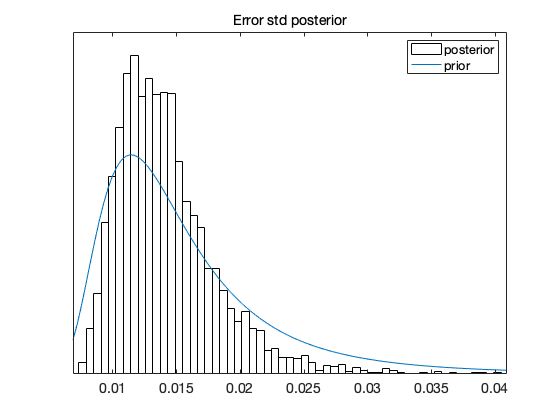

### monodex_05.png

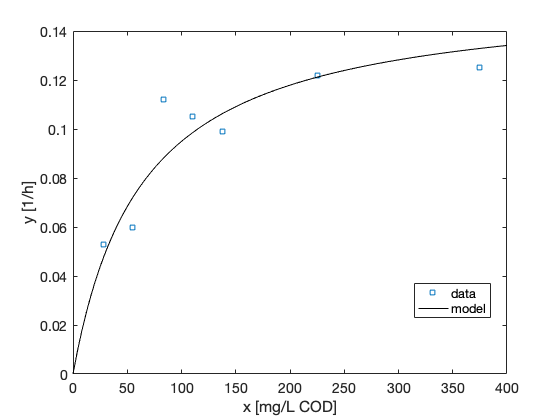

### monodex_06.png

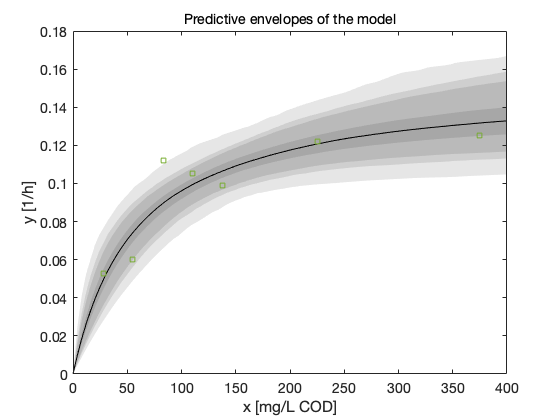

### monodex_eq04958806355743788402.png

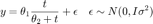

### normalex50.png

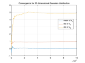

### normalex50_01.png

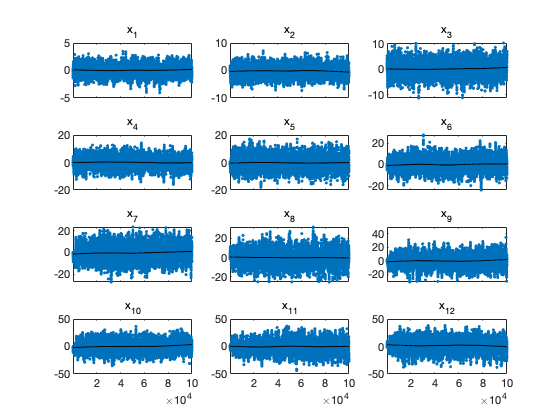

### normalex50_02.png

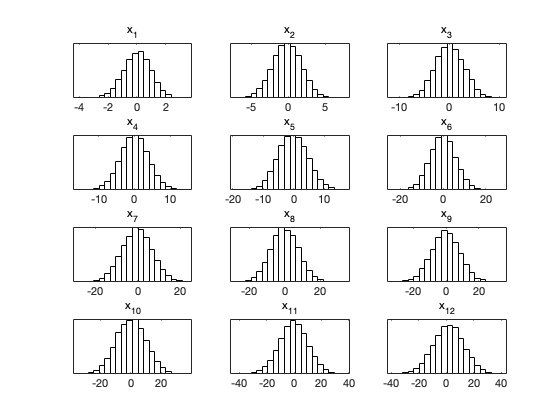

### normalex50_03.png

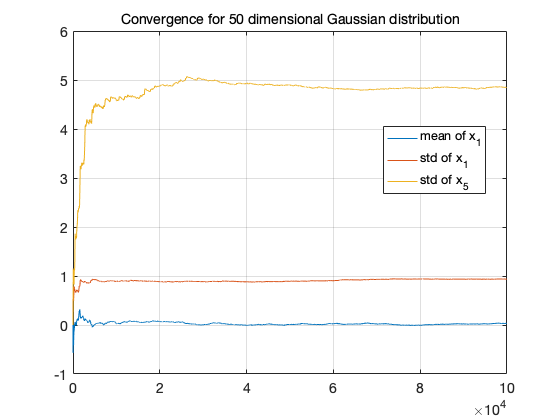

### normalex.png

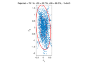

### normalex_01.png

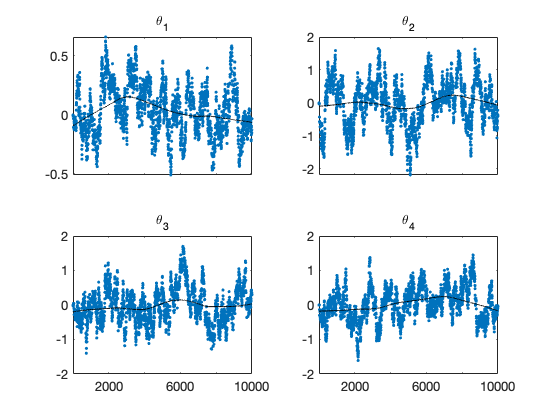

### normalex_02.png

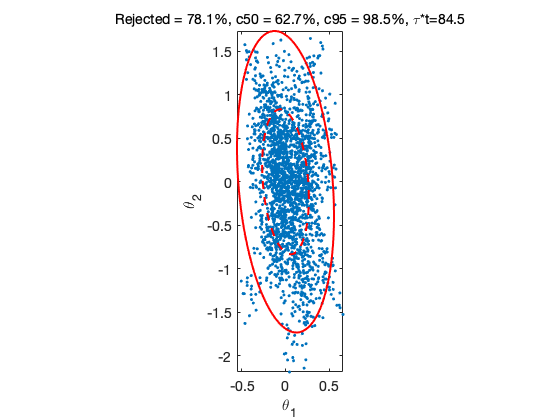

### S1 Figure

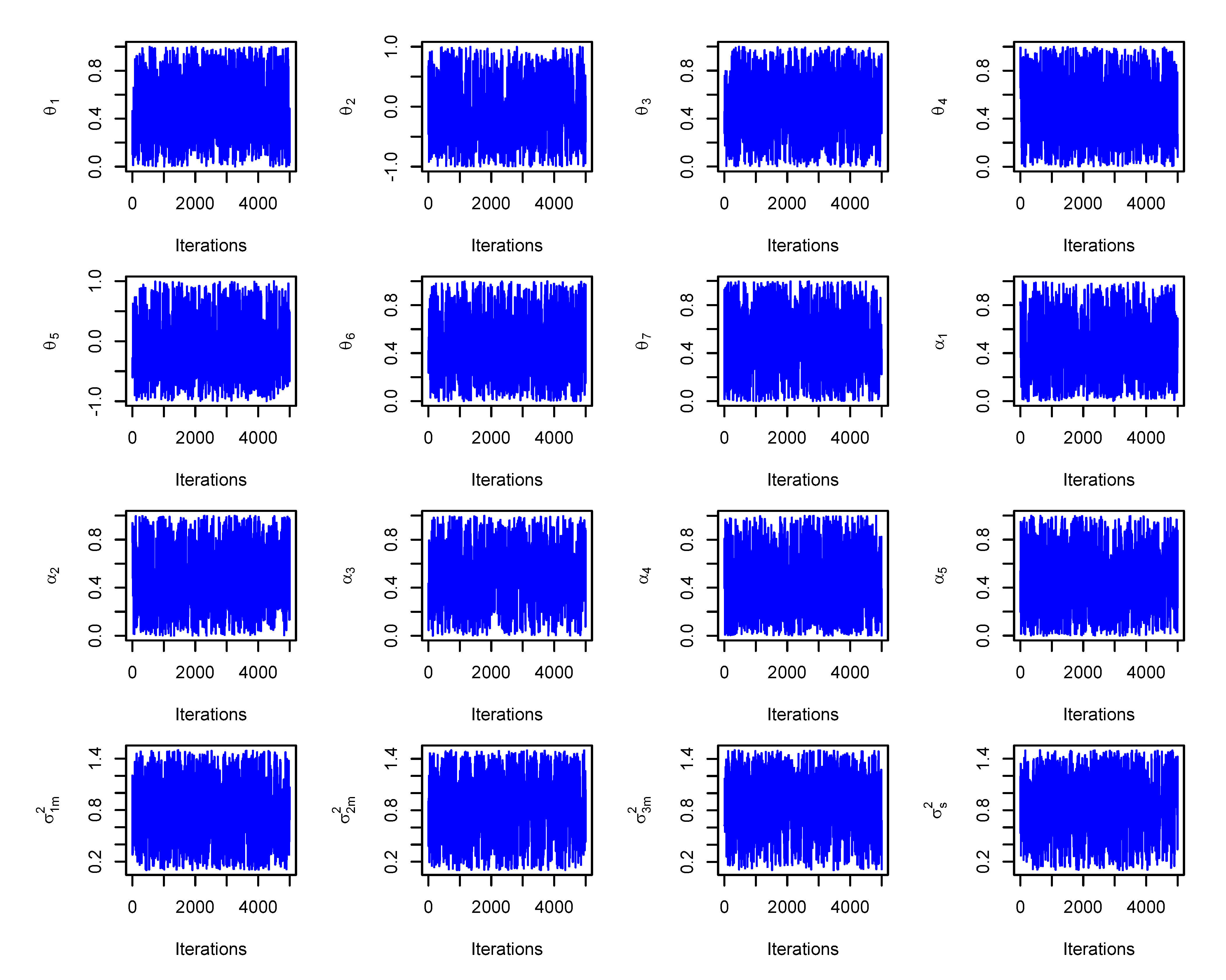

### S2 Figure

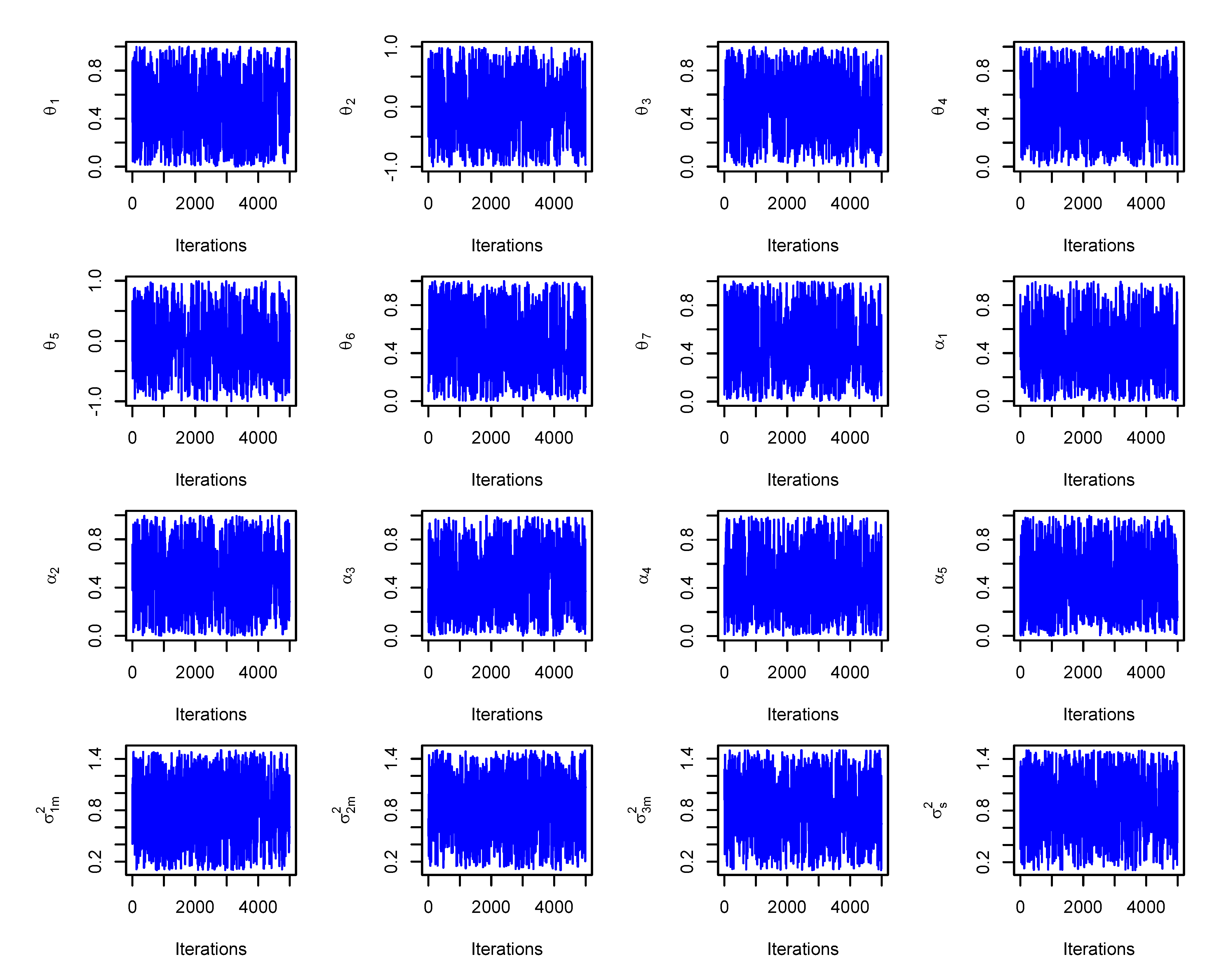

### S3 Figure

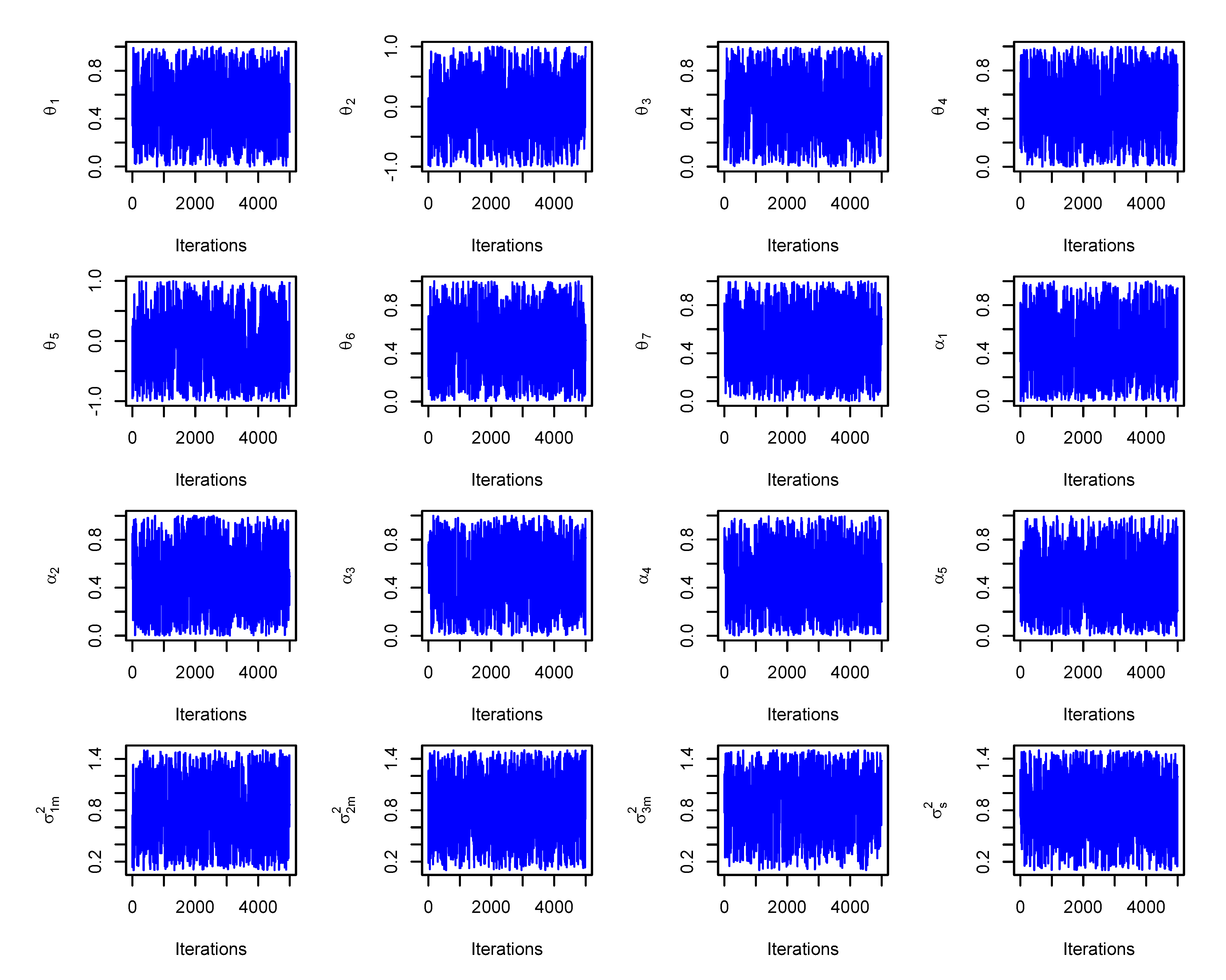

### S4 Figure

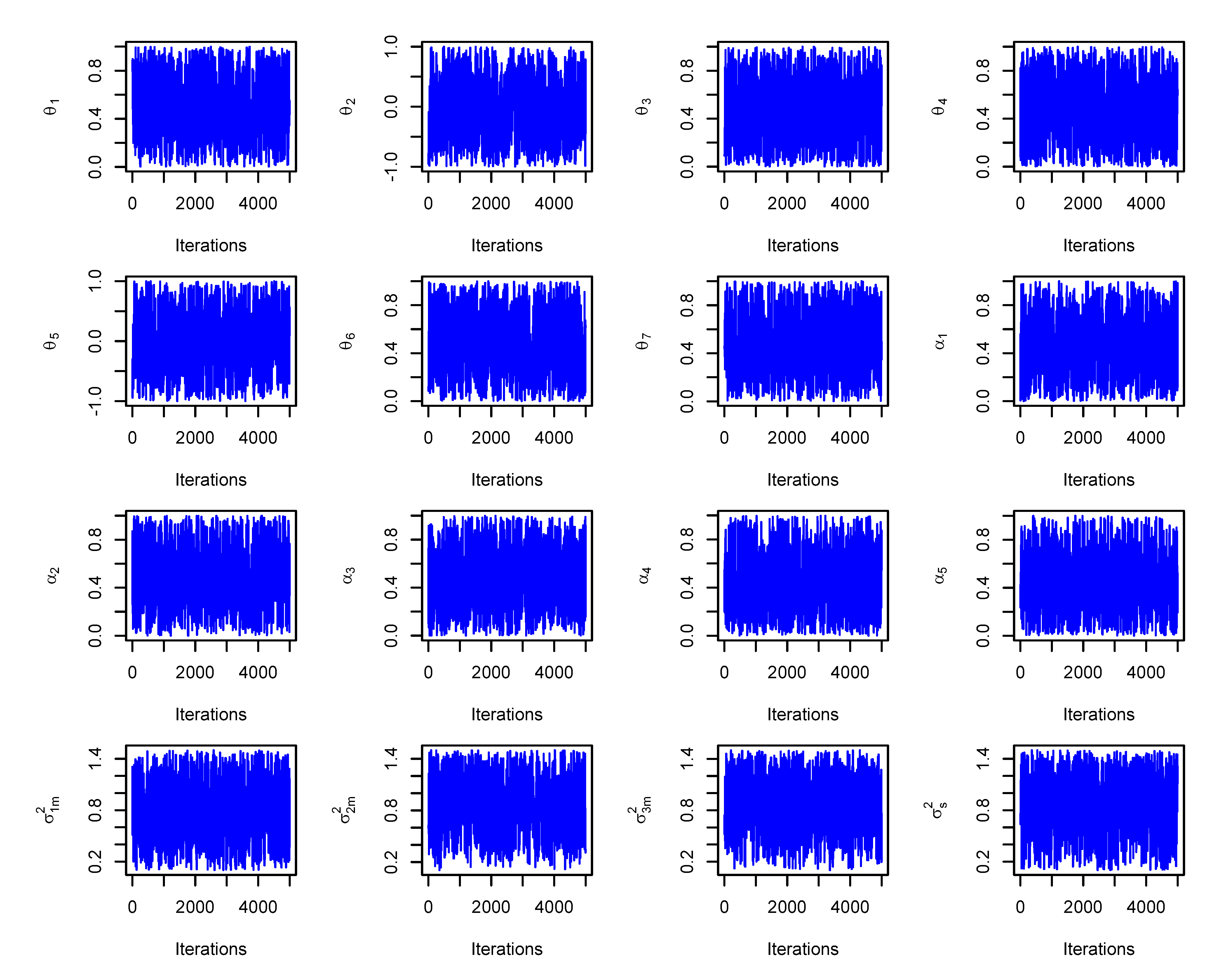

### S5 Figure

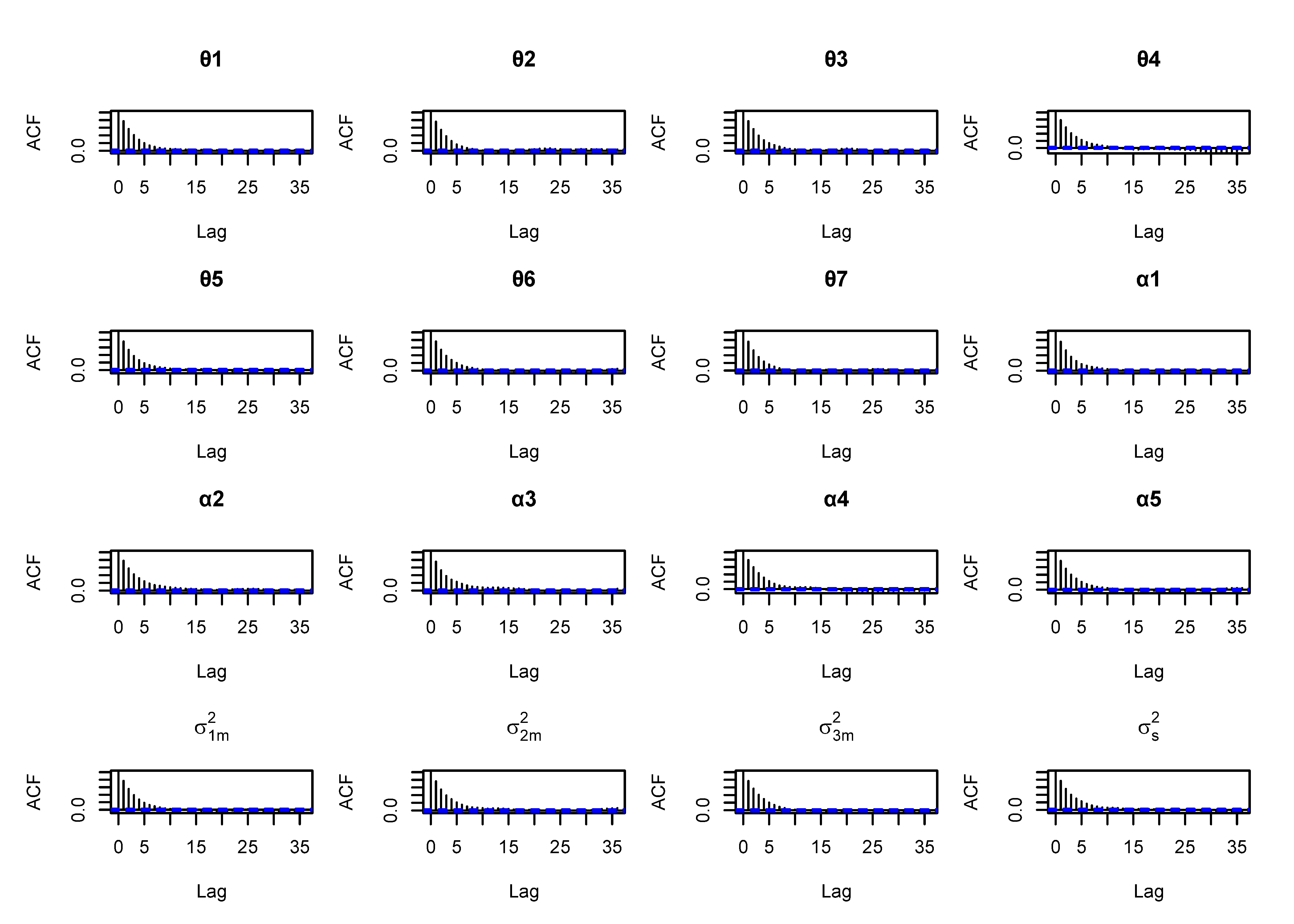

### S6 Figure

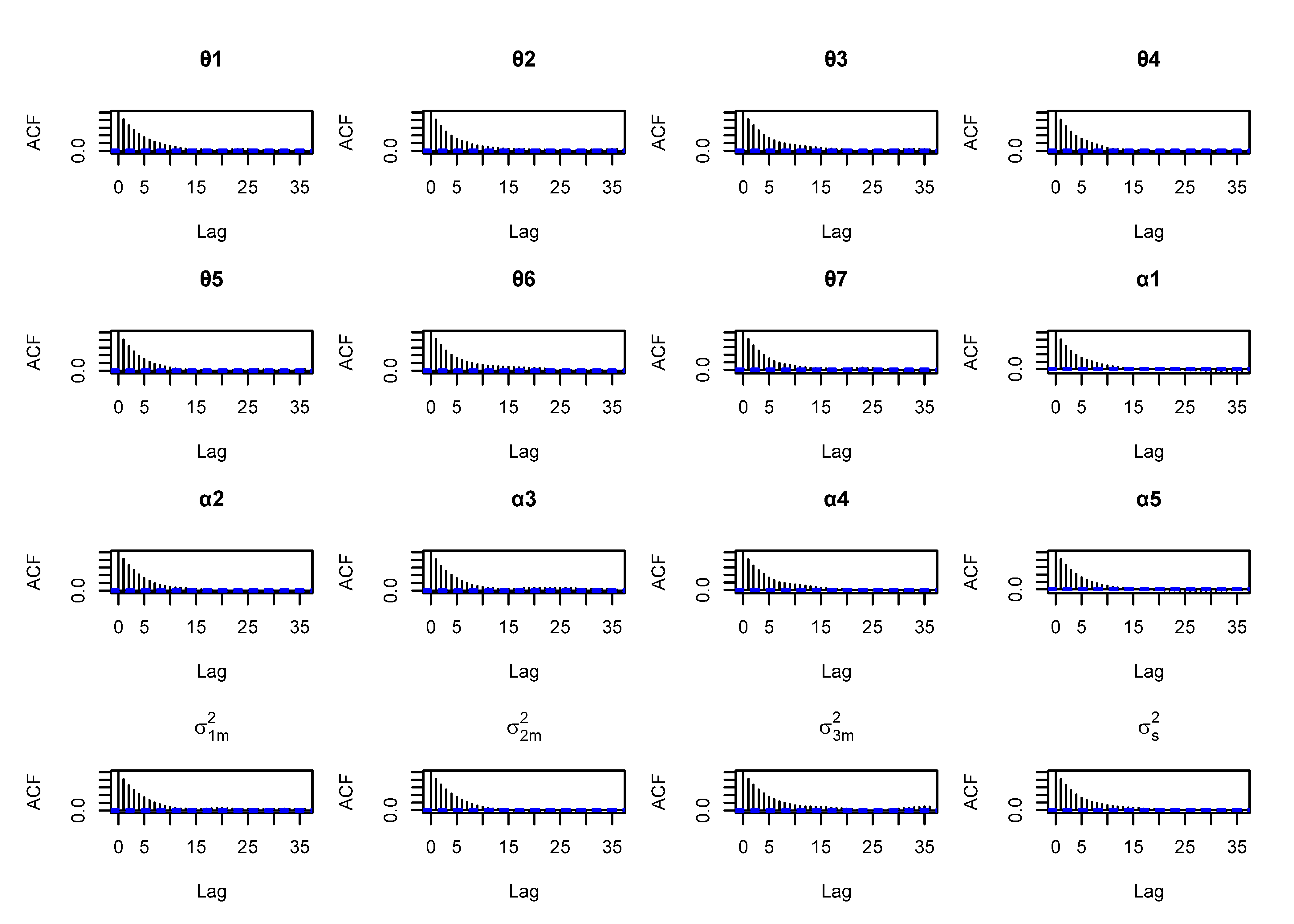

### S7 Figure

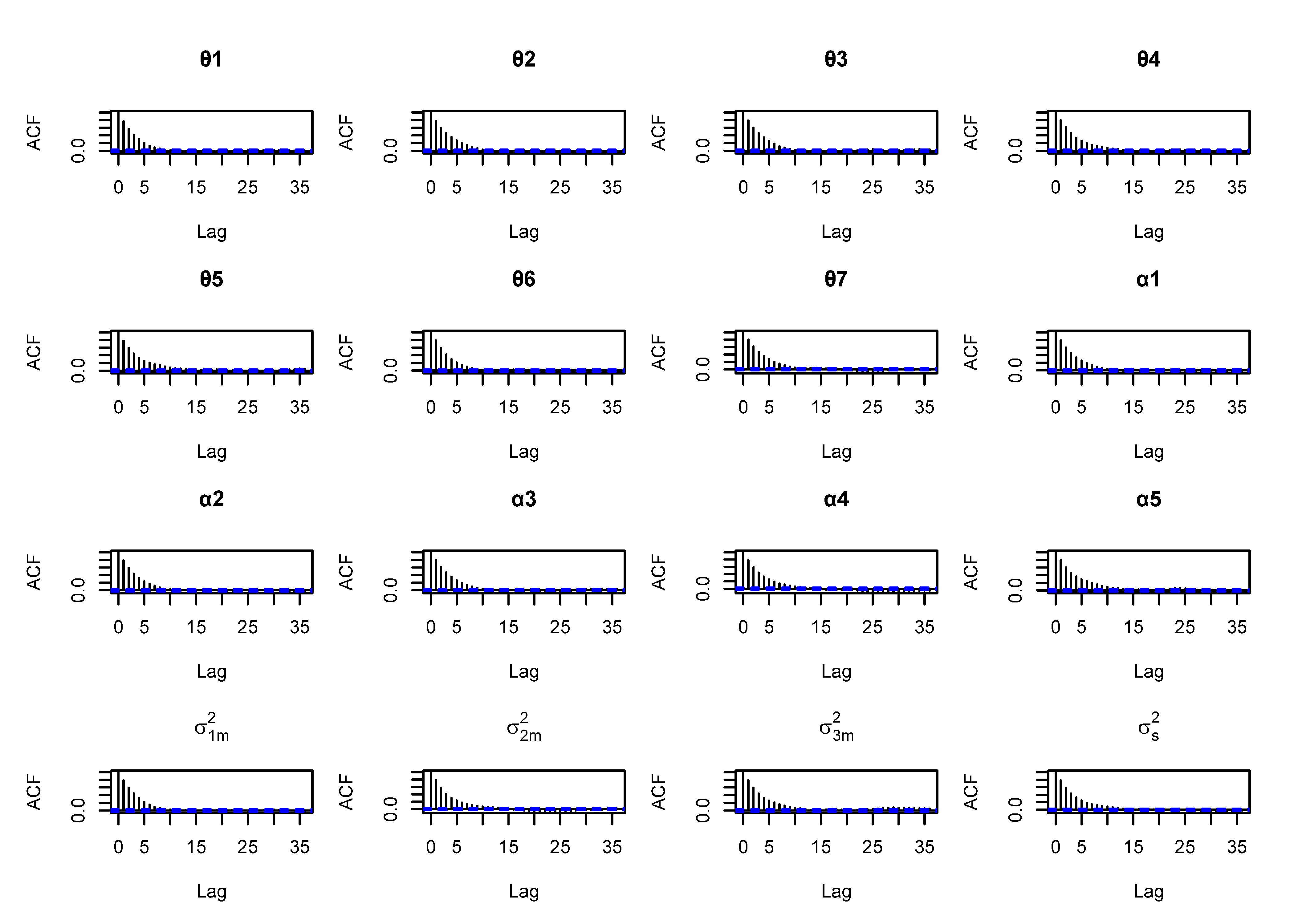

### S8 Figure

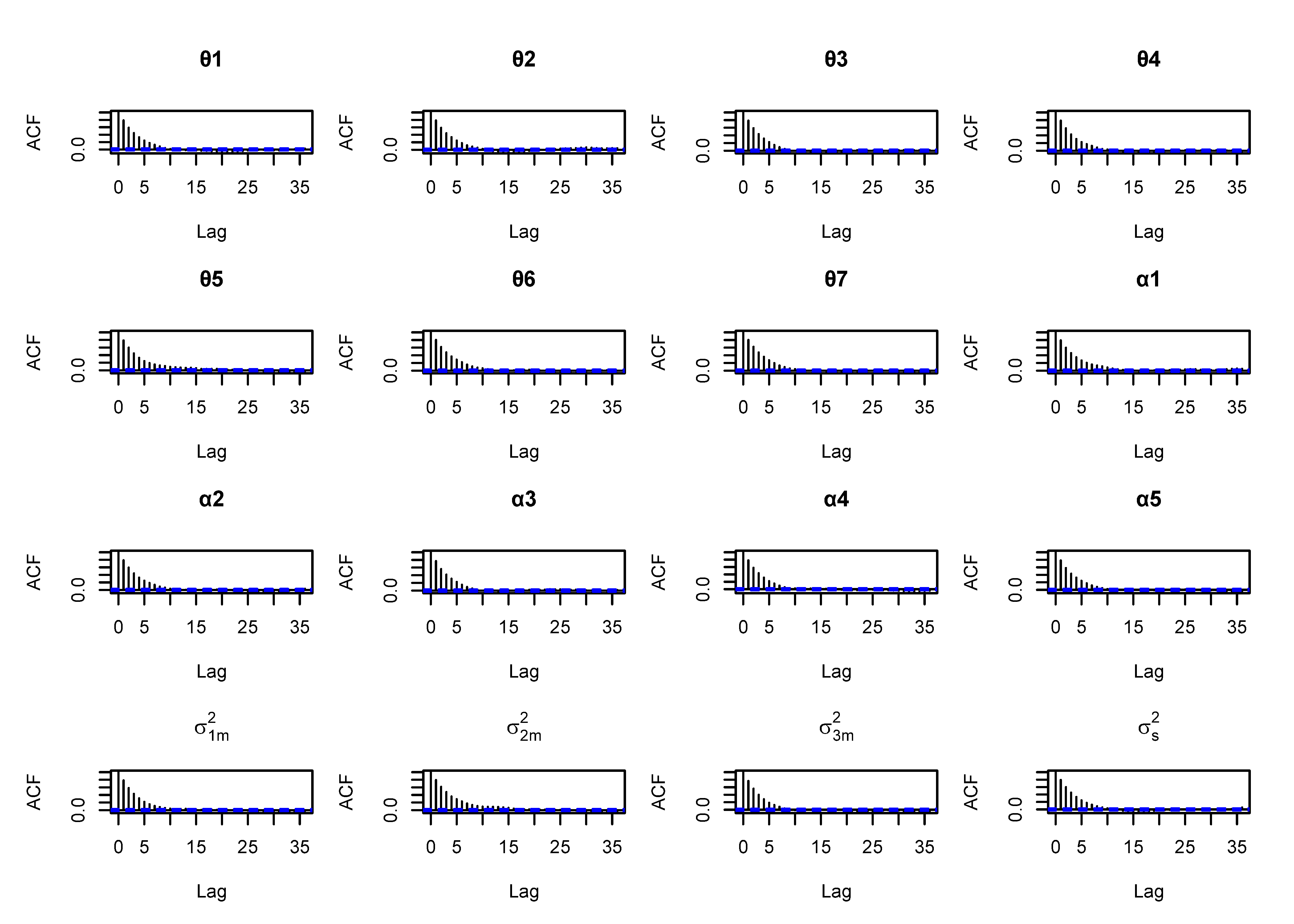

### S9 Figure

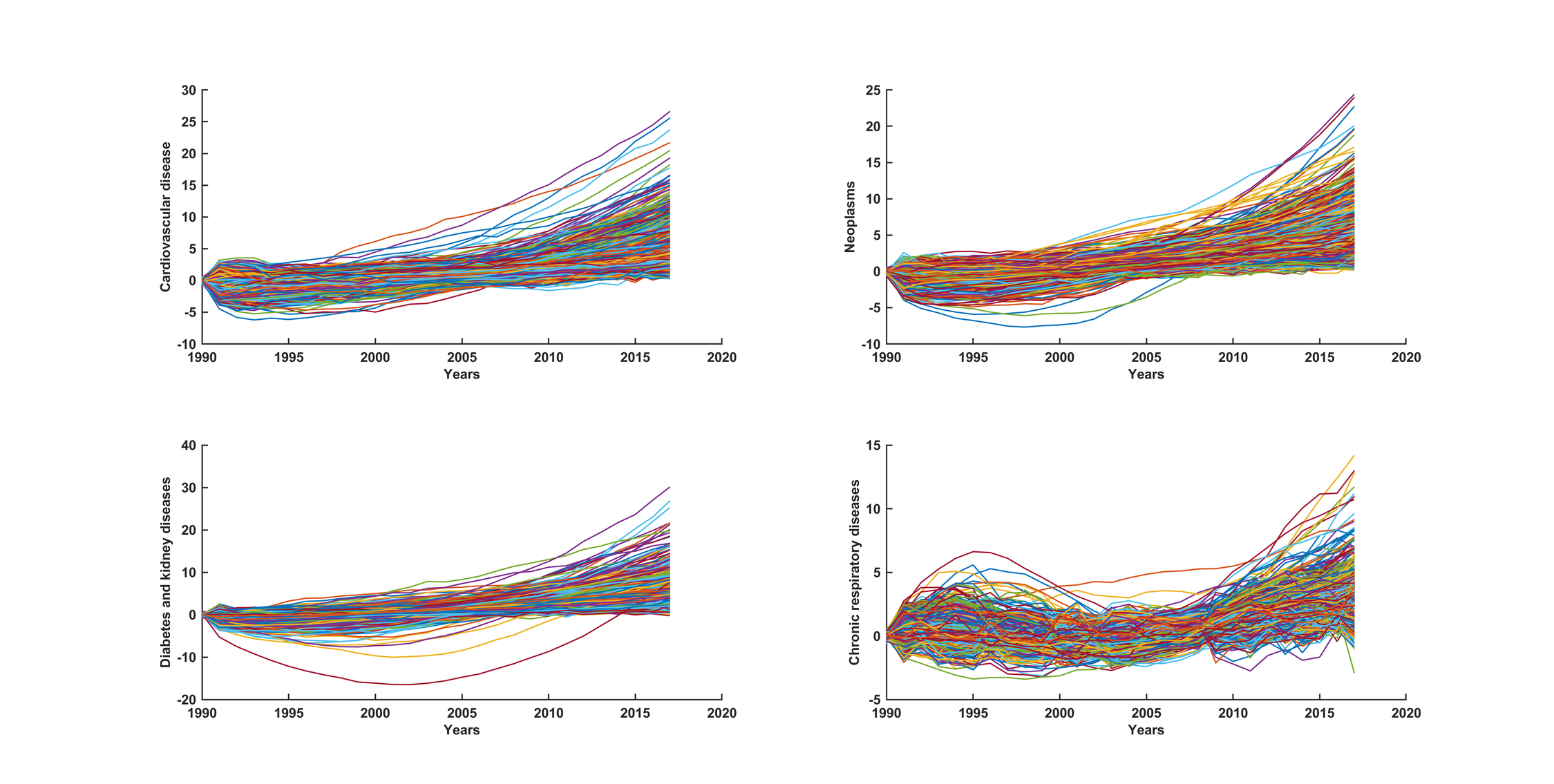

### threemodesex.png

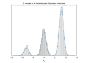

### threemodesex_01.png

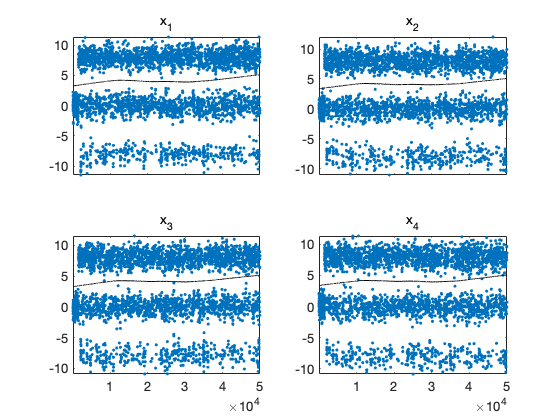

### threemodesex_02.png

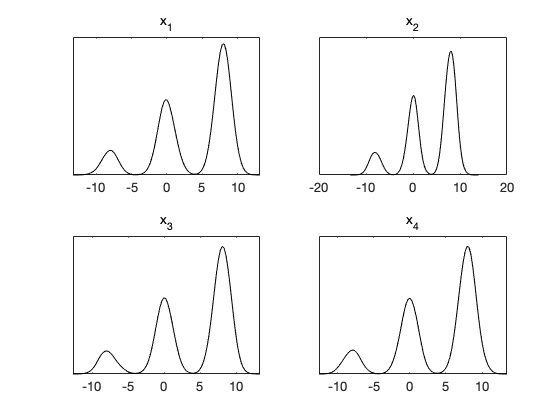

### threemodesex_03.png

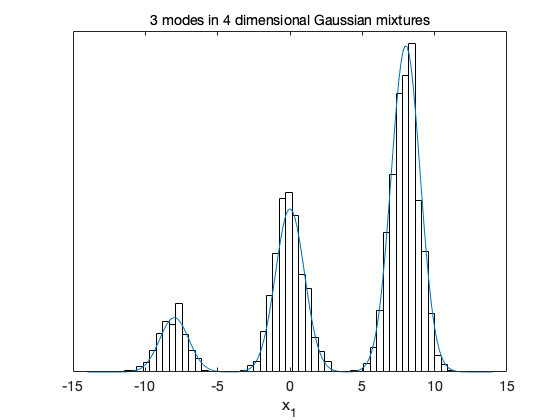
