## Supplementary material for "Quantifying non-communicable diseases’ burden in Egypt using State-Space model": S1 File: Figures_diagnostics_plos.m

```
%Diagnostics 
%open mcmcstat-master, and mcmcdiag
ParaName={'\theta_1','\theta_2','\theta_3','\theta_4','\theta_5','\theta_6','\theta_7','\alpha1','\alpha_2','\alpha_3','\alpha_4','\alpha_5','\sigma^2_{1m}','\sigma^2_{2m}','\sigma^2_{3m}','\sigma^2_{s}','Interpreter','Latex'};
s = strcat(ParaName);
 
%Drawing MCMC tracing plots
r=samplresul([1:16],:);
 for j=1:16             % Plotting McMc sample trace
     subplot(4,4,j);   %for all model parameters
     plot (r(j,:));
     ylabel(ParaName(1,j));
     title('MCMC sample trace');
 end 

%Histogram for PIMH parameters and state
mcmcplot(r',[1:16],s,'hist');
 %Aucorrelation, to chech dependence between samples
 [acf,lags,bounds] = autocorr((samplresul(1,:)'));
 % test for the null hypothesis that the chain converged, see geweke stat
 % (p. value)
 chainstats((samplresul)')
 %Percentage of accepted parameters from proposed distribution
 tabulate(acceptrate)
 %Draw trajectory of xhat
  years=1990:2017;
  plot(years, xhatfiltered(1251:6250,:)); 
 plot(years, xhat); 
%Draw autocorrelation between samples
 mcmcplot((samplresul)',[1:16],s,'acf',100);
  % Thinning the chain, and get new chain , with 5 values difference
  samplresul2=samplresul';
  z= thin((sampl)' ,1251,5,6250);
  % Checking of autocorrelation after thinning, the chain is 1000 values
  % (good mixing)
  mcmcplot(z,[1:15],s,'acf',10)
  
  
  
 % Drawing density of the state variable (particle filter)
 [f,xi] = ksdensity(x); 

plot(xi,f);
 %online estimation(particle filter)
 years=1990:2017;
 plot(years, zhat(:,1),'DisplayName','Burden of diabetes and kidney diseases')
 plot(years,zhat(:,2),'DisplayName','\theta1')
 hold on
 plot(years,zhat(:,3),'DisplayName','\theta2')
 plot(years,zhat(:,4),'DisplayName','\theta3')
 plot(years,zhat(:,5),'DisplayName','\theta4')
 plot(years,zhat(:,6),'DisplayName','\theta5')
 plot(years,zhat(:,7),'DisplayName','\theta6')
 plot(years,zhat(:,8),'DisplayName','\theta7')
 plot(years,zhat(:,9),'DisplayName','\alpha1')
 plot(years,zhat(:,10),'DisplayName','\alpha2')
 plot(years,zhat(:,11),'DisplayName','\alpha3')
 plot(years,zhat(:,12),'DisplayName','\alpha4')
 %plot(years,zhat(:,13),'DisplayName','\alpha5')
 plot(years,zhat(:,14),'DisplayName','\sigma^2_{1m}')
 plot(years,zhat(:,15),'DisplayName','\sigma^2_{2m}')
 plot(years,zhat(:,16),'DisplayName','\sigma^2_{3m}')
 plot(years,zhat(:,17),'DisplayName','\sigma^2_{s}')
 hold off
 
 
 


  
```
