## Supplementary material for "Quantifying non-communicable diseases’ burden in Egypt using State-Space model": S1 File: computenextstate_mytrial3.m

```
function x = computenextstate_mytrial3(z,u,snoise)
%load ( "variable_imputedwithbounds1","risk1","prevneop","dmneop","dsneop");
% risk1 is combined risk factor, u2 is the prevalence, u3 is the average
% disability weight of mild and moderate cases, u4 is the average
% disability weight of severe cases

x= z(:,9).*z(:,1)+ z(:,10).*u(:,1)+z(:,11).*u(:,2)+z(:,12).*u(:,3)+z(:,13).*u(:,4)+snoise';
end
 
```
