## Supplementary material for "Quantifying non-communicable diseases’ burden in Egypt using State-Space model": S1 File: My_trial_computey3.m

```
function y = My_trial_computey3(z,z_data,mnoise)
%load ( "variable_imputedwithbounds1", "beds1", "specneop1","gdp");
%z1 is the number of beds, z2 is the number of specialists, z3 is the GDP
%per capita
y1=z(:,2).*z(:,1)+mnoise(1,1);
y2=z(:,3).*z(:,1)+z(:,4).*z_data(:,2)+z(:,5).*z_data(:,3)+mnoise(1,2);
y3=z(:,6).*z(:,1)+z(:,7).*z_data(:,1)+z(:,8).*z_data(:,3)+mnoise(1,3);
y(:,1)=y1;
y(:,2)=y2;
y(:,3)=y3;
end
```
