## Supplementary material for "Quantifying non-communicable diseases’ burden in Egypt using State-Space model": S1 File: my_trial_computey4.m

```
% State transition equations

function [y] = my_trial_computey4(x,z_data, paradox,mnoise)
%load ( "variable_imputedwithbounds1", "beds1", "specneop1","gdp");
y1=paradox(1).*x+mnoise(1,1);
y2=paradox(2).*x+paradox(3).*z_data(:,2)+paradox(4).*z_data(:,3)+mnoise(1,2);
y3=paradox(5).*x+paradox(6).*z_data(:,1)+paradox(7).*z_data(:,3)+mnoise(1,3);
y(:,1)=y1;
y(:,2)=y2;
y(:,3)=y3;
end
```
