## Supplementary material for "Quantifying non-communicable diseases’ burden in Egypt using State-Space model": S1 File: resampling.m

```
function index=resampling(q)

        qc=cumsum(q);
        M=length(q);
       % u=(0:M-1+rand(1))/M;
        u=(0:M-1)/M;
        u = u + rand()/M; % MOHAMED SALEH 4.MAY.2019
        index=zeros(1,M); 
        k=1;
        for j=1:M
            while(qc(k)<u(j))
                k=k+1;
            end
            index(j)=k;
        end
```
