## Supplementary material for "Quantifying non-communicable diseases’ burden in Egypt using State-Space model": S1 File: Particle_filter.m

```
%Online estimation (Particle Filter)
%%Initialize and define variables
% format long g;
tic
N= 1000;                % Number of particles
T=28;
spec(isnan(spec))=mean(spec(~isnan(spec)));      % fill the missing values with average, input variables
beds(isnan(beds))=mean(beds(~isnan(beds)));

%%
%Varaibles standardization
beds=(beds-mean(beds))/std(beds);
spec=(spec-mean(spec))/std(spec);
days1=(days1-nanmean(days1))/nanstd(days1);
risk1=(risk1-mean(risk1))/std(risk1);
outpat=(outpat-nanmean(outpat))/nanstd(outpat);
prev=(prev-mean(prev))/std(prev);
dwm=eps((dwm-mean(dwm)))/eps(std(dwm));
dws=eps(dws-nanmean(dws))/eps(nanstd(dws));
igdppercapusd10=(igdppercapusd10-mean(igdppercapusd10))/std(igdppercapusd10);
yll=(yll-mean(yll))/std(yll);
%%
 %Number of Particles
noParticles=N;
x=unifrnd(-3,3,N,1);% i used the distribution of the first indicator y1

%The distribution of parameters
theta = unifrnd(0,1, N,7);
theta(:,2)=unifrnd(-1,1, N,1);
theta(:,5)=unifrnd(-1,1, N,1);
alpha = unifrnd(0,1, N,5);
Mu=zeros(4,N);
pw=unifrnd(0.1,1.5, N,4);


%%
%  control variables or determinants of transition equation
% risk1 Factor created by time series factor analysis to risk factors variables 
%prev Prevalence of disease 
%dwm Average of the mild and moderate disability weights
% dws Average of severe disability weights

u=[risk1,prev,dwm,dws];

%% initiate empty matrices for Model parameters and inputs, assuming initial values for the
%parameters,
%I have five alphas in the state equation, 7 thetas in the observation equations,and
%three meausrement error variances
%equation
% z is the vector of unknowns
z=[x,theta(:,1),theta(:,2),theta(:,3),theta(:,4),theta(:,5),theta(:,6),theta(:,7),alpha(:,1),alpha(:,2),alpha(:,3),alpha(:,4),alpha(:,5),pw(:,1),pw(:,2),pw(:,3),pw(:,4)];

%Vector for each estimated parameter

zhat=ones(T,17);
zhat(1,:)=[mean(x),mean(theta(:,1)),mean(theta(:,2)),mean(theta(:,3)),mean(theta(:,4)),mean(theta(:,5)),mean(theta(:,6)),mean(theta(:,7)),mean(alpha(:,1)),mean(alpha(:,2)),mean(alpha(:,3)),mean(alpha(:,4)),mean(alpha(:,5)),mean(pw(:,1)),mean(pw(:,2)),mean(pw(:,3)),mean(pw(:,4))];          %Initial values

% Data, y variables, measurements , or indicators

y_data=ones(T,3);

y1_data = yll;             %Number of years lost due to death
y2_data = outpat;          %Number of outpatients
y3_data=  days1;           %Number of days spent in a hospital

y_data(:,1)= y1_data;
y_data(:,2)= y2_data;
y_data(:,3)= y3_data;

%%Input variable of meausrement equations
  %beds1 Number of beds
 %specneop1 Number of specialists
 %GDp per capita in usd2010
 
z_data=[beds,spec,igdppercapusd10];


RMSD=zeros(T,3);


%%
for t = 2:T
 % The distribution of state and observation noises

snoise=mvnrnd(Mu(4,:),(z(:,17))');

    % time varying error in transition equations of the parameters
    eta=normrnd(0,0.01,[1,16]);
   
   %prediction next_state, next parameter & y_hat
   
       x = computenextstate_mytrial3(z,u(t-1,:),snoise);
       z(:,1)=x;  
       z(:,2:17)=z(:,2:17)+eta(1:16);
       mnoise(1,:)=mvnrnd(Mu(1,:),(z(:,14))');
       mnoise(2,:)=mvnrnd(Mu(2,:),(z(:,15))');
       mnoise(3,:)=mvnrnd(Mu(3,:),(z(:,16))');
       y= My_trial_computey3(z,z_data(t,:),mnoise); 

        e1 =repmat(y1_data(t),N,1)-y(:,1);%Prediction Error of y1
        e2 =repmat(y2_data(t),N,1)-y(:,2);%Prediction Error of y2
        e3 =repmat(y3_data(t),N,1)-y(:,3);%Prediction Error of y3
        %compute the difference between true value and predicted value(RMSD)
        RMSD(t,1)=(nansum(e1.^2))/N;
        RMSD(t,2)=(nansum(e2.^2))/N;
        RMSD(t,3)=(nansum(e3.^2))/N;
   % Compute importance weights according to the available information of y's, with if statements includes all alternatives     
        %%%%%%%%%%%%%%%%%%%%%%%%%%%%%%%%%%%%%%%%%%%%%%%%%%%%%%%%%%%%%%%%%%%%%%%%%%%%%%%%%%%%%%%%
       if isnan(y_data(t,1))
        logl1=0;
       else
        logl1=-1/2*log(2*pi*z(:,14)) - 1/(2*z(:,14))*e1.^2;  
       end
        %%%%%%%%%%%%%%%%%%%%%%%%%%%%%%%%%%%%%%%%%%%%%%%%%%%%%%%%
        if isnan(y_data(t,2))
        logl2=0;
        else
        logl2=-1/2*log(2*pi*z(:,15)) - 1/(2*z(:,15))*e2.^2;    
        end
  %%%%%%%%%%%%%%%%%%%%%%%%%%%%%%%%%%%%%%%%%%%%%%%%%%%%%      
        if isnan(y_data(t,3))
        logl3=0;        
       else
        logl3=-1/2*log(2*pi*z(:,16)) - 1/(2*z(:,16))*e3.^2;   
        end
        
        logweights=logl1+logl2+logl3; %Weight calculations, log(l1*l2*l3)    
        const = max(logweights); % Subtract the maximum value for numerical stability, shift for better calculation
        weights = eps(exp(logweights-const));
        nw= weights/sum(weights);  % Save the normalized weights 
     %%%%%%%%%%%%%%%%%%%%%%%%%%%%%%%%%%%%%%%%%%%%%%%%%%%%%%%%%%%%%%%%%%%%%
         
      %State and parameter estimation
     zhat(t,:)= nw'*z(:,1:17);
     %Mean square error (MSE)
     for g=1:17
     MSE(t,g)=  (sum(((z(:,g)))-(repmat(zhat(t,g),N,1))).^2)/N;
     end 
     index=resampling(nw);   %calling resampling function
     z=z((index)',:);              %Resampled Particles 
     
end


Zhat=array2table(zhat, 'VariableNames', {'State','theta1', 'theta2', 'theta3','theta4','theta5','theta6','theta7','alpha1', 'alpha2', 'alpha3','alpha4','alpha5','w1', 'w2', 'w3','w4'});
 %Theil inequality
 RMSD=sqrt(nansum(RMSD(:,1:3))/T);
for j=1:17
 RMSE(j)=sqrt(sum(MSE(:,j))/T);
 MSE1 (j)=  (sum(MSE(:,j)))/T;
 
 end
  toc
```
