## Supplementary material for "Quantifying non-communicable diseases’ burden in Egypt using State-Space model": S1 File: PIMH.m

```
%particle independent metropolis hastings
%%Initialize and define variables
tic
noIterations=5000;
N= 1000;                %Number of Particles
noburniniteration=0.2;  %start convergence after 20% of the iterations
T=28;
x=unifrnd(-3,3,N,1);
%% Standardization of variables
beds=(beds-mean(beds))/std(beds);
spec=(spec-mean(spec))/std(spec);
days1=(days1-nanmean(days1))/nanstd(days1);
risk1=(risk1-mean(risk1))/std(risk1);
outpat=(outpat-nanmean(outpat))/nanstd(outpat);
prev=(prev-mean(prev))/std(prev);
dwm=eps(dwm-mean(dwm))/eps(std(dwm));
%dws=eps(dws-mean(dws))/eps(std(dws));
 
igdppercapusd10=(igdppercapusd10-mean(igdppercapusd10))/std(igdppercapusd10);
yll=(yll-mean(yll))/std(yll);
%% We have three measurement equations
%%Input variable of meausrement equations
  %beds1 Number of beds
 %spec Number of specialists
 %GDp per capita in usd2010
z_data=[beds,spec,igdppercapusd10];
%% %  control variables or determinants of transition equation
  %u1 Factor created by time series factor analysis to risk factors variables 
  % u2 Prevalence of disease
  % u3 Average of the mild and moderate disability weights
   %u4  Average of severe disability weights
u=[risk1,prev,dwm,dws];

sampl=zeros(16, noIterations/(1-noburniniteration));
xhatfiltered=zeros(noIterations/(1-noburniniteration), T);

%% initiate empty matrices for Model parameters and inputs, assuming initial values for the
%parameters,
%I have five alphas in the state equation and 7 thetas in the observation,
%3 observation errors and one state error
%equation
theta=zeros(1,7);
alpha=zeros(1,5);
pw=ones(1,4);
Mu=zeros(4,1);
mnoise=mvnrnd(Mu(1:3,:),pw(:,1:3));
snoise=mvnrnd(Mu(4,:),pw(:,4));

% Number of parameters
para0 = [0;0;0;0;0;0;0;0;0;0;0;0;0.01;0.01;0.01;0.01 ];
priordispar= [0 1; -1 1; 0 1;0 1; -1 1; 0 1; 0  1; 0 1;0 1;0 1;0 1;0 1;0.1    1.5;0.1       1.5;0.1   1.5;0.1   1.5];      %Uniform prior disetribution of the parameters 
para1 =zeros(16,1);
%Initial values for the parameters are the first column in sampl matrix
sampl(:,1)=para0;                                                                            % Initial sample of parameters

% Data

y_data=ones(28,3);

y1_data = yll;
y2_data = outpat;
y3_data=  days1;

y_data(:,1)= y1_data;
y_data(:,2)= y2_data;
y_data(:,3)= y3_data;


%% MCMC iterations
    para0proposed=zeros(16,noIterations/(1-noburniniteration));
    para1proposed=zeros(16,noIterations/(1-noburniniteration));
   
    para1(:,1)=para0(:,1);
    para0proposed(:,1)=para0(:,1);
    para1proposed(:,1)= para1(:,1);              % The first column is the initial values in paraproposed 1 and 0
    
     
     
    %% estimation of  initial likelihood using initial values of parameters
for i=1
     xhat=zeros (1,T);
     xhat(1)=mean(x);                % The first value at(t=1) in xhat is the mean of the simulated values from the assumed dist
     loglik0=0;
    
    
for t = 2:T
paradox=para1proposed(:,i);
    %newx=x1proposed(:,i);
   % Compute importance weights
   %step3:prediction next_state &
   
pw=ones(1,4);
Mu=zeros(4,1);
pw(:,1:4)=para1proposed(13:16,i);
mnoise=mvnrnd(Mu(1:3,:),pw(:,1:3));
snoise=mvnrnd(Mu(4,:),pw(:,4));

        x = computenextstate_mytrial4(x,u(t-1,:), paradox,snoise);
        y= my_trial_computey4(x,z_data(t,:), paradox,mnoise);
       
        %STEP4:Compute Weight
        e1 =repmat(y1_data(t),N,1)-y(:,1);%Prediction Error of y1
        e2 =repmat(y2_data(t),N,1)-y(:,2);%Prediction Error of y2
        e3 =repmat(y3_data(t),N,1)-y(:,3);%Prediction Error of y3
        
        
        
        logl1 = -1/2*log(2*pi*(para1proposed(13,i))) - 1/(2*(para1proposed(13,i)))*e1.^2;
        logl2 = -1/2*log(2*pi*(para1proposed(14,i))) - 1/(2*(para1proposed(14,i)))*e2.^2;
        logl3 = -1/2*log(2*pi*(para1proposed(15,i))) - 1/(2*(para1proposed(15,i)))*e3.^2;
        
    %% 
    
    logweights= logl1+ logl2+logl3; %Weight calculations, log(l1*l2*l3)
    const = max(logweights); % Subtract the maximum value for numerical stability
    weights = exp(logweights-const);
    w= weights/sum(weights);  % Save the normalized weights
    loglik0=loglik0+const+log(sum(weights))-log(N);
     %Step5:State estimation
     xhat(t)=w'*x;  %Estimate for state 
     %Step6:Resampling
     index=resampling(w);   %calling resampling function
      x=x((index)',1);              %Resampled Particles
 end %Step 7:End time loopl1
 end
   %%
   xhatfiltered(1,:)=xhat; 
   loglikproposed=zeros(noIterations/(1-noburniniteration),1);
   loglikproposed(1)=loglik0;
   acceptrate=zeros(noIterations/(1-noburniniteration),1);
   acceptprobproposed=zeros(noIterations/(1-noburniniteration),1);
   acceptrate(1)=1;
   %%
for i=2:noIterations/(1-noburniniteration)
    %Propose new parameters
    para1proposed(13:16,i)=unifrnd(0.1,1.5,4,1);
    para1proposed(1,i)=  rand(1,1);                 
    para1proposed(2,i)=unifrnd(-1,1,1,1);                
    para1proposed(3:4,i)=rand(2,1);              
    para1proposed(5,i)=unifrnd(-1,1,1,1);                    
    para1proposed(6:12,i)=rand(7,1);              
    xhat=zeros (1,T);
    x=randi([-3  3],N,1);
    xhat(1)=mean(x);
    loglik=0;
    
for t = 2:T
    
paradox=para1proposed(:,i);
pw(1,1:4)=para1proposed(13:16,i);
mnoise=mvnrnd(Mu(1:3,:),pw(:,1:3));
snoise=mvnrnd(Mu(4,:),pw(:,4));
   % Compute importance weights
   %step3:prediction next_state & 
        x = computenextstate_mytrial4(x,u(t-1,:), paradox,snoise);
        y = my_trial_computey4(x,z_data(t,:), paradox,mnoise);
        %STEP4:Compute Weight
        e1 =repmat(y1_data(t),N,1)-y(:,1);%Prediction Error of y1
        e2 =repmat(y2_data(t),N,1)-y(:,2);%Prediction Error of y2
        e3 =repmat(y3_data(t),N,1)-y(:,3);%Prediction Error of y3
        
        logl1 = -1/2*log(2*pi*(para1proposed(13,i))) - 1/(2*(para1proposed(13,i)))*e1.^2;
        logl2 = -1/2*log(2*pi*(para1proposed(14,i))) - 1/(2*(para1proposed(14,i)))*e2.^2;
        logl3 = -1/2*log(2*pi*(para1proposed(15,i))) - 1/(2*(para1proposed(15,i)))*e3.^2;
       
        logweights=logl1+logl2+logl3; %Weight calculations, log(l1*l2*l3)
        const = max(logweights); % Subtract the maximum value for numerical stability, shift for better calculation
        weights = exp(logweights-const);
        w= weights/sum(weights);  % Save the normalized weights
        % Compute loglikelihood
        loglik= loglik+ const + log(sum(weights)) - log(N);

     %Step5:State estimation
     xhat(t)=w'*x;  %Estimate for state 
     %Step6:Resampling
     index=resampling(w);   %calling resampling function
      x=x((index)',1); %Resampled Particles
    
end %Step 7:End time loopl1
   % Compute the acceptance probability (reject if unstable system)
 %%    
     loglikproposed(i)=loglik;
     acceptprob = eps(exp(loglikproposed(i) - loglikproposed(i-1)));%likelihood contribution
     prior1=eps(prod(unifpdf(para1proposed (:,i), priordispar(:,1),priordispar(:,2))));
     prior0=eps(prod(unifpdf(para1proposed(:,i-1), priordispar(:,1),priordispar(:,2))));
     acceptprob = min(1,acceptprob);
     acceptprobproposed(i)=acceptprob;  
    %Generate random number from uniform distribution
    random = unifrnd(0, 1);
    accept = random < acceptprob;
    acceptrate(i)=accept;
   
   %% Acceptance decision  
    if (accept> 0) %  abs(para1proposed(1:12,i)<1)
    para0proposed(:,i)=para1proposed(:,i);
     xhatfiltered(i,:)=xhat;
    sampl(:,i)=para1proposed(:,i);
    else
     xhatfiltered(i,:)=xhatfiltered(i-1,:);
     para1proposed(:,i)=para1proposed(:,i-1);
     sampl(:,i)=para1proposed(:,i-1);
    end
 %%   
     samplresul=sampl(:,1251:end);  
end
  
    
    %% Taking the average of best samples 
pest=struct('theta1',mean(samplresul(1,:)),'theta2',mean(samplresul(2,:)),'theta3',mean(samplresul(3,:)),'theta4',mean(samplresul(4,:)),'theta5',mean(samplresul(5,:)),'theta6',mean(samplresul(6,:)),'theta7',mean(samplresul(7,1251:end)),'alpha1',mean(samplresul(8,:)),'alpha2',mean(samplresul(9,1251:end)),'alpha3',mean(samplresul(10,:)),'alpha4',mean(samplresul(11,:)),'alpha5',mean(samplresul(12,:)),'w1',mean(samplresul(13,:)),'w2',mean(samplresul(14,:)),'w3',mean(samplresul(15,:)),'w4',mean(samplresul(16,:)));


%Final estimation with estimated parameters
x=randi([-3  3],N,1);
xhat(1)=mean(x);
for t = 2:T
    
paradox(1:12,1)=mean((samplresul(1:12,:)),2);
pw(1,1:4)=mean(samplresul(13:16,2));
mnoise=mvnrnd(Mu(1:3,:),pw(:,1:3));
snoise=mvnrnd(Mu(4,:),pw(:,4));
   % Compute importance weights
   %step3:prediction next_state & 
        x = computenextstate_mytrial4(x,u(t-1,:), paradox,snoise);
        y = my_trial_computey4(x,z_data(t,:), paradox,mnoise);
        %STEP4:Compute Weight
        e1 =repmat(y1_data(t),N,1)-y(:,1);%Prediction Error of y1
        e2 =repmat(y2_data(t),N,1)-y(:,2);%Prediction Error of y2
        e3 =repmat(y3_data(t),N,1)-y(:,3);%Prediction Error of y3
        
        logl1 = -1/2*log(2*pi*(para1proposed(13,i))) - 1/(2*(para1proposed(13,i)))*e1.^2;
        logl2 = -1/2*log(2*pi*(para1proposed(14,i))) - 1/(2*(para1proposed(14,i)))*e2.^2;
        logl3 = -1/2*log(2*pi*(para1proposed(15,i))) - 1/(2*(para1proposed(15,i)))*e3.^2;
       
        logweights=logl1+logl2+logl3; %Weight calculations, log(l1*l2*l3)
        const = max(logweights); % Subtract the maximum value for numerical stability, shift for better calculation
        weights = exp(logweights-const);
        w= weights/sum(weights);  % Save the normalized weights
        % Compute loglikelihood
        loglik= loglik+ const + log(sum(weights)) - log(N);

     %Step5:State estimation
     xhat(t)=w'*x;  %Estimate for state 
     %Step6:Resampling
     index=resampling(w);   %calling resampling function
      x=x((index)',1); %Resampled Particles
    
end 

toc
```
