## Supplementary material for "Quantifying non-communicable diseases’ burden in Egypt using State-Space model": S1 File: acorr.m

```
function c = acorr(x,maxlag)
%ACORR Estimate autocorrelation function of time series
%
%   C = ACORR(X,MAXLAG) returns normalized autocorrelation
%   sequences for each column of X using 
%   C(:,i)=XCORR(X(:,i)-MEAN(X(:,i)),MAXLAG,'coeff'), but returns only
%   lags 1:MAXLAG. Default MAXLAG = M-1;
%
%   See also
%     XCORR

% Copyright (C) 2000 Aki Vehtari
%
% This software is distributed under the GNU General Public 
% Licence (version 2 or later); please refer to the file 
% Licence.txt, included with the software, for details.

if nargin < 1
  error('Not enough input arguments.');
end
if nargin < 2
  maxlag=length(x)-1;
end
[m,n]=size(x);
c=zeros(maxlag,n);
for i1=1:n
  ct=xcorr(x(:,i1)-mean(x(:,i1)),maxlag,'coeff');
  ct=ct(maxlag+2:end);
  c(:,i1)=ct;
end
```
