## Supplementary material for "Quantifying non-communicable diseases’ burden in Egypt using State-Space model": S1 File: acorrtime.m

```
function t = acorrtime(x,maxlag)
% ACORRTIME Estimate autocorrelation evolution of time series
%
%   T = ACORRTIME(X,MAXLAG) returns estimate for autocorrelation 
%   time defined as        MAXLAG
%             t = 1 + 2 sum    r(l),
%                          l=1
%   where r(l) is the sample autocorrelation at the lag l.
%   Sum is cutted of at a finite value L beyond which the
%   autocorrelation estimate is close to zero, since adding
%   autocorrelations for higher lags will only add in excess noise.
%   Autocorrelation sequence is estimated using r=ACORR(X,MAXLAG).
%
%   If MAXLAG is not given, maximum autocorrelation time over all possible
%   MAXLAG values is returned. This serves as a pessimistic estimate.
%
%   See also
%     ACORR

%   References:
%      [1] R. Neal, "Probabilistic Inference Using Markov Chain
%          Monte Carlo Methods", Technical Report CRG-TR-93-1,
%          Dept. of Computer Science, University of Toronto, 1993. p.105.
%      [2] R. E. Kass et al., "Markov chain Monte Carlo in practice:
%          A roundtable discussion", American Statistician,
%          52:93-100, 1998. p. 99.
%      [3] L. Zhu and B. P. Carlin, "Comparing hierarchical models
%          for spatio-temporally misaligned data using DIC
%          criterion. Technical report, Division of Biostatistics,
%          University of Minnesota, 1999. p. 9.

% Copyright (C) 2000-2001 Aki Vehtari
%
% This software is distributed under the GNU General Public 
% Licence (version 2 or later); please refer to the file 
% Licence.txt, included with the software, for details.

if nargin > 1
  t=1+2*sum(acorr(x,maxlag));
else
  t=1+2*max(cumsum(acorr(x)));
end
```
