## Supplementary material for "Quantifying non-communicable diseases’ burden in Egypt using State-Space model": S1 File: Contents.m

```
% MCMC Diagnostics Toolbox for Matlab 6.x
% Version 1.1 2004-01-23
% Copyright (C) 1999 Simo S�rkk�
% Copyright (C) 2000-2004 Aki Vehtari
%
% This software is distributed under the GNU General Public 
% Licence (version 2 or later); please refer to the file 
% Licence.txt, included with the software, for details.
%
% Covergence diagnostics
%   PSRF     - Potential Scale Reduction Factor
%   CPSRF    - Cumulative Potential Scale Reduction Factor
%   MPSRF    - Multivariate Potential Scale Reduction Factor
%   CMPSRF   - Cumulative Multivariate Potential Scale Reduction Factor
%   IPSRF    - Interval-based Potential Scale Reduction Factor
%   CIPSRF   - Cumulative Interval-based Potential Scale Reduction Factor
%   KS       - Kolmogorov-Smirnov goodness-of-fit hypothesis test
%              (requires Statistics toolbox) 
%   HAIR     - Brooks' hairiness convergence diagnostic
%   CUSUM    - Yu-Mykland convergence diagnostic for MCMC
%   SCORE    - Calculate score-function convergence diagnostic
%   GBINIT   - Initial iterations for Gibbs iteration diagnostic
%   GBITER   - Estimate number of additional Gibbs iterations
%
% Time series analysis
%   ACORR      - Estimate autocorrelation function of time series
%   ACORRTIME  - Estimate autocorrelation evolution of time series (simple)
%   GEYER_ICSE - Compute autocorrelation time tau using Geyer's
%                initial convex sequence estimator
%                (requires Optimization toolbox) 
%   GEYER_IMSE - Compute autocorrelation time tau using Geyer's
%                initial monotone sequence estimator
%
% Kernel density estimation etc.:
%   KERNEL1  - 1D Kernel density estimation of data
%   KERNELS  - Kernel density estimation of independent components of data
%   KERNELP  - 1D Kernel density estimation, with automatic kernel width
%   NDHIST   - Normalized histogram of N-dimensional data
%   HPDI     - Estimates the Bayesian HPD intervals
%
% Manipulation of MCMC chains
%   THIN     - Delete burn-in and thin MCMC-chains
%   JOIN     - Join similar structures of arrays to one structure of arrays
%
% Misc:
%   CUSTATS  - Calculate cumulative statistics of data
%
```
