## Supplementary material for "Quantifying non-communicable diseases’ burden in Egypt using State-Space model": S1 File: cpsrf.m

```
function [R,neff,V,W,B] = cpsrf(varargin)
%CPSRF Cumulative Potential Scale Reduction Factor
%
%   [R,neff,V,W,B] = CPSRF(X,[n0]) or
%   [R,neff,V,W,B] = CPSRF(x1,x2,x3,...[,n0])
%   returns Cumulative Potential Scale Reduction Factor for
%   collection of MCMC-simulations. Analysis is first based
%   on PSRF-analysis of samples 1 to n0. Then for samples
%   1 to n0+1 and so on.
%
%   Default value for parameter n0 is |X|/2.
%
%   See also
%     PSRF, MPSRF, CMPSRF

% Copyright (C) 1999 Simo S�rkk�
%
% This software is distributed under the GNU General Public 
% Licence (version 2 or later); please refer to the file 
% Licence.txt, included with the software, for details.

% 2004-01-22 Added neff, R^2->R, and cleaning

% Handle the input arguments
if prod(size(varargin{nargin}))==1
  count = nargin-1;
  n0 = varargin{nargin};
else
  count = nargin;
  n0 = [];
end

if count<=1
  X = varargin{1};
else
  X = zeros([size(varargin{1}) count]);
  for i=1:count
    X(:,:,i) = varargin{i};
  end
end
if isempty(n0)
  n0 = floor(size(X,1)/2) + 1;
end
if n0 < 2
  error('n0 should be at least 2');
end

[N,D,M]=size(X);
R = zeros(N-n0+1,D);
neff = R;
V = R;
W = R;
for i=n0:N
  [R(i-n0+1,:),neff(i-n0+1,:),V(i-n0+1,:),W(i-n0+1,:),B(i-n0+1,:)]=...
      psrf(X(1:i,:,:));
end
```
