## Supplementary material for "Quantifying non-communicable diseases’ burden in Egypt using State-Space model": S1 File: custats.m

```
function [CA,CS] = custats(X,n0)
%CUSTATS Calculate cumulative statistics of data
%
%   [CA,CS] = custats(X) or [CA,CS] = custats(X,n0)
%   returns cumulative statistics of X. CA is the
%   cumulative average and CS is the cumulative variance.

% Copyright (C) 1999 Simo S�rkk�
%
% This software is distributed under the GNU General Public 
% Licence (version 2 or later); please refer to the file 
% Licence.txt, included with the software, for details.


if nargin==1
  n0 = 0;
end

X = X((n0+1):end,:,:);
X2 = X .* X;
CA = cumsum(X)./ repmat((1:size(X,1))',[1 size(X,2) size(X,3)]);
CS = cumsum(X2)./ repmat((1:size(X,1))',[1 size(X,2) size(X,3)]);
CS = CS - CA.*CA;
```
