## Supplementary material for "Quantifying non-communicable diseases’ burden in Egypt using State-Space model": S1 File: cusum.m

```
function C = cusum(W,n0)
%CUSUM Yu-Mykland convergence diagnostic for MCMC
%
%   C = cusum(W,n0) or C = cusum(W) returns
%   cumulative sum of each column of W as:
%
%           n
%          ___
%          \
%   C(i) = /__( W(i) - S ),
%         i=n0+1
%
%   where S is "empirical average":
%            n
%           ___
%           \
%   S = 1/T /__ W(i)
%          i=n0+1
%
%   Default value for "burn-in" variable n0 is 0.
%
%   See also
%     HAIR

% Copyright (C) 1999 Simo S�rkk�
%
% This software is distributed under the GNU General Public 
% Licence (version 2 or later); please refer to the file 
% Licence.txt, included with the software, for details.

if nargin==2
  W = W(n0+1:end,:);
end
T = size(W,1);
S = sum(W) / T;
C = cumsum(W-repmat(S,size(W,1),1));
```
