## Supplementary material for "Quantifying non-communicable diseases’ burden in Egypt using State-Space model": S1 File: gbinit.m

```
function nmin = gbinit(q,r,s)
%GBINIT Initial iterations for Gibbs iteration diagnostic
%
%   nmin = gbinit(q,r,s) returns number of
%   initial iterations needed for estimating how
%   many additional iterations are needed for
%   given precision.
%
%   The definition of the precisions parameters:
%
%   "Suppose that U is function of theta, which is the
%    parameter to be estimated. We want to estimate
%    P[U <= u | y] to within +-r with probability s.
%    We will find the approximate number of iterations
%    needed to do this when the correct answer is q."
%
%    Use q=0.025, r=0.005, s=0.95 if you are unsure.
%
%   See also
%     GBITER

% Copyright (C) 1999 Simo S�rkk�
%
% This software is distributed under the GNU General Public 
% Licence (version 2 or later); please refer to the file 
% Licence.txt, included with the software, for details.

nmin = round(norminv((s+1)/2)^2*q*(1-q)/r^2);
```
