## Supplementary material for "Quantifying non-communicable diseases’ burden in Egypt using State-Space model": S1 File: gbiter.m

```
function [M,N,k] = gbiter(X,q,r,s)
%GBITER Estimate number of additional Gibbs iterations
%
%   [M,N,k] = gbiter(X,[q,r,s]) or returns number of
%   additional iterations required for accurate result
%   and of what portion of the samples should be used.
%   The returned values are as follows:
%
%   M defines how many of the first iterations should be
%   thrown away. N is the number of iterations should be done
%   (after the first M). Only every k'th sample should be used
%
%   X has the initial >Nmin iterations which are
%   used for estimation. Parameters q, r and s
%   are defined as follows:
%
%   "Suppose that U is function of theta, which is the
%    parameter to be estimated. We want to estimate
%    P[U <= u | y] to within +-r with probability s.
%    We will find the approximate number of iterations
%    needed to do this when the correct answer is q."
%
%   Use q=0.025, r=0.005, s=0.95 (defaults) if you are unsure.
%   Thus, it could be reasonable to try different q-values
%   instead of selecting the default 0.025. See Brooks and
%   Roberts (1999) for more detailed discussion.
%
%  References
%    Brooks, S.P. and Roberts, G.O. (1999) On Quantile Estimation
%    and MCMC Convergence. Biometrika.
%
%  See also
%    GBINIT

% Copyright (C) 1999 Simo S�rkk�
%
% This software is distributed under the GNU General Public 
% Licence (version 2 or later); please refer to the file 
% Licence.txt, included with the software, for details.


if nargin < 4
  s = 0.95;
end
if nargin < 3
  r = 0.005;
end
if nargin < 2
  q = 0.025;
end

% Estimate q-quontiles u(:) for each variable and
% create binary sequences Z(:,n).
% (Method is from "gibbsit"-program.)
e = 0.001;
Z = zeros(size(X,1),size(X,2));
for n=1:size(X,2)
  x = sort(X(:,n));
  order = q * (size(X,1)-1) + 1;
  fract = mod(order,1.0);
  low  = max(floor(order),1);
  high = min(low+1, size(X,1));
  u = (1 - fract) * x(low) + fract * x(high);
  Z(:,n) = (X(:,n) <= u);
end

k1 = zeros(1,size(Z,2));
alpha = zeros(1,size(Z,2));
beta  = zeros(1,size(Z,2));
k2 = zeros(1,size(Z,2));

for m=1:size(Z,2)

  % Find out when sequence turns from second
  % order Markov to first order
  bic = 1;
  kthin = 0;
  while bic >= 0
    kthin = kthin + 1;
    z = Z(1:kthin:end,m);

    % Estimate second order Markov chain
    M = zeros(2,2,2);
    for n=3:size(z,1)
      M(z(n-2)+1, z(n-1)+1, z(n)+1) = ...
          M(z(n-2)+1, z(n-1)+1, z(n)+1) + 1;
    end

    % Do g2 test
    count = size(z,1)-2;
    g2 = 0;
    for i=1:2
      for j=1:2
        for k=1:2
          if M(i,j,k) ~= 0
            fitted = sum(M(i,j,:)) * sum(M(:,j,k));
            fitted = fitted / sum(sum(M(:,j,:)));
            g2 = g2 + log( M(i,j,k) / fitted) * M(i,j,k);
          end
        end
      end
    end
    g2 = 2*g2;  
    bic = g2 - 2*log(count);
  end
  k2(m) = kthin;

  % Estimate first order Markov chain
  M = zeros(2,2);
  for n=2:size(z,1)
    M(z(n-1)+1, z(n)+1) = ...
        M(z(n-1)+1, z(n)+1) + 1;
  end
  alpha(m) = M(1,2) / (M(1,1) + M(1,2));
  beta(m) = M(2,1) / (M(2,1) + M(2,2));

  % Find out when sequence turns from first
  % order Markov to independent sequence
  bic = 1;
  kthin = kthin-1;

  while bic >= 0
    kthin = kthin + 1;
    z = Z(1:kthin:end,m);

    % Estimate first order Markov chain
    M = zeros(2,2);
    for n=2:size(z,1)
      M(z(n-1)+1, z(n)+1) = ...
          M(z(n-1)+1, z(n)+1) + 1;
    end

    % Do g2 test
    count = size(z,1)-1;
    g2 = 0;
    for i=1:2
      for j=1:2
        if M(i,j) ~= 0
          fitted = sum(M(i,:)) * sum(M(:,j)) / count;
          g2 = g2 + log( M(i,j) / fitted) * M(i,j);
        end
      end
    end
    g2 = 2*g2;
    bic = g2 - log(count);
  end

  k1(m) = kthin;
end

% Calculate k, N and nburn
tmp = log((alpha + beta) * e ./ ...
          max(alpha,beta)) ./ log(abs(1 - alpha - beta));
nburn = round(tmp) .* k2;

phi = norminv((s+1)/2);
tmp = (2-alpha-beta) .* alpha .* beta ./ (alpha+beta).^3;
tmp = tmp * (phi/r)^2;
N = max(round(max(1,tmp)) .* k2);
k = max(k2);
M = max(nburn);
```
