## Supplementary material for "Quantifying non-communicable diseases’ burden in Egypt using State-Space model": S1 File: geyer_icse.m

```
function [t,t1] = geyer_icse(x,maxlag)
% GEYER_ICSE - Compute autocorrelation time tau using Geyer's
%              initial convex sequence estimator
   
%   References:
%      [1] C. J. Geyer, (1992). "Practical Markov Chain Monte Carlo",
%          Statistical Science, 7(4):473-511

% Copyright (C) 2002 Aki Vehtari
%
% This software is distributed under the GNU General Public 
% Licence (version 2 or later); please refer to the file 
% Licence.txt, included with the software, for details.

% compute autocorrelation
if nargin > 1
  cc=acorr(x,maxlag);
else
  cc=acorr(x);
end
[n,m]=size(cc);

% acorr returns values starting from lag 1, so add lag 0 here
cc=[ones(1,m);cc];
n=n+1;

% now make n even
if mod(n,2)
  n=n-1;
  cc(end,:)=[];
end

% loop through variables
t=zeros(1,m);
t1=zeros(1,m);
opt=optimset('LargeScale','off','display','off');
for i1=1:m
  c=cc(:,i1);
  c=sum(reshape(c,2,n/2),1);
  ci=find(c<0);
  if isempty(ci)
    warning(sprintf('Inital positive could not be found for variable %d, using maxlag value',i1));
    ci=n/2;
  else
    ci=ci(1)-1; % initial positive
  end
  c=[c(1:ci) 0];    % initial positive sequence
  t1(i1)=-1+2*sum(c); % initial positive sequence estimator
  if ci>2
    ca=fmincon(@se,c,[],[],[],[],0*c,c,@sc,opt,c); % monotone convex sequence
  else
    ca=c;
  end
  t(i1)=-1+2*sum(ca); % monotone convex sequence estimator
end

function e = se(x,xx)
% SE - Error in monotone convex sequene estimator
e=sum((xx-x).^2);

function [c,ceq] = sc(x,xx)
% SE - Constraint in monotone convex sequene estimator
ceq=0*x;
c=ceq;
d=diff(x);
dd=-diff(d);
d(d<0)=0;d=d.^2;
dd(dd<0)=0;dd=dd.^2;
c(1:end-1)=d;c(2:end)=c(2:end)+d;
c(1:end-2)=dd;c(2:end-1)=c(2:end-1)+dd;c(3:end)=c(3:end)+dd;
```
