## Supplementary material for "Quantifying non-communicable diseases’ burden in Egypt using State-Space model": S1 File: geyer_imse.m

```
function [t,t1] = geyer_imse(x,maxlag)
% GEYER_IMSE - Compute autocorrelation time tau using Geyer's
%              initial monotone sequence estimator
   
%   References:
%      [1] C. J. Geyer, "Practical Markov Chain Monte Carlo",
%          Statistical Science, 7(4):473-511
%
%   This function is replacment for GEYER_ICSE, when Optimization
%   toolbox is not available
%
%       See also
%       GEYER_ICSE

% Copyright (C) 2002-2003 Aki Vehtari
%
% This software is distributed under the GNU General Public 
% Licence (version 2 or later); please refer to the file 
% Licence.txt, included with the software, for details.


% compute autocorrelation
if nargin > 1
  cc=acorr(x,maxlag);
else
  cc=acorr(x);
end
[n,m]=size(cc);

% acorr returns values starting from lag 1, so add lag 0 here
cc=[ones(1,m);cc];
n=n+1;

% now make n even
if mod(n,2)
  n=n-1;
  cc(end,:)=[];
end

% loop through variables
t=zeros(1,m);
t1=zeros(1,m);
for i1=1:m
  c=cc(:,i1);
  c=sum(reshape(c,2,n/2),1);
  ci=find(c<0);
  if isempty(ci)
    warning(sprintf('Inital positive could not be found for variable %d, using maxlag value',i1));
    ci=n/2;
  else
    ci=ci(1)-1; % initial positive
  end
  c=[c(1:ci) 0];    % initial positive sequence
  t1(i1)=-1+2*sum(c); % initial positive sequence estimator
  if ci>2
    for i2=length(c):-1:2
      if c(i2)>c(i2-1)
        c(i2-1)=c(i2); % monotone sequence
      end
    end
  end
  t(i1)=-1+2*sum(c); % monotone sequence estimator
end
```
