## Supplementary material for "Quantifying non-communicable diseases’ burden in Egypt using State-Space model": S1 File: hair.m

```
function [DA,DS] = hair(W,n0)
%HAIR Brooks' hairiness convergence diagnostic
%
%   [DA,DS] = hair(W,n0) or [DA,DS] = hair(W)
%   returns a measurements of hairiness calculated
%   separately for every column of W
%                n
%                __
%               \
%   DA(t) = 1/t /__ d(i)
%              i=n0+1
%
%                           n
%                           __
%                          \
%   DS(t) = -DA(t)^2 + 1/t /__ d(i)^2
%                          i=n0+1
%
%   where d(i) is 1 if there is local maximum/
%   minimum in CUSUM C(i) of the column of W,
%   0 otherwise. DA is the empirical average and
%   DS variance.
%
%   Default value for "burn-in" variable n0 is 0.
%
%   See also
%     CUSUM

% Copyright (C) 1999 Simo S�rkk�
%
% This software is distributed under the GNU General Public 
% Licence (version 2 or later); please refer to the file 
% Licence.txt, included with the software, for details.

if nargin==2
  C = cusum(W,n0);
else
  C = cusum(W,0);
end
s = sign(C(1:end-1,:) - C(2:end,:));
d = s(1:end-1,:) .* s(2:end,:) < 0;
[DA,DS] = custats(d);
```
