## Supplementary material for "Quantifying non-communicable diseases’ burden in Egypt using State-Space model": S1 File: hpdi.m

```
function hpdi = hpdi(x, p)
% HPDI - Estimates the Bayesian HPD intervals
%
%   Y = HPDI(X,P) returns a Highest Posterior Density (HPD) interval
%   for each column of X. P must be a scalar. Y is a 2 row matrix
%   where ith column is HPDI for ith column of X.

%   References:
%      [1] Chen, M.-H., Shao, Q.-M., and Ibrahim, J. Q., (2000).
%          Monte Carlo Methods in Bayesian Computation. Springer-Verlag.

% Copyright (C) 2001 Aki Vehtari
%
% This software is distributed under the GNU General Public 
% Licence (version 2 or later); please refer to the file 
% Licence.txt, included with the software, for details.

if nargin < 2
  error('Not enough arguments')
end

m=size(x,2);
pts=linspace(0.1,99.9-p,20);
pt1=prctile(x,pts);
pt2=prctile(x,p+pts);
cis=abs(pt2-pt1);
[foo,hpdpi]=min(cis);
if m==1
  hpdi=[pt1(hpdpi); pt2(hpdpi)];
else
  hpdpi=sub2ind(size(pt1),hpdpi,1:m);
  hpdi=[pt1(hpdpi); pt2(hpdpi)];
end
```
