## Supplementary material for "Quantifying non-communicable diseases’ burden in Egypt using State-Space model": S1 File: ipsrf.m

```
function [R] = ipsrf(varargin)
%IPSRF Interavl Potential Scale Reduction Factor
%
%   [R] = IPSRF(X) or
%   [R] = IPSRF(x1,x2,...,xs)
%   returns "Potential Scale Reduction Factor" (PSRF) for
%   collection of MCMC-simulations. X is a NxDxM matrix
%   which contains M MCMC simulations of length N, each with
%   dimension D. MCMC-simulations can be given as separate
%   arguments x1,x2,... which should have the same length.
%
%   Returns 
%     R     PSRF in a row vector of length D
%
%   The idea of the PSRF is that if R is not near 1 (below 1.1 for
%   example) one may conclude that the tested samples were not from
%   the same distribution (chain might not have been converged
%   yet). Instead of normality assumption, 80% empirical intervals
%   are used to compute R.
%
%   If only one simulation is given, the factor is calculated
%   between first and last third of the chain. Note that use of
%   only one chain will produce over-optimistic result.
%
%   Method is from:
%      Brooks, S.P. and Gelman, A. (1998) General methods for
%      monitoring convergence of iterative simulations. Journal of
%      Computational and Graphical Statistics. 7, 434-455. 
%
%   See also
%     CIPSRF, PSRF

% Copyright (C) 1999 Simo S�rkk�
% Copyright (C) 2004 Aki Vehtari
%
% This software is distributed under the GNU General Public 
% Licence (version 2 or later); please refer to the file 
% Licence.txt, included with the software, for details.

% In case of one argument split to two halves (first and last thirds)
onechain=0;
if nargin==1
  X = varargin{1};
  if size(X,3)==1
    n = floor(size(X,1)/3);
    x = zeros([n size(X,2) 2]);
    x(:,:,1) = X(1:n,:);
    x(:,:,2) = X((end-n+1):end,:);
    X = x;
    onechain=1;
  end
elseif nargin==0
  error('Cannot calculate PSRF of scalar');
else
  X = zeros([size(varargin{1}) nargin]);
  for i=1:nargin
    X(:,:,i) = varargin{i};
  end
end

[N,D,M]=size(X);

if N<1
  error('Too few samples');
end

W = zeros(1,D);
V = zeros(1,D);
for d=1:D
  x=X(:,d,:);
  W(1,d)=mean(diff(prctile(x,[10 90])));
  V(1,d)=diff(prctile(x(:),[10 90]));
end
R = V./W;
```
