## Supplementary material for "Quantifying non-communicable diseases’ burden in Egypt using State-Space model": S1 File: join.m

```
function r = join(rs, joinall)
%JOIN Join similar structures of arrays to one structure of arrays
%
%   JOIN can be used to combine statistics collected from several
%   independent simulations which collect samples to structure
%   of arrays.
%
%   R = JOIN(RS) returns structure of arrays R, RS is array or
%   cell-array of similar structures of arrays. Arrays of R are
%   contanation of arrays in RS. Arrays having first dimension 1
%   are not concataned, but copied from the first element of RS.
%
%   R = JOIN(RS, JOINALL) if JOINALL is 1, arrays having first
%   dimension 1 are concataned.
%
%   See also
%     THIN

% Copyright (C) 2000-2003  Aki Vehtari
%
% This software is distributed under the GNU General Public 
% Licence (version 2 or later); please refer to the file 
% Licence.txt, included with the software, for details.

if nargin <2
  joinall=false;
end

[m,n]=size(rs);

r=[];
if (m==1 & n==1)
  r=rs;
else
  if iscell(rs)
    rs=[rs{:}];
  end
  if isstruct(rs)
    r=rs(1);
    names = fieldnames(r);
    for i1=1:size(names,1)
      tmp1=getfield(r,names{i1});
      if iscell(tmp1)
	tmp2=eval(['cat(1,rs(:).' names{i1} ')']);
	for i2=1:length(tmp1(:))
	  eval(['r.' names{i1} '{i2}=cat(1,tmp2{:,i2});']);
	end
      elseif length(tmp1) > 1 || joinall
	eval(['r.' names{i1} '=cat(1,rs.' names{i1} ');']);
      end
    end
  end
end
```
