## Supplementary material for "Quantifying non-communicable diseases’ burden in Egypt using State-Space model": S1 File: kernel1.m

```
function [P,X,sigma] = kernel1(T,sigma,bins,extra)
%KERNEL1 1D Kernel density estimation of data
%
%   [P,X,sigma] = kernel1(T,sigma,bins,extra) returns kernel based
%   marginal density estimate of each column of T. Default value
%   for the number of bins is min{50,sqrt(|T|)}. Default value
%   for the standard deviation sigma is max(STD(T)/2). Default
%   value for fraction of empty extra space is 0.2 (that is 20%).
%
%   If no output arguments is given, functions plots the
%   graphs of each density component. Otherwise smoothed
%   and normalized densities are returned in P and the
%   corresponding coordinates in X.
%
%   See also
%     NDHIST

% Copyright (C) 1999 Simo S�rkk�
%
% This software is distributed under the GNU General Public 
% Licence (version 2 or later); please refer to the file 
% Licence.txt, included with the software, for details.

% 2000-03-27  Aki Vehtari  <>
%             Get size of T only once and check if T is horizontal vector.
% 2002-08-09  Aki Vehtari  <>
%             Make sure P >= 0
% 2003-11-04  Aki Vehtari  <>
%             Define normpdf because not everyone has Statistics toolbox

  [s1,s2]=size(T);
  if s1==1
    T=T(:);
    [s1,s2]=size(T);
  end

  if nargin < 2
    sigma = [];
  end
  if nargin < 3
    bins = [];
  end
  if nargin < 4
    extra = [];
  end

  if isempty(sigma)
    sigma = std(T)/2;
  else
    sigma = sigma(:)';
  end
  if isempty(bins)
    bins = max(50,floor(sqrt(s1))+1);
  end
  if isempty(extra)
    extra = 0.2;
  end
  if size(sigma,2)~=s2
    sigma = ones(1,s2)*sigma;
  end

  if s2 > 1
    P = zeros(bins,s2);
    X = zeros(bins,s2);
    for i=1:s2
      [P(:,i),X(:,i),sigma(i)] = kernel1(T(:,i),sigma(i),bins,extra);
    end
  else
    mx = max(T);
    mn = min(T);
    delta = extra * (mx - mn);
    mn = mn - delta/2;
    mx = mx + delta/2;
    dx = (mx - mn) / bins;
    [H,X] = ndhist(T,bins,mn,mx);
    H = H(:) / sum(H);
    x = 2*(max(X)-min(X))*(0:(2*size(H,1)-1))'/size(H,1);
    G = normpdf(x,mean(x),sigma);
    G = ifftshift(G(:));
    P = real(ifft(fft(G,size(G,1)) .* fft(H,size(G,1))));
    P = P(1:size(H,1));
    P = P / (sum(P) * dx);
    P(P<0)=0; % Make sure P >= 0
  end

  if nargout == 0
    m = 2;
    n = ceil(size(X,2)/m);
    while m*m < n
      m = m + 1;
      n = ceil(size(X,2)/m);
    end
    if s2 > 1
      for i=1:s2
	subplot(m,n,i);
	plot(X(:,i),P(:,i));
	set(gca,'XLim',minmax(X(:,i)'));
	ylim = get(gca,'YLim');
	set(gca,'YLim',[0 ylim(2)]);
	title(['T_' num2str(i)]);
      end
    else
      plot(X,P);
    end
    clear X;
    clear P;
    clear sigma;
  end

function y = normpdf(x,mu,sigma)
y = -0.5 * ((x-mu)./sigma).^2 -log(sigma) -log(2*pi)/2;
y=exp(y);
```
