## Supplementary material for "Quantifying non-communicable diseases’ burden in Egypt using State-Space model": S1 File: kernelp.m

```
function [p,xx]=kernelp(x,xx)
%KERNELP 1D Kernel density estimation of data, with automatic kernel width
%
%  [P,XX]=KERNELP(X,XX) return density estimates P in points XX,
%  given data and optionally ecvaluation points XX. Density
%  estimate is based on simple Gaussian kernel density estimate
%  where all kernels have equal width and this width is selected by
%  optimising plug-in partial predictive density. Works well with
%  reasonable sized X.
%

% Copyright (C) 2001-2003 Aki Vehtari
%
% This software is distributed under the GNU General Public 
% Licence (version 2 or later); please refer to the file 
% Licence.txt, included with the software, for details.

if nargin < 1
  error('Too few arguments');
end
[n,m]=size(x);
if n>1 && m>1
  error('X must be a vector');
end
x=x(:);

if nargin < 2
  n=200;
  xa=min(x);xb=max(x);xd=xb-xa;
  xa=xa-xd/20;xb=xb+xd/20;
  xx=linspace(xa,xb,n);
else
  [n,m]=size(xx);
  if n>1 && m>1
    error('XX must be a vector');
  end
  xx=xx(:);
end
m=length(x)/2;
stdx=std(x);
xd=gminus(x(1:m),x(m+1:end)');
sh=fminbnd(@err,stdx/5,stdx*20,[],xd);
p=mean(normpdf(gminus(x(1:m),xx),0,sh));

function e=err(s,xd)
e=-sum(log(sum(normpdf(xd,0,s))));

function y = normpdf(x,mu,sigma)
y = -0.5 * ((x-mu)./sigma).^2 -log(sigma) -log(2*pi)/2;
y=exp(y);

function y=gminus(x1,x2)
%GMINUS   Generalized minus.
y=genop(@minus,x1,x2);

function y=genop(f,x1,x2)
% GENOP - Generalized operation
%   
%   C = GENOP(F,A,B) Call function F with exapanded matrices Y and X.
%   The dimensions of the two operands are compared and singleton
%   dimensions in one are copied to match the size of the other.
%   Returns a matrix having dimension lengths equal to
%   MAX(SIZE(A),SIZE(B))
%
% See also GENOPS

% Copyright (C) 2003 Aki Vehtari
%
% This software is distributed under the GNU General Public 
% Licence (version 2 or later); please refer to the file 
% Licence.txt, included with the software, for details.

s1=size(x1);
s2=size(x2);
ls1=numel(s1);
ls2=numel(s2);
l=max(ls1,ls2);
d=ls1-ls2;
if d<0
  s1(ls1+1:ls1+d)=1;
elseif d>0
  s2(ls2+1:ls2+d)=1;
end
if any(s1>1 & s2>1 & s1~=s2)
  error('Array dimensions are not appropriate.');
end
r1=ones(1,l);
r2=r1;
r1(s1==1)=s2(s1==1);
r2(s2==1)=s1(s2==1);
y=feval(f,repmat(x1,r1),repmat(x2,r2));
```
