## Supplementary material for "Quantifying non-communicable diseases’ burden in Egypt using State-Space model": S1 File: kernels.m

```
function [P,X,V,D] = kernels(T,varargin)
%KERNELS Kernel density estimation of principal components of data
%
%   [P,X,V,D] = kernels(T,[sigma,bins]) returns kernel based
%   marginal density estimate of each pricipal component of T.
%   Default value for the number of bins is min{50,sqrt(|T|)}.
%   Default value for the standard deviation sigma is the STD(T*)/2
%   where T* is the independent component of T.
%
%   If no output arguments is given, functions plots the
%   graphs of each density component. Otherwise densities are
%   returned in P, the corresponding coordinates in X, directions
%   of principal components in V and variances of the principal
%   components in diagonal of D.
%
%   Returned pricipal components are the uncorrelated (but not
%   independent) directions of the density. Relationship between
%   T and coordinates in X is X = T*V.
%
%   See also
%     KERNEL1

% Copyright (C) 1999 Simo S�rkk�
%
% This software is distributed under the GNU General Public 
% Licence (version 2 or later); please refer to the file 
% Licence.txt, included with the software, for details.


m = mean(T);
[V,D] = eig(cov(T));
[C,A,W] = fastica((T-repmat(m,size(T,1),1))','verbose','off',...
                  'displayMode','off');
V = A';
V = V ./ repmat(sqrt(sum(V.*V)),size(V,1),1);
[P,X] = kernel1(C'+repmat(m,size(T,1),1),varargin{:});
if nargout == 0
  m = 2;
  n = ceil(size(X,2)/m);
  while m*m < n
    m = m + 1;
    n = ceil(size(X,2)/m);
  end
  for i=1:size(X,2)
    subplot(m,n,i);
    plot(X(:,i),P(:,i));
    title(['T^*_' num2str(i) ' = ' '[ ' num2str(V(:,i)',' %.2f') ' ]']);
  end
end
```
