## Supplementary material for "Quantifying non-communicable diseases’ burden in Egypt using State-Space model": S1 File: ks.m

```
function P = ks(varargin)
%PSRF Kolmogorov-Smirnov goodness-of-fit hypothesis test.
%
%   P = KS(X) or
%   P = KS(X1,X2,...,XN)
%   returns p-value(s) for Kolmogorov-Smirnov goodness-of-fit
%   hypothesis test for two MCMC-simulations.  X is a NxMx2 
%   matrix which contains 2 MCMC simulations of length N,
%   each with dimension M. MCMC-simulations can be given as separate
%   arguments X1,X2,... which should have the same length.
%
%   Function returns p-values in row vector R of length N.
%
%   If only one simulation is given, the factor is calculated
%   between first and last third of the chain. Note that use of
%   only one chain will produce over-optimistic result.
%
%   See also
%     KSTEST2

% Copyright (C) 2001 Aki Vehtari
%
% This software is distributed under the GNU General Public 
% Licence (version 2 or later); please refer to the file 
% Licence.txt, included with the software, for details.


% In case of one argument split to two halves (first and last thirds)
if nargin==1
  X = varargin{1};
  if size(X,3)==1
    n = floor(size(X,1)/3);
    x = zeros([n size(X,2) 2]);
    x(:,:,1) = X(1:n,:);
    x(:,:,2) = X((end-n+1):end,:);
    X = x;
  elseif size(X,3)~=2
    error('S must be 1 or 2');
  end
else
  X = zeros([size(varargin{1}) nargin]);
  for i=1:nargin
    X(:,:,i) = varargin{i};
  end
end

if (size(X,1)<1)
  error('Too few samples');
end
  
% Calculate means W of the variances
P = zeros(1,size(X,2));
for i1=1:size(X,2)
  [h,P(i1)]=kstest2(X(:,i1,1),X(:,i1,2));
end
```
