## Supplementary material for "Quantifying non-communicable diseases’ burden in Egypt using State-Space model": S1 File: mpsrf.m

```
function [R,neff,V,W,B] = mpsrf(varargin)
%MPSRF Multivariate Potential Scale Reduction Factor
%
%   [R,neff,V,W,B] = MPSRF(X) or
%   [R,neff,V,W,B] = MPSRF(x1,x2,...,xs)
%   returns "Multivariate Potential Scale Reduction Factor"
%   (MPSRF) for collection of MCMC-simulations. X is a NxMxS
%   matrix which contains S MCMC simulations of length M with
%   dimension M. MCMC-simulations can be given as separate
%   arguments x1,x2,... which should have the same length.
%
%   Returns 
%     R     PSRF (R=sqrt(V/W)) in row vector of length D
%     neff  estimated effective number of samples mean(diag(M*N*V./B))
%     V     estimated mixture-of-sequences cvariance
%     W     estimated within sequence covariance
%     B     estimated between sequence covariance
%
%   If only one simulation is given, the factor is calculated
%   between first and last third of the chain. Note that use of
%   only one chain will produce over-optimistic result.
%
%   Method is from:
%   Method is from:
%      Brooks, S.P. and Gelman, A. (1998) General methods for
%      monitoring convergence of iterative simulations. Journal of
%      Computational and Graphical Statistics. 7, 434-455. Note that
%      this function returns square-root definiton of R (see Gelman
%      et al (2003), Bayesian Data Analsyis p. 297).
%
%   See also
%     CMPSRF, PSRF, CPSRF

% Copyright (C) 1999 Simo S�rkk�
%
% This software is distributed under the GNU General Public 
% Licence (version 2 or later); please refer to the file 
% Licence.txt, included with the software, for details.

% 2004-01-22 Added neff, R^2->R, and cleaning


% In case of one argument split to two halves
onechain=0;
if nargin==1
  X = varargin{1};
  if size(X,3)==1
    n = floor(size(X,1)/3);
    x = zeros([n size(X,2) 2]);
    x(:,:,1) = X(1:n,:);
    x(:,:,2) = X((end-n+1):end,:);
    X = x;
    onechain=1;
  end
elseif nargin==0
  error('Cannot calculate PSRF of scalar');
else
  X = zeros([size(varargin{1}) nargin]);
  for i=1:nargin
    X(:,:,i) = varargin{i};
  end
end

[N,D,M]=size(X);

% Calculate mean W of the covariances
W = zeros(D);
for n=1:M
  x = X(:,:,n) - repmat(mean(X(:,:,n)),N,1);
  W = W + x'*x;
end
W = W / ((N-1) * M);

% Calculate covariance B (in fact B/n) of the means.
Bpn = zeros(D);
m = mean(reshape(mean(X),D,M)');
for n=1:M
  x = mean(X(:,:,n)) - m;
  Bpn = Bpn + x'*x;
end
Bpn = Bpn / (M-1);

% Calculate reduction factor R
E = sort(abs(eig(W \ Bpn)));
R = (N-1)/N + E(end) * (M+1)/M;
V = (N-1) / N * W + (1 + 1/M) * Bpn;
R = sqrt(R);  
B = Bpn*N;
neff = mean(min(diag(M*N*V./B),M*N));
if onechain & (nargout>1)
  neff=neff*3/2;
end
```
