## Supplementary material for "Quantifying non-communicable diseases’ burden in Egypt using State-Space model": S1 File: ndhist.m

```
function [H,varargout] = ndhist(X,bins,mins,maxs,cdim)
%NDHIST Normalized histogram of N-dimensional data
%
%   [H,P] = ndhist(X,bins,[mins,maxs])
%   [H,x1,x2,...] = ndhist(X,bins,[mins,maxs])
%
%   Returns normalized N-dimensional histogram H and the bin center
%   coordinates in P or variables x1,x2,... which all are of the
%   same size as H and contain the coordinates in same form as
%   output of ndgrid.
%
%   Histogram is calculated for the points in argument X..
%   bins is a vector containing number of bins for each dimension.
%   Optional arguments mins and maxx specify the maximum allowed
%   values for each dimension.
%
%   If no output arguments are given the function plots
%   graph of the histogram.
%
%   Examples:
%   >> X = gamrnd(3,3,1000,1);
%   >> [H,x] = ndhist(X,50);
%   >> bar(x,H);
%
%   >> X = mvnrnd([1 1],eye(2),1000) + round(rand(1000,1))*[-2 -2];
%   >> [H,x,y] = ndhist(X,[20 20]);
%   >> surf(x,y,H);
%
%   See also
%     KERNEL1, HIST, NDGRID

% Copyright (C) 1999 Simo S�rkk�
%
% This software is distributed under the GNU General Public 
% Licence (version 2 or later); please refer to the file 
% Licence.txt, included with the software, for details.


% Check input and output arguments
if nargin < 2
  bins = [];
end
if nargin < 3
  mins = [];
end
if nargin < 4
  maxs = [];
end
if nargin < 5
  cdim = 1;
  if size(X,1)==1
    X = X';
  end
end

if isempty(bins)
  bins = 10;
end
if isempty(mins)
  mins = min(X);
end
if isempty(maxs)
  maxs = max(X);
end

tmp = (maxs - mins) ./ bins;
mins = mins - tmp/2;
maxs = maxs + tmp/2;
dx = (maxs - mins) ./ bins;

if (nargout~=0) & (nargout~=1) & (nargout~=2) & (nargout~=size(X,2)+1)
  error('Illegal number of output arguments');
end

bins = reshape(bins,1,prod(size(bins)));
if size(bins,2)~=size(X,2)
  if size(bins,2)==1
    bins = bins * ones(1,size(X,2));
  else
    error('Wrong number of bin sizes');
  end
end

% Create histogram for this dimension and
% recursively for each sub-dimension.
[tmpx,tmpi] = sort(X(:,cdim));
X = X(tmpi,:);
if cdim < size(X,2)
  H = zeros(bins(cdim),prod(bins(cdim+1:end)));
else
  H = zeros(bins(cdim),1);
end
bnd = mins(cdim)+dx(cdim);
beg = 1;
ind = 1;
k = 1;

while (k <= size(X,1)) & (X(k,cdim) < mins(cdim))
  k = k + 1;
end
beg = k;
while (k <= size(X,1)) & (X(k,cdim) <= maxs(cdim)) & (ind <= size(H,1))
  if X(k,cdim) <= bnd
    k = k + 1;
  else
    if k-beg > 0
      if cdim < size(X,2)
        H(ind,:) = ndhist(X(beg:k-1,:),bins,mins,maxs,cdim+1);
      else
        H(ind) = k-beg;
      end
    end
    bnd = bnd + dx(cdim);
    ind = ind + 1;
    beg = k;
  end
end

% Handle the last bin
if (ind <= size(H,1))
  if cdim < size(X,2)
    H(ind,:) = ndhist(X(beg:k-1,:),bins,mins,maxs,cdim+1);
  else
    H(ind) = k-beg;
  end
end

% Reshape H to linear or multidimensional and
% determine point positions and set output variables
if cdim == 1
  if size(X,2)==1
    H = H(:);
    x = (0:(bins-1))'*(maxs-mins)/bins+mins+dx/2;
    varargout{1} = x;
  else
    H = reshape(H,bins);
    k = 0;
    tmp = cell(size(X,2),1);
    for i=1:size(X,2)
      x = (0:(bins(i)-1))'/bins(i)*(maxs(i)-mins(i))+mins(i)+dx(i)/2;
      args = cell(size(X,2),1);
      for j=1:size(X,2)
        args{j} = x;
      end
      tmp{i} = shiftdim(ndgrid(args{:}),k);
      k = mod(k + size(X,2)-1,size(X,2));
    end
    if nargout==2
      P = zeros(size(X,2),prod(bins));
      for i=1:size(X,2)
        P(i,:) = reshape(tmp{i},1,prod(bins));
      end
      varargout{1} = reshape(P,[size(X,2) bins]);
    else
      varargout = tmp;
    end
  end
  c = sum(reshape(H,1,prod(bins)));
  H = H/c;
else
  H = reshape(H,1,prod(bins(cdim:end)));
end

% If there were no output arguments, draw a graph
if nargout==0
  if size(X,2)==1
    bar(varargout{1},H,'hist');
  elseif size(X,2)==2
    surf(varargout{1},varargout{2},H);
  else
    error('Unable to draw >2 dimensional densities');
  end
  clear H;
  clear varargout;
end
```
