## Supplementary material for "Quantifying non-communicable diseases’ burden in Egypt using State-Space model": S1 File: psrf.m

```
function [R,neff,V,W,B] = psrf(varargin)
%PSRF Potential Scale Reduction Factor
%
%   [R,NEFF,V,W,B] = PSRF(X) or
%   [R,NEFF,V,W,B] = PSRF(x1,x2,...,xs)
%   returns "Potential Scale Reduction Factor" (PSRF) for
%   collection of MCMC-simulations. X is a NxDxM matrix
%   which contains M MCMC simulations of length N, each with
%   dimension D. MCMC-simulations can be given as separate
%   arguments x1,x2,... which should have the same length.
%
%   Returns 
%     R     PSRF in a row vector of length D
%     neff  estimated effective number of samples M*N*V/B
%     V     estimated mixture-of-sequences variances
%     W     estimated within sequence variances
%     B     estimated between sequence variances
%
%   The idea of the PSRF is that if R is not near 1 (below 1.1 for
%   example) one may conclude that the tested samples were not from
%   the same distribution (chain might not have been converged
%   yet).
%
%   If only one simulation is given, the factor is calculated
%   between first and last third of the chain. Note that use of
%   only one chain will produce over-optimistic result.
%
%   Method is from:
%      Brooks, S.P. and Gelman, A. (1998) General methods for
%      monitoring convergence of iterative simulations. Journal of
%      Computational and Graphical Statistics. 7, 434-455. Note that
%      this function returns square-root definiton of R (see Gelman
%      et al (2003), Bayesian Data Analsyis p. 297).
%
%   See also
%     CPSRF, MPSRF, IPSRF

% Copyright (C) 1999 Simo S�rkk�
% Copyright (C) 2003 Aki Vehtari
%
% This software is distributed under the GNU General Public 
% Licence (version 2 or later); please refer to the file 
% Licence.txt, included with the software, for details.

% 2004-01-22 Added neff, R^2->R, and cleaning

% In case of one argument split to two halves (first and last thirds)
onechain=0;
if nargin==1
  X = varargin{1};
  if size(X,3)==1
    n = floor(size(X,1)/3);
    x = zeros([n size(X,2) 2]);
    x(:,:,1) = X(1:n,:);
    x(:,:,2) = X((end-n+1):end,:);
    X = x;
    onechain=1;
  end
elseif nargin==0
  error('Cannot calculate PSRF of scalar');
else
  X = zeros([size(varargin{1}) nargin]);
  for i=1:nargin
    X(:,:,i) = varargin{i};
  end
end

[N,D,M]=size(X);

if N<1
  error('Too few samples');
end

% Calculate means W of the variances
W = zeros(1,D);
for n=1:M
  x = X(:,:,n) - repmat(mean(X(:,:,n)),N,1);
  W = W + sum(x.*x);
end
W = W / ((N-1) * M);

% Calculate variances B (in fact B/n) of the means.
Bpn = zeros(1,D);
m = mean(reshape(mean(X),D,M)');
for n=1:M
  x = mean(X(:,:,n)) - m;
  Bpn = Bpn + x.*x;
end
Bpn = Bpn / (M-1);

% Calculate reduction factors
S = (N-1)/N * W + Bpn;
R = (M+1)/M * S ./ W - (N-1)/M/N;
V = R .* W;
R = sqrt(R);  
B = Bpn*N;
neff = min(M*N*V./B,M*N);
if onechain & (nargout>1)
  neff=neff*3/2;
end
```
