## Supplementary material for "Quantifying non-communicable diseases’ burden in Egypt using State-Space model": S1 File: Readme.txt

MCMC Diagnostics Toolbox for Matlab 6.x

Copyright (C) 1999 Simo Särkkä <>
Copyright (C) 2000-2004 Aki Vehtari <>
Maintainer: Aki Vehtari <>

In 1999 Simo Särkkä implemented several MCMC diagnostics in Matlab
at Helsinki University of Technology, Laboratory of Computational
Engineering <http://www.lce.hut.fi>. Later Aki Vehtari added a few
additonal utilities, fixed bugs and improved the documentation.

This software is distributed under the GNU General Public Licence
(version 2 or later); please refer to the file Licence.txt,
included with the software, for details.
