## Supplementary material for "Quantifying non-communicable diseases’ burden in Egypt using State-Space model": S1 File: score.m

```
function [CA,CS] = score(X,gradF,n0,varargin)
%SCORE Calculate score-function convergence diagnostic
%
%   [CA,CS] = score(X,gradF,n0,P1,P2,...) 
%   returns convergence diagnostic based on score-functions
%   d ln(p(x))/dx_k that should should approach 0 as n increases.
%
%   gradF is name of the gradient logarithm function (or
%   its opposite) and X's are the sampled values. n0 is length
%   of "burn-in" and defaults to 0.
%
%   The idea is from:
%     Anne Philippe and Christian P. Robert (Oct, 1998)
%     Riemann Sums for MCMC Estimation and
%     Convergence Monitoring. EP CNRS

% Copyright (C) 1999 Simo S�rkk�
%
% This software is distributed under the GNU General Public 
% Licence (version 2 or later); please refer to the file 
% Licence.txt, included with the software, for details.

if (nargin < 3) | isempty(n0)
  n0 = 0;
end
X = X((n0+1):end,:,:);
G = zeros(size(X));
for i=1:size(X,1)
  for j=1:size(X,3)
    G(i,:,j) = feval(gradF,X(i,:,j),varargin{:});
  end
end

[CA,CS] = custats(G);
```
