## Supplementary material for "Quantifying non-communicable diseases’ burden in Egypt using State-Space model": S1 File: thin.m

```
function x = thin(x,nburn,nthin,nlast)
% THIN Delete burn-in and thin MCMC-chains
%
%   X = THIN(X,NBURN,NTHIN,NLAST) returns chain containing only
%   every NTHIN:th simulation sample starting from sample number
%   NBURN+1 and continuing to sample number NLAST.
%
%   See also
%     JOIN

% Copyright (C) 1999 Simo S�rkk�
% Copyright (C) 2000 Aki Vehtari
%
% This software is distributed under the GNU General Public 
% Licence (version 2 or later); please refer to the file 
% Licence.txt, included with the software, for details.

if nargin < 4
  nlast = [];
end
if nargin < 3
  nthin = [];
end
if nargin < 2
  nburn = [];
end

[m,n]=size(x);

if isfield(x,'rstate')
  x=rmfield(x,'rstate');
end

if isstruct(x)
  if (m>1 | n>1)
    % array of structures
    for i=1:(m*n)
      x(i) = thin(x(i),nburn,nthin,nlast);
    end
  else
    % single structure
    names = fieldnames(x);
    for i=1:size(names,1)
      value = getfield(x,names{i});
      if length(value) > 1
	x = setfield(x,names{i},thin(value,nburn,nthin,nlast));
      elseif iscell(value)
	x = setfield(x,names{i},{thin(value{1},nburn,nthin,nlast)});
      end
    end
  end
elseif iscell(x)
  % cell array
  for i=1:(m*n)
    x{i} = thin(x{i},nburn,nthin,nlast);
  end
elseif m > 1
  % field array
  if isempty(nburn)
    nburn = 0;
  elseif (nburn < 0) | (nburn >= m)
    error('Illegal burn-in value');
  end
  if isempty(nthin)
    nthin = 1;
  elseif (nthin < 1) | (nthin > m)
    error('Illegal thinning value');
  end
  if isempty(nlast)
    nlast = m;
  elseif (nlast < 1) | (nlast > m)
    error('Illegal last index');
  end
  x = x((nburn+1):nthin:nlast,:);
end
```
