## Supplementary material for "Quantifying non-communicable diseases’ burden in Egypt using State-Space model": S1 File: acf.m

```
function y=acf(x,lagmax)
%ACF Autocorrelation function
% ACF(X,maxlag)
% default maxlag is floor(10*log10(length(x)))

% There is also acf.mex version which is much faster
% it is used if Matlab finds it

% Marko Laine <>
% $Revision: 1.5 $  $Date: 2012/09/27 11:47:33 $

x = x(:)'-mean(x);
n = length(x);
if nargin<2
  lagmax = floor(10*log10(n));
  lagmax = min(lagmax, n-1);
end
y = filter(x(n:-1:1),1,x);
%y  = conv(flipud(x),x);
y = y(n:-1:1)/n;
y = y/y(1);
y = y(1:lagmax+1);
```
