## Supplementary material for "Quantifying non-communicable diseases’ burden in Egypt using State-Space model": S1 File: addbin.m

```
function status=addbin(filename,x)
%ADDBIN add columns to a V4 matfile
% Usage: addbin(filename,x)
% Adds columns of x to data in mat file. Columns are added because
% MATLAB stores data columnwise. Transpose the data to add rows.
% The file should contain only one double precision matrix.
% This is used in MCMCRUN to save and append temporary mcmc
% chains into binary files.
% See also SAVEBIN and READBIN.

% $Revision: 1.3 $  $Date: 2010/12/10 10:47:38 $

m  = size(x,1);
n  = size(x,2);

fid = fopen(filename,'r+b');
if fid == -1 
  error(sprintf('error opening binary file %s',filename));
end

if fseek(fid,4,'bof') == -1
  fclose(fid);
  error('error seeking file');
end

mrows   = fread(fid,1,'integer*4');
mcols   = fread(fid,1,'integer*4');
imagf   = fread(fid,1,'integer*4');
namelen = fread(fid,1,'integer*4');

if mrows > 0 & mrows ~= m
  error('x should have same number of rows');
end

% goto begining of x
if fseek(fid,namelen,'cof') == -1
  fclose(fid);
  error('error seeking file');
end

if fseek(fid,0,'eof') == -1
  fclose(fid);
  error('error seeking eof');
end

% write the new data to file
fwrite(fid,x,'real*8');

% fix mrows
if mrows == 0
  if fseek(fid,4,'bof') == -1
    fclose(fid);
    error('error seeking file');
  end
  fwrite(fid,m,'integer*4');
end

% write the new column size
if fseek(fid,8,'bof') == -1
  fclose(fid);
  error('error seeking file');
end
fwrite(fid,mcols+n,'integer*4');

fclose(fid);

if nargout > 0
  status = 1;
end
```
