## Supplementary material for "Quantifying non-communicable diseases’ burden in Egypt using State-Space model": S1 File: arimagen.m

```
function y=arimagen(ar,ma,n,s)
% ARIMAGEN - ARMA time series generation
% ARIMAGEN(ar,ma,n,s), ar, ma vectors of AR and MA coefficients, n
% length of the series, s innovation std


% Marko Laine <>
% $Revision: 1.4 $  $Date: 2012/09/27 11:47:33 $

if nargin<4;s=1;end

ar=ar(:);
ma=ma(:);
p=length(ar);
q=length(ma);

% y = filter([1, -ma],[1, -ar],randn(n+p,1).*s);
% y = y(p+1:n+p);

wmean=0;

const=wmean*(1-sum(ar));

a = randn(n+q,1).*s;
x = zeros(n+p,1);

for i=1:n
   x(p+i) = const + sum(ar.*x(i:p+i-1)) + a(i+q) ...
      - sum(ma.*a(i:q+i-1));
end

y=x(p+1:n+p);
```
