## Supplementary material for "Quantifying non-communicable diseases’ burden in Egypt using State-Space model": S1 File: assifun.m

```
function y=assifun(x,a,varargin)
%ASSIFUN assing value to a element in a matrix and return the new matrix
% y=assifun(x,a,i,j,...)
% Useful mainly in inline functions.

%, 2013
% $Revision: 1.1 $  $Date: 2013/01/31 12:06:26 $

y = x;
y(varargin{:}) = a;
```
