## Supplementary material for "Quantifying non-communicable diseases’ burden in Egypt using State-Space model": S1 File: betabinpf.m

```
function y=betabinpf(x,n,a,b)
% BETABINPF Beta Binomial probability function
% BETABINPF(x,n,a,b)

% Mean n*a/(a+b)
% Var  n*a*b(n+a+b)/(a+b)^2/(1+a+b)

% Marko Laine <>
% $Revision: 1.3 $  $Date: 2015/08/07 11:07:11 $
% y = binom(n,x).*beta(x+a,n-x+b)./beta(a,b);
% log version
y = exp(gammaln(n+1)-gammaln(x+1)-gammaln(n-x+1)+betaln(x+a,n-x+b)-betaln(a,b));
```
