## Supplementary material for "Quantifying non-communicable diseases’ burden in Egypt using State-Space model": S1 File: betabinr.m

```
function r=betabinr(m,n,nn,a,b)
%BETABINR  Beta Binomial random numbers
% BETABINR(mr,nr,n,a,b) mr,nr shape of the result, a,b beta parameters

% Marko Laine <>
% $Revision: 1.2 $  $Date: 2012/09/27 11:47:33 $
p = betar(m,n,a,b);
r = zeros(m,n);
for i=1:m*n
  r(i) = binr(1,1,nn,p(i));
end
```
