## Supplementary material for "Quantifying non-communicable diseases’ burden in Egypt using State-Space model": S1 File: betapf.m

```
function y=betapf(x,a,b)
%BETAPF  Beta probability density function
% y = betapf(x,a,b) returns G(a+b)/G(a)/G(b)*x^(a-1)*(1-x)^(b-1)
% where G is the gamma function

% Marko Laine <>
% $Revision: 1.5 $  $Date: 2012/09/27 11:47:33 $
%y = gamma(a+b)/gamma(a)/gamma(b)*x.^(a-1).*(1-x).^(b-1);
y = x.^(a-1).*(1-x).^(b-1)./beta(a,b);
```
