## Supplementary material for "Quantifying non-communicable diseases’ burden in Egypt using State-Space model": S1 File: betaqf.m

```
function y=betaqf(p,a,b)
%BETAQF  Beta inverse cumulative density function
% BETAQF(p,a,b)

% Marko Laine <>
% $Revision: 1.3 $  $Date: 2012/09/27 11:47:33 $

if any(p>=1|p<=0)
  error('argument p must be in open interval (0,1)')
end

% does not work if p=0 or p=1
y = zeros(size(p));
% find zero by bisection
zfun = @(x,a,b,p) betainc(x,a,b)-p;
for i=1:prod(size(y))
  y(i) = bisect(zfun,0,1,optimset('TolX',1e-6),a,b,p(i));
end
```
