## Supplementary material for "Quantifying non-communicable diseases’ burden in Egypt using State-Space model": S1 File: betar.m

```
function r=betar(m,n,a,b)
%BETAR  Beta random numbers
% BETAR(m,n,a,b) m,n shape of the result, a,b beta parameters

% Marko Laine <>
% $Revision: 1.5 $  $Date: 2012/09/27 11:47:33 $
xa = chir(m,n,2*a);
xb = chir(m,n,2*b);
r = xa./(xa+xb);
```
