## Supplementary material for "Quantifying non-communicable diseases’ burden in Egypt using State-Space model": S1 File: binom.m

```
function y=binom(n,k)
% BINOM(n,k) binomial coefficient

% Note that matlab already has NCHOOSEK

% Marko Laine <>
% $Revision: 1.4 $  $Date: 2012/09/27 11:47:33 $
y=exp(gammaln(n+1)-gammaln(k+1)-gammaln(n-k+1));
```
