## Supplementary material for "Quantifying non-communicable diseases’ burden in Egypt using State-Space model": S1 File: binr.m

```
function y=binr(mr,nr,n,p)
%BINPF  random numbers from binomial distribution
% BINR(mr,nr,n,p), mr, nr shape of the results, n, p parameters of
% the distribution 

% Marko Laine <>
% $Revision: 1.3 $  $Date: 2012/09/27 11:47:33 $
y = sum(rand(mr,nr,n)<=p,3);
```
