## Supplementary material for "Quantifying non-communicable diseases’ burden in Egypt using State-Space model": S1 File: bisect.m

```
function [x,y]=bisect(fun,a,b,options,varargin)
%BISECT find zero by bisection
% Usage: x0 = bisect(fun,a,b,options,varargin)
% Function must change signs between a and b.
% Use options field 'TolX' to set the 'x' tolerance.

% Marko Laine <>
% $Revision: 1.3 $  $Date: 2012/09/27 11:47:34 $

if nargin < 4, options=[]; end

tolf   = optimget(options,'TolFun',1e-12);
tolx   = optimget(options,'TolX',1e-6);
maxiter = optimget(options,'MaxIter',40);


fa = feval(fun,a,varargin{:});
fb = feval(fun,b,varargin{:});

if (fa*fb) >= 0
   error 'f(a)*f(b) not negative'
end

x = (a+b)/2;
y = feval(fun,x,varargin{:});

for i=1:maxiter
   if abs(b-a)/max(abs(b),1) < tolx || abs(y) < tolf
      return
   end
   if (fa*y)<0
      b = x;
      fb = feval(fun,b,varargin{:});
   else
      a = x;
      fa = feval(fun,a,varargin{:});
   end
   x = (a+b)/2;
   y = feval(fun,x,varargin{:});
end

warning('Maxiter exceeded')
```
