## Supplementary material for "Quantifying non-communicable diseases’ burden in Egypt using State-Space model": S1 File: bmstd.m

```
function s=bmstd(x,b)
%BMSTD standard deviation calculated from batch means
% s = bmstd(x,b) - x matrix - b length of the batch
% bmstd(x) gives an estimate of the Monte Carlo std of the 
% estimates calculated from x

% Marko Laine <>
% $Revision: 1.2 $  $Date: 2012/09/27 11:47:34 $

[n,p] = size(x);

if nargin<2
  b = max(10,fix(n/20));
end

inds = 1:b:(n+1);
nb = length(inds)-1;
if nb < 2
  error('too few batches');
end

y = zeros(nb,p);

for i=1:nb
  y(i,:)=mean(x(inds(i):inds(i+1)-1,:));
end

% calculate the estimated std of MC estimate
s = sqrt( sum((y - repmat(mean(x),nb,1)).^2)/(nb-1)*b );
```
