## Supplementary material for "Quantifying non-communicable diseases’ burden in Egypt using State-Space model": S1 File: bootstrap.m

```
function res=bootstrap(data,nboot,bootfun,varargin)
%BOOTSTRAP Generate bootstrap sample
%
% BOOTSTRAP(DATA,NBOOT,BOOTFUN,P1,P2,...)
% BOOTFUN is called as BOOTFUN(BOOTDATA,P1,P2,...), 
%   where BOOTDATA is the current bootstrap sample from DATA
%
% Examples:
%
% 1. bootstrap variance
%
%    x = randn(100,1);  % data
%    b = bootstrap(x,1000,@var);
%    hist(b)
%
% 2. bootstrap residuals to sample regression coefficients
%
%    x = [ones(10,1) (0:9)']; 
%    y = x*[1;2] + randn(10,1)*5; 
%    yfit = x*(x\y);
%    bootfun = inline('(x\(yfit+r))''','r','x','yfit')
%    b = bootstrap(y-yfit,1000,bootfun,x,yfit);
%    plot(b)
%    cov(b)   % compare covariance to one obtained from normal inference
%   inv(x'*x)*sum((y-yfit).^2)/(size(x,1)-2)

% Marko Laine <>
% $Revision: 1.2 $  $Date: 2012/09/27 11:47:34 $

ndata = size(data,1);
nx    = length(feval(bootfun,data,varargin{:}));
res   = zeros(nboot,nx);

for i=1:nboot
   bdata    = data(ceil(rand(ndata,1)*ndata),:);
   res(i,:) = feval(bootfun,bdata,varargin{:});
end
```
