## Supplementary material for "Quantifying non-communicable diseases’ burden in Egypt using State-Space model": S1 File: boxplot.m

```
function h=boxplot(y,names,outliers)
%BOXPLOT Draws a boxplot
% boxplot(y)  - draws boxplots for the columns of y

% ML 2002
% testaus : boxplot(randn(20,7))

% Marko Laine <>
% $Revision: 1.10 $  $Date: 2012/09/27 11:47:34 $

if nargin<2
  names=[];
end

if nargin<3
  outliers=1;
end

[n,m]=size(y);

if n==1
  y=y';
  n = m;
  m = 1;
end

if n<5
%  error('Must have at least 5 data points to draw a boxplot');
end

if strcmp(get(gca,'NextPlot'),'replace');cla;end
xlim([0 100]);
set(gca,'XTick',[]);

c  = linspace(0,100,2*m+1);             % centers of the boxplots
c  = c(2:2:end-1);
w1 = 5/m;                               % width of the narrow rectangle
w2 = 50/m;                              % width of the wide rectangle

if length(names)>0
  set(gca,'XTick',c);
  set(gca,'XTickLabel',names);
  %tl = get(gca,'TickLength');set(gca,'TickLength',[0 tl(2)]);
end

for i=1:m
  yi  = y(isfinite(y(:,i)),i);
  n=length(yi);
  if n<5|range(yi)<10*eps
    hold on; plot(c(i)*ones(n,1),yi,'.k'); hold off
    continue;
  end
  q   = [0.025,0.25,0.5,0.75,0.975];
  l5  = interp1(sort(yi),(n-1)*q+1);% quantiles
  med = l5(3);                          % median
  iq  = (l5(4)-l5(2))*1.5;              % inter quantile range * 1.5
%% largest y not over 1.5*IQR
  l5(1) = min(yi(yi>=med-iq));
  l5(5) = max(yi(yi<=med+iq));

  p = [c(i)-w1/2,w1,c(i)-w2/2,w2];

  if l5(2)>l5(1)
    rectangle('Position',[p(1),l5(1),p(2),l5(2)-l5(1)],'FaceColor','k');
  end
  if l5(3)>l5(2)
    rectangle('Position',[p(3),l5(2),p(4),l5(3)-l5(2)],'FaceColor','w');
  end
  if l5(4)>l5(3)
    rectangle('Position',[p(3),l5(3),p(4),l5(4)-l5(3)],'FaceColor','w');
  end
  if l5(5)>l5(4)
    rectangle('Position',[p(1),l5(4),p(2),l5(5)-l5(4)],'FaceColor','k');
  end
    
%%% outliers
 if outliers
   ol = find(abs(yi-med)>iq);
   if length(ol)>0
     hold on
     plot(c(i)*ones(size(ol)),yi(ol),'.k');
     hold off
   end
 end

 %% debug
% hold on; plot(c(i)*ones(n),y(:,i),'*b'); hold off
 
end

if nargout>0
  h = gca;
end
```
