## Supplementary material for "Quantifying non-communicable diseases’ burden in Egypt using State-Space model": S1 File: cauchydf.m

```
function y=cauchydf(x,a,b)
% CAUCHYDF Cauchy distribution function

% Marko Laine <>
% $Revision: 1.3 $  $Date: 2012/09/27 11:47:34 $
if nargin<2, a=0; end
if nargin<3, b=1; end

y=atan((x-a)./b)./pi + 0.5;
```
