## Supplementary material for "Quantifying non-communicable diseases’ burden in Egypt using State-Space model": S1 File: cauchypf.m

```
function y=cauchypf(x,a,b)
% CAUCHYPF Cauchy probability density function

% Marko Laine <>
% $Revision: 1.3 $  $Date: 2012/09/27 11:47:34 $
if nargin<2, a=0; end
if nargin<3, b=1; end

y=1./(1+((x-a)./b).^2)./pi./b;
```
