## Supplementary material for "Quantifying non-communicable diseases’ burden in Egypt using State-Space model": S1 File: cauchyqf.m

```
function y=cauchyqf(x,a,b)
% CAUCHYQF inverse of Cauchy distribution function

% Marko Laine <>
% $Revision: 1.3 $  $Date: 2012/09/27 11:47:34 $
if nargin<2, a=0; end
if nargin<3, b=1; end

y = tan(pi.*(x-0.5)).*b + a;
```
