## Supplementary material for "Quantifying non-communicable diseases’ burden in Egypt using State-Space model": S1 File: cauchyr.m

```
function y=cauchyr(m,n,a,b)
% CAUCHYR Cauchy random numbers

% Marko Laine <>
% $Revision: 1.3 $  $Date: 2012/09/27 11:47:34 $
if nargin<3, a=0; end
if nargin<4, b=1; end

y = (tan(pi*(rand(m,n)-0.5))+a).*b;
```
