## Supplementary material for "Quantifying non-communicable diseases’ burden in Egypt using State-Space model": S1 File: chainstats.m

```
function y=chainstats(chain,results,fid)
%CHAINSTATS some statistics from the MCMC chain
% chainstats(chain,results)
%    chain    nsimu*npar MCMC chain
%    results  output from mcmcrun function

% $Revision: 1.4 $  $Date: 2009/08/13 15:47:35 $

if nargin<3, fid=1; end % fid=1, standard output

if nargin>1
  if isstruct(results) % results struct from mcmcrun
    names=results.names;
  else
    names = results; % char array of names
  end
end

mcerr = bmstd(chain)./sqrt(size(chain,1));

[z,p]  = geweke(chain);
tau    = iact(chain);
stats  = [mean(chain)',std(chain)',mcerr',tau', p'];

[m,n] = size(stats);

fprintf(fid,'MCMC statistics, nsimu = %g\n\n', size(chain,1));

if nargin>1
  fprintf(fid,'% 10s ','');
end
fprintf(fid,'% 10s  % 10s  % 10s  % 10s  % 10s\n','mean','std','MC_err','tau','geweke');
if nargin>1
  fprintf(fid,'-----------');
end
fprintf(fid,'----------------------------------------------------------\n');
for i = 1:m
  if nargin>1
    fprintf(fid,'% 10s ',names{i});
  end
  fprintf(fid,'%10.5g  %10.5g  %10.5g  %10.5g  %10.5g\n',stats(i,:));
end
if nargin>1
  fprintf(fid,'-----------');
end
fprintf(fid,'----------------------------------------------------------\n');
fprintf(fid,'\n');

if nargout>0
  %  y=[stats,stats2];
  y=stats;
end

return
%% more statistics (NOT printed now)
l      = plims(chain,[0.25,0.5,0.75]);
stats2 = [min(chain)', l',max(chain)'];

if nargin>1
  fprintf(fid,'% 10s ','');
end
fprintf(fid,'% 10s  % 10s  % 10s  % 10s  % 10s\n','min','Q1','med','Q3','max');
if nargin>1
  fprintf(fid,'-----------');
end
fprintf(fid,'----------------------------------------------------------\n');
for i = 1:m
  if nargin>1
    fprintf(fid,'% 10s ',names{i});
  end
  fprintf(fid,'%10.5g  %10.5g  %10.5g  %10.5g  %10.5g\n',stats2(i,:));
end
if nargin>1
  fprintf(fid,'-----------');
end
fprintf(fid,'----------------------------------------------------------\n');
fprintf(fid,'\n');
```
