## Supplementary material for "Quantifying non-communicable diseases’ burden in Egypt using State-Space model": S1 File: chi1pf.m

```
function y=chi1pf(x,df)
%CHI1PF Chi probability density function
% CHI1PF(x,df), x value, df degrees of freedom

% Marko Laine <>
% $Revision: 1.2 $  $Date: 2012/09/27 11:47:34 $

% huom gamma parametrisointi

% chipf(x^2,df)*2*x
y = gammapf(x.^2,df/2,2)*2.*x;
```
