## Supplementary material for "Quantifying non-communicable diseases’ burden in Egypt using State-Space model": S1 File: chidf.m

```
function p=chidf(x,df,nc)
%CHIDF  Chi squared cumulative distribution function
%  CHIDF(x,df,nc) x quantile, df degrees of freedon, nc noncentrality

% Marko Laine <>
% $Revision: 1.7 $  $Date: 2013/09/05 06:54:15 $

if nargin<3,nc=0;end
  
if nc ~= 0 & exist('distribs') == 3 % mex version for non centrality
  p = distribs('chidf',x,df,nc);
  p(isnan(p)&x>0) = 1; % fix problems in fortran code for large x
else
  if nc ~= 0
    error('no noncentrality in this version')
  end
  p = gammadf(x,df/2,2);
end
```
