## Supplementary material for "Quantifying non-communicable diseases’ burden in Egypt using State-Space model": S1 File: chipf.m

```
function y=chipf(x,df)
%CHIPF Chi squared probability density function
% CHIPF(x,df), x value, df degrees of freedom

% Marko Laine <>
% $Revision: 1.6 $  $Date: 2012/09/27 11:47:34 $

% huom gamma parametrisointi
y = gammapf(x,df/2,2);
```
