## Supplementary material for "Quantifying non-communicable diseases’ burden in Egypt using State-Space model": S1 File: chiqf.m

```
function x=chiqf(p,df,nc)
%CHIQF  Inverse of chi squared distribution
% CHIQF(p,df,nc) 
%  p  probability in [0,1], 
%  df degrees of freedom, 
%  nc noncentrality (not avilable in non mex version)

% Marko Laine <>
% $Revision: 1.6 $  $Date: 2012/09/27 11:47:34 $

if nargin<3,nc=0;end
if ~isunix&exist('distribs') == 3 % mex version with non centrality
  x = distribs('chiin',p,df,nc);
else
  if any(nc~=0)
    error('non centrality not available in non win/mex version')
  end
  % uses gammaqf, the m code version of gamma quantile function
  x=gammaqf(p,df/2,2);
end
```
