## Supplementary material for "Quantifying non-communicable diseases’ burden in Egypt using State-Space model": S1 File: chir.m

```
function r=chir(m,n,df)
%CHIR    Chi squared random numbers
% usage: chir(m,n,df), m,n shape of the result, df degrees of freedom

% Marko Laine <>
% $Revision: 1.5 $  $Date: 2012/09/27 11:47:34 $
r = gammar(m,n,df/2,2);
```
