## Supplementary material for "Quantifying non-communicable diseases’ burden in Egypt using State-Space model": S1 File: confband.m

```
function out=confband(x,y,s,k,varargin)
%CONFBAND confidence band
% confband(x,y,ystd) plots x vs. y with confidence bands from ystd.
% confband(x,y,ystd,k) uses [y-k*ystd, y+k*ystd], default k=2.

%, 2013
% $Revision: 1.2 $  $Date: 2013/02/14 11:29:48 $

colo  = [0.0 0.5 1.0]; % lines
colog = [0.9 0.9 0.9]; % grey area

if nargin < 4
  k = 2;
end

hf = fillyy(x,y-k*s,y+k*s,colog);
set(hf,'FaceAlpha',0.3); % transparency

ih = ishold;
hold on
h1 = plot(x,y,     '-','linewidth',2,'color',colo,varargin{:});
h2 = plot(x,y-k*s, ':','linewidth',1,'color',colo);
h3 = plot(x,y+k*s, ':','linewidth',1,'color',colo);

if ~ih
  hold off
end

if nargout>0
  out=[hf;h1;h2;h3];
end
```
