## Supplementary material for "Quantifying non-communicable diseases’ burden in Egypt using State-Space model": S1 File: Contents.m

```
% MCMCSTAT toolbox for MCMC statistical analysis
% Version 1.0 2007-08-08
%
% MCMC run and plots 
%
% mcmcrun    - adaptive Metropolis - Hastings Markov chain Monte Carlo
% mcmcplot   - plot mcmc chain
% mcmcpred   - predictive calculations from the chain
% chainstats - some statistics of the chain matrix
%
% Convergence diagnostics
%
% cusum      - Cusum plot
% geweke     - Geweke's MCMC convergence diagnostic
% gewekeplot - Plot Geweke's diagnostic for increasing number of iterations
% psrf       - Gelman-Rubin-Brooks potential scale reduction factor
% mpsrf      -        multivariate potential scale reduction factor
% psrfplot   - plot psrf for increasing number of iterations
% hwdiag     - Heidelberger-Welch MCMC convergence diagnostics
%
% Statistical functions
% elementary:
%  range       - range
%  iqrange     - interquantile range
%  covupd      - covariance update
%  mad         - median absolute deviation
%  means       - summary statistics for columns
%  logit       - logit transformation
%  meantrim    - trimmed mean
%  plims       - empirical quantiles
%  binom       - binomial coefficient
%  cov2cor     - covariance matrix into correlation matrix
%  covcond     - make covariance matrix with given cond number
%
% distributions: probability/density, distribution, quantile, random
%  norpf, nordf, norqf, norr             - Gaussian 
%  lognorpf, lognordf, lognorqf, lognorr - lognormal
%  gammapf, gammadf, gammaqf, gammar     - Gamma
%  chipf, chidf, chiqf, chir             - Chi squared
%  tpf, tdf, tqf, tr                     - t
%  fpf, fdf, fqf, fr                     - F
%  invchipf, invchir                     - Inverse chi squared
%  invchi1pf, invchi1r                   - Inverse chi
%  cauchypf, cauchydf, cauchyqf, cauchyr - Cauchy
%  betapf, betadf, betaqf, betar         - Beta
%  exppf, expdf, expqf, expr             - Exponential
%  dexppf, dexpdf, dexpr                 - Laplace (double exponential)
%  poipf, poidf, poir                    - Poisson
%  logipf, logidf, logir                 - Logistic
%  extpf, extdf, extr                    - Extreme value
%
%  bootstrap   - generate bootstrap sample
% 
% plots:
%  density     - Kernel density estimator
%  density2d   - 2d kernel estimator
%  histp       - histogram scaled as probabilities
%  pairs       - pairs plot
%  boxplot     - box plot
%  qqnor       - quantile-quantile plot for normality
%  qqplot      - general quantile-quantile plot
%  xquad       - integrate data using Hermite spline
%  xyplot      - plot two first columns of xy matrix
%  removepoints- remove point from a plot
%  hline       - add horizontal / vertical line
%  ellipse     - draw an ellipse
%  fillxx      - fill a space between lines with a color
%  fillyy      - fill a space between lines with a color
%  epsfile     - dump figure into an eps file

% beware: not all functions are well tested!
```
