## Supplementary material for "Quantifying non-communicable diseases’ burden in Egypt using State-Space model": S1 File: covcond.m

```
function [Sig,Lam] = covcond(c,a)
%COVCOND covariance matrix with given condition number
% [Sig,Lam] = covcond(condnum,dire) generates covariance matrix and its
% inverse with given cond number and first direction.

% $Revision: 1.4 $  $Date: 2017/01/23 14:19:23 $

% create orthogonal basis z, with 1. direction given by 'a'
a     = a(:);
e     = fliplr(1./linspace(c,1,length(a)));
a(1)  = a(1) + sign(a(1)) * norm(a);  	% the Householder trick 
z     = eye(length(a)) - 2.0/norm(a)^2*a*a'; 
Sig   = z * diag(e) * z' ;              % target covariance
Lam   = z * diag(1./e) * z';            % and its inverse
```
