## Supplementary material for "Quantifying non-communicable diseases’ burden in Egypt using State-Space model": S1 File: covupd.m

```
function [xcov,xmean,wsum,R]=covupd(x,w,oldcov,oldmean,oldwsum,oldR)
%COVUPD covariance update
% [xcov,xmean,wsum]=covupd(x,w,oldcov,oldmean,oldwsum)

% optionally updates also the Cholesky factor R

% Marko Laine <>
% $Revision: 1.4 $  $Date: 2012/09/27 11:47:34 $

[n,p]=size(x);
if n == 0 % nothing to update with
  xcov = oldcov; xmean = oldmean; wsum = oldwsum;
  return
end

if nargin<2 | isempty(w)
  w = 1;
end
if length(w) == 1
  w = ones(n,1)*w;
end

if nargin < 6 | isempty(oldR)
  R = [];
else
  R = oldR;
end

if nargin>2 & ~isempty(oldcov) % update

  for i=1:n
    xi     = x(i,:);
    wsum   = w(i);
    xmeann = xi;
    xmean  = oldmean + wsum/(wsum+oldwsum)*(xmeann-oldmean);

    if ~isempty(R)
      R = cholupdate(sqrt((oldwsum-1)/(wsum+oldwsum-1))*R, ...
                     (xi-oldmean)'* ...
                     sqrt((wsum*oldwsum)/(wsum+oldwsum-1)/(wsum+oldwsum)));
    end
    
    xcov =  (oldwsum-1)./(wsum+oldwsum-1).*oldcov + ...
            wsum.*oldwsum/(wsum+oldwsum-1)./(wsum+oldwsum) .* ...
             ((xi-oldmean)' *(xi-oldmean));
    wsum    = wsum+oldwsum;
    oldcov  = xcov;
    oldmean = xmean;
    oldwsum = wsum;
  end
  
else % no update

  wsum  = sum(w);
  xmean = zeros(1,p);
  xcov  = zeros(p,p);
  for i=1:p
    xmean(i) = sum(x(:,i).*w)./wsum;
  end
  if wsum>1
    %%% (wsum-oldwsum/wsum)
    for i=1:p
      for j=1:i
        xcov(i,j) = (x(:,i)-xmean(i))' * ((x(:,j)-xmean(j)).*w)./(wsum-1);
        if (i ~= j)
          xcov(j,i) = xcov(i,j);
        end
      end
    end
  end

  if nargout>3
    [R,p] = chol(xcov);
    if p~=0
      R=[];
    end
  end
  
end
```
