## Supplementary material for "Quantifying non-communicable diseases’ burden in Egypt using State-Space model": S1 File: cramerdf.m

```
function y=cramerdf(q)
% CRAMERDF  Distribution function of the Cramer-von Mises statistic
% y = cramerdf(q), q quantile, y cumulative probability

% based on the corresponding R version

% Marko Laine <>
% $Revision: 1.4 $  $Date: 2012/09/27 11:47:34 $

logeps = log(1e-5);
y = zeros(4,length(q));
for k = 0:3 
  z =  gamma(k+0.5)*sqrt(4*k+1)./(gamma(k+1)*pi^(3/2).*sqrt(q));
  u = (4 *k+1)^2./(16*q);
  ind = (u<=-logeps);
  y(k+1,~ind) = 0;
  y(k+1, ind) = z(ind).*exp(-u(ind)).*besselk(1/4,u(ind));
end
y = sum(y);
```
