## Supplementary material for "Quantifying non-communicable diseases’ burden in Egypt using State-Space model": S1 File: cusum.m

```
function [d,c,st]=cusum(chain)
%CUSUM  Cusum diagnostic plot for MCMC chain
% [d,c,st]=cusum(chain)
% Returns Brooks statistics d, its 95% confidence limits c, and
% the cumulative sum st.
% See:
% Stephen P. Brooks and Gareth O. Roberts.
% Assessing convergence of Markov chain Monte Carlo algorithms.
% Statistics and Computing, 8:319--335, 1998.

% this is cusum for mean of the chain

% ML, 2002
% $Revision: 1.3 $  $Date: 2003/05/07 12:22:19 $

[nsimu,npar] = size(chain);
mu = mean(chain);
st = cumsum(chain-repmat(mu,nsimu,1));
%h  = plot(st);
for i=1:npar
  subplot(npar,1,i);
  plot(st(:,i));
end

%% Brooks statistic
%% does not work on MH chain ??
d = sum(abs(diff(sign(diff(st))))==2)/(nsimu-1);
a = 0.95;
c = 0.5 + [1 -1].*norqf((1+a)/2)*sqrt(1/4/(nsimu-1));
```
