## Supplementary material for "Quantifying non-communicable diseases’ burden in Egypt using State-Space model": S1 File: datamerge.m

```
function xy = datamerge(x,y,intinds)
%DATAMERGE Merge matrises by first columns
% datamerge(x,y) - merges matrises x and y by their first columns
% matrises should be sorted

% Marko Laine <>
% $Revision: 1.6 $  $Date: 2012/09/27 11:47:34 $

if nargin < 3
  intinds=[]; % interpolation for these colums
end

byind=1; % index column

if isempty(x)
  xy=y;
  return;
end

if min(diff(x(:,byind)))<=0 | min(diff(y(:,byind)))<=0
  error('indexes should be strictly increasing');
end

%x = sortrows(x,byind);
%y = sortrows(y,byind);

[nx,mx]=size(x);
[ny,my]=size(y);

[xyby,ix,iy] = union(x(:,byind),y(:,byind));

xy = zeros(length(xyby),mx+my-1)*NaN;
xy(:,1) = xyby;

xnoby = 1:mx~=byind;
ynoby = 1:my~=byind;

% intersect does the trick
[c,i1,i2] = intersect(xyby,x(:,byind));
xy(i1,2:mx) = x(:,xnoby);
[c,i1,i2] = intersect(xyby,y(:,byind));
xy(i1,mx+1:end) = y(:,ynoby);

% interpolation for intinds columns
for i=intinds
  inds = ~isnan(xy(:,i));
  xy(~inds,i) = interp1(xy(inds,byind),xy(inds,i),xy(~inds,byind),'linear','extrap');
end
```
