## Supplementary material for "Quantifying non-communicable diseases’ burden in Egypt using State-Space model": S1 File: demean.m

```
function y=demean(x)
%DEMEAN  remove column means

% Marko Laine <>
% $Revision: 1.4 $  $Date: 2012/09/27 11:47:35 $
[m,n] = size(x);
if m==1 | n==1
  y = x - mean(x);
else
  y = x - repmat(sum(x)/m,m,1);
end
```
