## Supplementary material for "Quantifying non-communicable diseases’ burden in Egypt using State-Space model": S1 File: density2d.m

```
function [z,xo,yo,s]=density2d(x,xout,yout,ss,rho,plotit)
%DENSITY2D   2 dimensinal density estimator
% [z,x,y]=density2d(x,xout,yout,s,rho,plotit)
% x size n*2 data used for estimation
% xout  1. coordinate points returned (optional) 
% yout  2. coordinate points returned (optional)
% s relative smoothing factor (default = 1)
% rho correlation coefficient of the Gaussian kernel used (optional)
% plotit 0 = no plot, 1 = contour plot, 2 = mesh plot
%
% output: z,x,y cordinates of the estimator 

% ML 2000, see MASS 2nd ed, page 184

% Marko Laine <>
% $Revision: 1.6 $  $Date: 2012/09/27 11:47:35 $

if size(x,2)~= 2
   error('size(x,2)~=2')
end
nx=size(x,1);
if nargin<2 | isempty(xout)
   xmin=min(x(:,1)); xmax=max(x(:,1)); xrange=xmax-xmin;
   xout=linspace(xmin-0.06*xrange,xmax+0.06*xrange,25);
end
if nargin<3 | isempty(yout)
   xmin=min(x(:,2)); xmax=max(x(:,2)); xrange=xmax-xmin;
   yout=linspace(xmin-0.06*xrange,xmax+0.06*xrange,25);
end
s1=1.06*min(std(x(:,1)),iqrange(x(:,1))/1.34)*nx^(-1/6); % -1/5
s2=1.06*min(std(x(:,2)),iqrange(x(:,2))/1.34)*nx^(-1/6); % 
%% fixme, when iqrange = 0
if s1 == 0
  s1 = 1.06*std(x(:,1))*nx^(-1/6);
end
if s2 == 0
  s2 = 1.06*std(x(:,2))*nx^(-1/6);
end
s=[s1,s2];
if nargin>3 & ~isempty(ss)
   s=s.*ss;
end
if nargin<5 | isempty(rho)
% rho=0;
  rho4=corrcoef(x); rho=rho4(1,2);
else
  if abs(rho)>=1
    error('rho should be between -1 and 1')
  end
end
if nargin<6
   plotit=0;
end

[X,Y]=meshgrid(xout,yout);

[mX,nX]=size(X);
z=zeros(mX,nX);

r = 1-rho.^2;
c = 1./(2*pi*s(1)*s(2)*sqrt(r));
for i=1:(mX*nX)
  if 0
   z(i)=1/nx*sum(norpf((X(i)-x(:,1))/s(1)).*...
      norpf((Y(i)-x(:,2))/s(2)))/prod(s);
  else
   z(i) = 1./nx .* sum(c * exp(-0.5/r*( ...
          ((X(i)-x(:,1))./s(1)).^2 - ...
       2*rho*(X(i)-x(:,1))./s(1).*(Y(i)-x(:,2))./s(2) + ...
          ((Y(i)-x(:,2))./s(2)).^2 )));
  end
end

if nargout>1
   xo=xout;
end
if nargout>2
   yo=yout;
end

if plotit==1 | nargout == 0
   contour(xout,yout,z); 
   hold on;
   plot(x(:,1),x(:,2),'.'); 
   hold off
elseif plotit==2
   mesh(xout,yout,z);
%   hold on;
%   plot(x(:,1),x(:,2),'o'); 
%   hold off 
end
```
