## Supplementary material for "Quantifying non-communicable diseases’ burden in Egypt using State-Space model": S1 File: density.m

```
function [y,xo]=density(x,xout,ss,gaus)
%DENSITY  Density estimator using Gaussian kernel
% Y = DENSITY(X,XOUT,S)
% X is the vector of data values.
% The density estimator is evaluated at XOUT points.
% S is a scale factor for the default kernel bandwidth,
% default S = 1.
% Without output arguments the density is plotted.

% Marko Laine <>
% $Revision: 1.11 $  $Date: 2014/09/28 17:46:32 $

if nargin<3
  ss=1;
end
if nargin<4
  gaus=1;
end

if nargin<2 | isempty(xout)
  xmin=min(x); xmax=max(x); xrange=xmax-xmin;
  if length(x) > 200
    xout=linspace(xmin-0.08*xrange,xmax+0.08*xrange);
  else
    xout=linspace(mean(x)-4*std(x),mean(x)+4*std(x));
  end
end
y  = zeros(size(xout));
n  = length(xout);
nx = length(x);

%%% see MASS 2nd ed page 181.
if iqrange(x)<=0
  s=1.06*std(x)*nx^(-1/5);
else
  s=1.06*min(std(x),iqrange(x)/1.34)*nx^(-1/5);
end
%  s=1.144*std(x)*nx^(-1/5);
if ss>0
  s=ss*s;
elseif ss<0
  s = abs(ss);
end
if gaus==1
  % Gaussian kernel
  for i=1:n
    y(i) = 1/nx*sum(norpf((xout(i)-x)/s))./s;
  end
elseif gaus == 0
  % triangular kernel
  s=s*1.2113;
  for i=1:n
    y(i) = 1/nx*sum(max(0,1-abs(xout(i)-x)/s))./s;
  end
else
  % Gamma kernel
  if std(x)/mean(x) > 0.9
    b = s;
  elseif std(x)/mean(x) > 0.4
    b = s/2;
  elseif std(x)/mean(x) > 0.2
    b = s/5;
  else
    b = s/40; % how to choose this?
  end
  ii = x >= 2*b;
  for i=1:n    
    if xout(i)>0
      y(i) = 1/nx*sum(gammapf(xout(i),x(ii)/b,b));
      y(i) = y(i) + 1/nx*sum(gammapf(xout(i),(x(~ii)/2/b).^2+1,b));
%      y(i) = 1/nx*sum(gammapf(xout(i),x/b+1,b));
    end
  end  
end

if nargout>1
  xo=xout;
end

if nargout==0
  plot(xout,y)
  clear y % no output
end
```
