## Supplementary material for "Quantifying non-communicable diseases’ burden in Egypt using State-Space model": S1 File: dexpdf.m

```
function y=dexpdf(x,a,b)
%DEXPDF  Laplace cumulative density function
% dexpdf(x,a,b)

% Marko Laine <>
% $Revision: 1.4 $  $Date: 2012/09/27 11:47:35 $

if nargin<=1, a=0; end
if nargin<=2, b=1; end

y=0.5*(1 + sign(x-a).*(1-exp(-abs(x-a)./b)));
```
