## Supplementary material for "Quantifying non-communicable diseases’ burden in Egypt using State-Space model": S1 File: dexpr.m

```
function y=dexpr(m,n,a,b)
%DEXPR   Random numbers from Laplace distribution
% dexpr(m,n,a,b)

% Marko Laine <>
% $Revision: 1.6 $  $Date: 2012/09/27 11:47:35 $
if nargin<=2, a=0; end
if nargin<=3, b=1; end

y=b*(log(1-rand(m,n))-log(1-rand(m,n))+a);
```
