## Supplementary material for "Quantifying non-communicable diseases’ burden in Egypt using State-Space model": S1 File: algaeex.html

MCMC toolbox » Examples » Algae

### Algae example

The example uses functions algaesys, algaefun and algaess.

We study the following system

This is a simplified lake algae dynamics model. We consider phytoplankton *A*, zooplankton *Z* and nutrition *P* (eg. phosphorus) available for *A* in the water. The system is affected by the water outflow/inflow *Q*, incoming phosphorus load *Pin* and temperature *T*. It is described as a simple predator - pray dynamics between *A* and *Z*. The growth of *A* is limited by the availability of *P* and it depends on the water temperature *T*. The inflow/outflow *Q* affects both *A* and *P*, but not *Z*. We use the following equations:

dA/dt = (μ - ρ_a_ - Q/V - αZ) A  
dZ/dt = αZA-ρ_z_ Z  
dP/dt = -Q/V (P-P_in_) +
(ρ_a_-μ)A+ρ_z_Z

where the growth rate µ depends on both temperature *T* and phosphorus *P*

μ = μ^max^θ^(T-20)^P/(k+P).

The data set is stored in algaedata.mat. First we load and plot the data.

```
clear model data params options
load algaedata.mat
figure(1); clf
for i =1:3
  subplot(2,3,i)
  plot(data.xdata(:,1),data.xdata(:,i+1),'-k');
  title(data.xlabels(i+1)); xlim([1,120])
end
subplot(2,1,2)
plot(data.ydata(:,1),data.ydata(:,2:end),'o-');
title('model state variable observations');
legend(data.ylabels(2:end),'Location','best');
xlabel('days');
```

The model sum of squares in file algaess.m is given in the model structure.

```
model.ssfun = @algaess;
```

All parameters are constrained to be positive. The initial concentrations are also unknown and are treated as extra parameters.

```
params = {
    {'mumax', 0.5,  0}
    {'rhoa',  0.03, 0}
    {'rhoz',  0.1,  0}
    {'k',     10,   0}
    {'alpha', 0.02, 0}
    {'th',    1.14, 0, Inf, 1.14, 0.2}  % N(0.14, 0.2^2)1{th>0} prior
% initial values for the model states
    {'A0', 0.77, 0, Inf, 0.77, 2 }
    {'Z0', 1.3,  0, Inf, 1.3,  2 }
    {'P0', 10,   0, Inf, 10,   2 }
    };
```

We assume having at least some prior information on the repeatability of the observation and assign rather non informational prior for the residual variances of the observed states. The default prior distribution is sigma2 ~ invchisq(S20,N0), the inverse chi squared distribution (see for example Gelman et al.). The 3 components (*A*, *Z*, *P*) all have separate variances.

```
model.S20 = [1 1 2];
model.N0  = [4 4 4];
```

First generate an initial chain.

```
options.nsimu = 1000;
[results, chain, s2chain]= mcmcrun(model,data,params,options);
```

```
Sampling these parameters:
name   start [min,max] N(mu,s^2)
mumax: 0.5 [0,Inf] N(0,Inf)
rhoa: 0.03 [0,Inf] N(0,Inf)
rhoz: 0.1 [0,Inf] N(0,Inf)
k: 10 [0,Inf] N(0,Inf)
alpha: 0.02 [0,Inf] N(0,Inf)
th: 1.14 [0,Inf] N(1.14,0.2^2)
A0: 0.77 [0,Inf] N(0.77,2^2)
Z0: 1.3 [0,Inf] N(1.3,2^2)
P0: 10 [0,Inf] N(10,2^2)
```

Then re-run starting from the results of the previous run, this will take couple of minutes.

```
options.nsimu = 5000;
[results, chain, s2chain] = mcmcrun(model,data,params,options, results);
```

```
Using values from the previous run
Sampling these parameters:
name   start [min,max] N(mu,s^2)
mumax: 0.398492 [0,Inf] N(0,Inf)
rhoa: 0.0348433 [0,Inf] N(0,Inf)
rhoz: 0.0982238 [0,Inf] N(0,Inf)
k: 9.4219 [0,Inf] N(0,Inf)
alpha: 0.0233986 [0,Inf] N(0,Inf)
th: 1.00471 [0,Inf] N(1.14,0.2^2)
A0: 1.16216 [0,Inf] N(0.77,2^2)
Z0: 2.07551 [0,Inf] N(1.3,2^2)
P0: 8.54463 [0,Inf] N(10,2^2)
```

Chain plots should reveal that the chain has converged and we can use the results for estimation and predictive inference.

```
figure(2); clf
mcmcplot(chain,[],results,'pairs');
figure(3); clf
mcmcplot(chain,[],results,'denspanel',2);
```

 

Function chainstats calculates mean ans std from the chain and estimates the Monte Carlo error of the estimates. Number tau is the integrated autocorrelation time and geweke is a simple test for a null hypothesis that the chain has converged.

```
chainstats(chain,results)
```

```
MCMC statistics, nsimu = 5000

                 mean         std      MC_err         tau      geweke
---------------------------------------------------------------------
     mumax     7.3133      8.9169      1.8501       612.9    0.034271
      rhoa   0.026332    0.014272    0.002472      416.33     0.27418
      rhoz   0.097931   0.0057379  0.00069317      81.933      0.9884
         k      346.6      434.88      90.632      615.33    0.030661
     alpha   0.023557   0.0013095  0.00015524      92.634      0.9452
        th     1.0123    0.013533   0.0016359      114.66     0.98836
        A0     1.0649     0.30144    0.024486      37.659      0.9064
        Z0     1.8118     0.45222    0.050899      66.175     0.82012
        P0      8.822     0.85789    0.065738      37.405     0.98279
---------------------------------------------------------------------
```

In order to use the mcmcpred function we need function modelfun with input arguments given as modelfun(xdata,theta). We construct this as an anonymous function.

```
modelfun = @(d,th) algaefun(d(:,1),th,th(7:9),d);
```

We sample 500 parameter realizations from chain and s2chain and calculate the predictive plots.

```
nsample = 500;
out = mcmcpred(results,chain,s2chain,data.xdata,modelfun,nsample);
figure(4); clf
mcmcpredplot(out);
% add the 'y' observations to the plot
hold on
for i=1:3
  subplot(3,1,i)
  hold on
  plot(data.ydata(:,1),data.ydata(:,i+1),'s');
  ylabel(''); title(data.ylabels(i+1));
  hold off
end
xlabel('days');
```

Published with MATLAB® R2018b
