## Supplementary material for "Quantifying non-communicable diseases’ burden in Egypt using State-Space model": S1 File: algaess.html

```
function ss = algaess(theta,data)
% algae sum-of-squares function

time   = data.ydata(:,1);
ydata  = data.ydata(:,2:end);
xdata  = data.xdata;

% 3 last parameters are the initial states
y0 = theta(end-2:end);

ymodel = algaefun(time,theta,y0,xdata);
ss = sum((ymodel - ydata).^2);
```

Published with MATLAB® R2018b
