## Supplementary material for "Quantifying non-communicable diseases’ burden in Egypt using State-Space model": S1 File: algaesys.html

```
function ydot = algaesys(t,y,theta,xdata)
% ode system function for MCMC algae example

A = y(1);
Z = y(2);
P = y(3);

% control variables are assumed to be saved
% at each time unit interval
QpV = xdata(ceil(t),2);
T   = xdata(ceil(t),3);
Pin = xdata(ceil(t),4);

mumax = theta(1);
rhoa  = theta(2);
rhoz  = theta(3);
k     = theta(4);
alpha = theta(5);
th    = theta(6);

mu = mumax*th^(T-20)*P/(k+P);

dotA = (mu - rhoa - QpV -alpha*Z)*A;
dotZ = alpha*Z*A - rhoz*Z;
dotP = -QpV*(P-Pin) + (rhoa-mu)*A + rhoz*Z;

ydot=[dotA;dotZ;dotP];
```

Published with MATLAB® R2018b
