## Supplementary material for "Quantifying non-communicable diseases’ burden in Egypt using State-Space model": S1 File: bananaex.html

MCMC toolbox » Examples » Banana

### Banana example

This techncal example constructs a non Gaussian target distribution by twisting two first dimensions of Gaussian distribution. The Jacobian of the transformation is 1, so it is easy to calculate the right probability regions for the banana and study different adaptive methods.

```
clear model data params options
```

'banana' sum-of-squares

```
bananafun = @(x,a,b) [a.*x(:,1),x(:,2)./a-b.*((a.*x(:,1)).^2+a^2),x(:,3:end)];
bananainv = @(x,a,b) [x(:,1)./a,x(:,2).*a+a.*b.*(x(:,1).^2+a^2),x(:,3:end)];
bananass  = @(x,d) bananainv(x-d.mu,d.a,d.b)*d.lam*bananainv(x-d.mu,d.a,d.b)';
```

```
a = 1; b = 1;         % banana parameters

npar = 12;             % number of unknowns
rho  = 0.9;            % target correlation
sig  = eye(npar); sig(1,2) = rho; sig(2,1) = rho;
lam  = inv(sig);       % target precision
mu   = zeros(1,npar);  % center
```

the data structure and parameters

```
data = struct('mu',mu,'a',a,'b',b,'lam',lam);
for i=1:npar
  params{i} = {sprintf('x_%d',i),0};
end

model.ssfun     = bananass;
model.N         = 1;

options.method  = 'dram';
options.nsimu   = 10000;
options.qcov    = eye(npar)*0.5; % [initial] proposal covariaance
```

```
[results,chain] = mcmcrun(model,data,params,options);
```

```
Setting nbatch to 1
Sampling these parameters:
name   start [min,max] N(mu,s^2)
x_1: 0 [-Inf,Inf] N(0,Inf)
x_2: 0 [-Inf,Inf] N(0,Inf)
x_3: 0 [-Inf,Inf] N(0,Inf)
x_4: 0 [-Inf,Inf] N(0,Inf)
x_5: 0 [-Inf,Inf] N(0,Inf)
x_6: 0 [-Inf,Inf] N(0,Inf)
x_7: 0 [-Inf,Inf] N(0,Inf)
x_8: 0 [-Inf,Inf] N(0,Inf)
x_9: 0 [-Inf,Inf] N(0,Inf)
x_10: 0 [-Inf,Inf] N(0,Inf)
x_11: 0 [-Inf,Inf] N(0,Inf)
x_12: 0 [-Inf,Inf] N(0,Inf)
```

```
figure(1); clf
mcmcplot(chain,[],results.names,'chainpanel')
```

```
figure(2); clf
mcmcplot(chain,[1,2],results.names,'pairs',0)
c50=1.3863; % critical values from chisq(2) distribution
c95=5.9915;
hold on
[xe,ye]=ellipse(mu,c50*sig(1:2,1:2));
xyplot(bananafun([xe,ye],a,b),'k-','LineWidth',2)
[xe,ye]=ellipse(mu,c95*sig(1:2,1:2));
xyplot(bananafun([xe,ye],a,b),'k-','LineWidth',2)
axis equal
hold off
title('2 first dimensions of the chain with 50% and 95% target contours')
```

Published with MATLAB® R2018b
