## Supplementary material for "Quantifying non-communicable diseases’ burden in Egypt using State-Space model": S1 File: beetleex.html

MCMC toolbox » Examples » Beetle

### Beetle mortality data

From A. Dobson, *An Introduction to Generalized Linear Models*, Chapman & Hall/CRC, 2002. Binomial logistic regression example with dose-response data. See beetless.m for -2log(likelihood) function.

```
clear model data params options

data = [
% dose   n  y
  1.6907 59  6
  1.7242 60 13
  1.7552 62 18
  1.7842 56 28
  1.8113 63 52
  1.8369 59 53
  1.8610 62 61
  1.8839 60 60
    ];

global BEETLE_LINK
BEETLE_LINK = 2; % 1=logit, 2=loglog, 3=probit

% the "sum-of-squares" is now -2log(likelihood) of the binomial model
model.ssfun = @beetless;

% initial values and model function according to the link function
switch BEETLE_LINK
 case 1
  b = [ 60,-35]; % logit
  modelfun = @(d,th) 1./(1+exp(th(1)+th(2).*d));
  label = 'Beetle data with logit link';
 case 2
  b = [-40, 22]; % logog
  modelfun = @(d,th) 1-exp(-exp(th(1)+th(2).*d));
  label = 'Beetle data with loglog link';
 case 3
  b = [-35, 20]; % probit
  modelfun = @(d,th) nordf(th(1)+th(2).*d);
  label = 'Beetle data with probit link';
end

% model parameters
params = {
    {'b_0', b(1)}
    {'b_1', b(2)}
    };

options.nsimu = 5000;

[res,chain] = mcmcrun(model,data,params, options);

% plot the chain
figure
mcmcplot(chain,[],res)
figure
mcmcplot(chain,[],res,'pairs')

% sample the predicted mean respose
out = mcmcpred(res,chain,[],linspace(1.5,2)',modelfun,500);
figure
mcmcpredplot(out)
hold on % add data points
plot(data(:,1),data(:,3)./data(:,2),'ok')
hold off
title(label)
ylabel('proportion killed')
xlabel('log dose')
```

```
Sampling these parameters:
name   start [min,max] N(mu,s^2)
b_0: -40 [-Inf,Inf] N(0,Inf)
b_1: 22 [-Inf,Inf] N(0,Inf)
```

  

Published with MATLAB® R2018b
