## Supplementary material for "Quantifying non-communicable diseases’ burden in Egypt using State-Space model": S1 File: beetless.html

```
function ss = beetless(theta,data)
% Beatle mortality example binomial -2*log(likelihood) function
% -2 log(likelihood) = -2*sum( y log(p) + (n-y) log(1-p) )

dose = data(:,1);
n    = data(:,2);
y    = data(:,3);

global BEETLE_LINK

switch BEETLE_LINK
 case 1
  % fitted probability from logistic model
  p  = 1./(1+exp(theta(1) + theta(2).*dose));
 case 2
  % loglog model
  p = 1-exp(-exp(theta(1)+theta(2).*dose));
 case 3
  % probit model
  p = nordf(theta(1) + theta(2).*dose);
end

ss = -2*sum(y.*log(p) + (n-y).*log(1-p));
```

Published with MATLAB® R2018b
