## Supplementary material for "Quantifying non-communicable diseases’ burden in Egypt using State-Space model": S1 File: boxoex.html

MCMC toolbox » Examples » Boxo

### Boxo chemical kinetics example.

Chemical kinetics example. We model reactions *A* -> *B* -> *C*, with Arrhenius temperature dependence in the reaction rates. Model is an ode system. See boxoM.m for the model function and boxoODE.m for the ode system function.

Model unknowns are reaction rate parameters *k1* and *k2*, activation energies *E1* and *E2* and the initial concentrations of *A* in both the batches (total of 6 unknowns).

```
% This will take some time if |boxoM| does not use |lsode_mex|.

clear model data params options
```

Set some parameters for the run.

```
method      = 'dram'; % adaptation method, 'mh', 'dr', 'am', or 'dram'
nsimu       = 5000;   % number of simulations
adaptint    = 500;    % how often to adapt the proposal
```

### Data

We have two data sets (=batches).

```
data{1}.ydata = [
%  time    A      B
   0   1.000   0.000
   1   0.504   0.416
   2   0.186   0.489
   3   0.218   0.595
   4   0.022   0.506
   5   0.102   0.493
   6   0.058   0.458
   7   0.064   0.394
   8   0.000   0.335
   9   0.082   0.309
];
data{2}.ydata = [
%  time    A       B
   0   1.000   0.000
   1   0.415   0.518
   2   0.156   0.613
   3   0.196   0.644
   4   0.055   0.444
   5   0.011   0.435
   6   0.000   0.323
   7   0.032   0.390
   8   0.000   0.149
   9   0.079   0.222
];
```

### Model parameters

The initial values for *A* and *B* and the (fixed) temperatures are local to the batches. Initial value for *A* has some error in it, but *B* is assumed to be exactly 0.

```
params = {
%      name,  init,        min, max, mu,  sig, target?, local?
    {'k1mean', 1.0,        0,  Inf,  NaN, Inf,   1,      0}
    {'E1'    , 0.01,       0,  Inf,  NaN, Inf,   1,      0}
    {'k2mean', 1.0,        0,  Inf,  NaN, Inf,   1,      0}
    {'E2',     0.01,       0,  Inf,  NaN, Inf,   1,      0}
    {'Tmean',  300,      -Inf, Inf,  NaN, Inf,   0,      0}
    {'Temp' ,  [283 313],  0,  0,    NaN, Inf,   0,      1}
    {'A0',     [1.0 1.0],  0,  Inf,  1,   0.1,   1,      1}
    {'B0',     [0.0 0.0],  0,  Inf,  NaN, Inf,   0,      1}
         };
```

### Model options

```
% model.ssfun     = @boxoSS;
model.modelfun   = @boxoM; % use mcmcrun generated ssfun instead
model.sigma2     = 0.01;   % initial error variance
model.N0         = 4;      % prior (invchisq) weight for sigma2

options.method      = method;        % adaptation method (mh,am,dr,dram)
options.nsimu       = nsimu;         % n:o of simulations
options.qcov        = eye(11)*0.001; % proposal covariance
options.adaptint    = adaptint; % adaptation interval
options.printint    = 200; % how often to show info on acceptance ratios
options.verbosity   = 1;  % how much to show output in Matlab window
options.waitbar     = 1;  % show garphical waitbar
options.updatesigma = 1;  % update error variance
options.stats       = 1;  % save extra statistics in results
```

### MCMC run

As we start from non optimized values the chain will need some time to find the location of the posterior. We do 3 runs, starting from the values of the previous run.

```
results = [];
[results,chain,s2chain,sschain]=mcmcrun(model,data,params,options,results);
[results,chain,s2chain,sschain]=mcmcrun(model,data,params,options,results);
[results,chain,s2chain,sschain]=mcmcrun(model,data,params,options,results);
```

```
Sampling these parameters:
name   start [min,max] N(mu,s^2)
k1mean: 1 [0,Inf] N(1,Inf)
E1: 0.01 [0,Inf] N(0.01,Inf)
k2mean: 1 [0,Inf] N(1,Inf)
E2: 0.01 [0,Inf] N(0.01,Inf)
A0[1]: 1 [0,Inf] N(1,0.1^2)
A0[2]: 1 [0,Inf] N(1,0.1^2)
Using values from the previous run
Sampling these parameters:
name   start [min,max] N(mu,s^2)
k1mean: 0.751341 [0,Inf] N(1,Inf)
E1: 0.00591968 [0,Inf] N(0.01,Inf)
k2mean: 0.211658 [0,Inf] N(1,Inf)
E2: 0.00648117 [0,Inf] N(0.01,Inf)
A0[1]: 0.987673 [0,Inf] N(1,0.1^2)
A0[2]: 1.05326 [0,Inf] N(1,0.1^2)
Using values from the previous run
Sampling these parameters:
name   start [min,max] N(mu,s^2)
k1mean: 0.793011 [0,Inf] N(1,Inf)
E1: 0.00572464 [0,Inf] N(0.01,Inf)
k2mean: 0.209713 [0,Inf] N(1,Inf)
E2: 0.00922497 [0,Inf] N(0.01,Inf)
A0[1]: 1.01171 [0,Inf] N(1,0.1^2)
A0[2]: 1.02802 [0,Inf] N(1,0.1^2)
```

### Chain plots

```
figure(1); clf
mcmcplot(chain,[],results,'chainpanel');
figure(2); clf
mcmcplot(chain,[],results,'pairs');
```

 

### Plot of the data and the mean posterior fit

```
mcmean = mean(chain); % posterior mean parameter vector
figure(3); clf
for i=1:results.nbatch
  subplot(2,1,i)
  plot(data{i}.ydata(:,1),data{i}.ydata(:,2:3),'o');
  t = linspace(0,10); % time for plots
  th = results.theta; % the whole parameter vector, including not sampled
  th(results.parind) = mcmean; % set sampled components
  thi = th(results.local==0|results.local==i); % local parameters
  [t,y] = ode45(@boxoODE,t,thi(end-1:end),[],thi);
  hold on; plot(t,y); hold off
  title(sprintf('batch %d',i));
end
```

### Preditive plots

We need to augment the time in ydata to get more plot points datamerge in the toolbox does this.

```
time = linspace(0,9,50)'; % new time vector
data{1}.ydata = datamerge(data{1}.ydata,time);
data{2}.ydata = datamerge(data{2}.ydata,time);
```

Function mcmcpred calculates the predictive plots, and mcmcpredplot does the plot.

```
out = mcmcpred(results,chain,[],data,@boxoM,500);
h = mcmcpredplot(out,data,1);
```

 

Published with MATLAB® R2018b
