## Supplementary material for "Quantifying non-communicable diseases’ burden in Egypt using State-Space model": S1 File: boxoM.html

```
function ymod=boxoM(data,theta)
% model function for the boxo example

% starting concentrations are at the end of the parameter vector
y0 = theta(end-1:end);
% time is the first column of data.ydata
t  = data.ydata(:,1);

% if using lsode mex, save parameter vector in global variable
global lsode_data
lsode_data = theta;

if exist('lsode_mex') == 3
  % use much faster mex code for ode
  [tout,y] = lsode22('internal',t,y0);
else
  [tout,y] = ode45(@boxoODE,t,y0,[],theta);
end

ymod = y;
```

Published with MATLAB® R2018b
