## Supplementary material for "Quantifying non-communicable diseases’ burden in Egypt using State-Space model": S1 File: boxoODE.html

```
function dy = boxoODE(t,y,theta)
% ode function for the boxo example

A = y(1);
B = y(2);

k1mean = theta(1);
e1     = theta(2);
k2mean = theta(3);
e2     = theta(4);
tmean  = theta(5);
temp   = theta(6);

e1 = e1*1e6;
e2 = e2*1e6;

R  =  8.314;
z  = 1./R *(1./temp-1./tmean);
k1 = k1mean * exp(-e1*z);
k2 = k2mean * exp(-e2*z);

dy =[ - k1 * A ;
        k1 * A - k2 * B];
```

Published with MATLAB® R2018b
