## Supplementary material for "Quantifying non-communicable diseases’ burden in Egypt using State-Space model": S1 File: cauchyex.html

MCMC toolbox » Examples » Cauchy distribution

### MCMC toolbox example

This example targets 10 dimensional Cauchy distribution. Cauchy distribution does not have finite second moment, so the AM method, which calculates chain variance will have troubles. In practice, it gives good results. Here, we use RAM adaptation, which does not use variance information (http://dx.doi.org/10.1007/s11222-011-9269-5).

```
clear model options params

nsimu = 40000;
npar = 10;

model.ssfun    = @(x,d) 2*sum(log(1+x.^2));
options.nsimu  = nsimu;
options.method = 'ram';
for i=1:npar, params{i} = {sprintf('x_{%d}',i), 0}; end

[res,chain] = mcmcrun(model,[],params,options);
```

```
Sampling these parameters:
name   start [min,max] N(mu,s^2)
x_{1}: 0 [-Inf,Inf] N(0,Inf)
x_{2}: 0 [-Inf,Inf] N(0,Inf)
x_{3}: 0 [-Inf,Inf] N(0,Inf)
x_{4}: 0 [-Inf,Inf] N(0,Inf)
x_{5}: 0 [-Inf,Inf] N(0,Inf)
x_{6}: 0 [-Inf,Inf] N(0,Inf)
x_{7}: 0 [-Inf,Inf] N(0,Inf)
x_{8}: 0 [-Inf,Inf] N(0,Inf)
x_{9}: 0 [-Inf,Inf] N(0,Inf)
x_{10}: 0 [-Inf,Inf] N(0,Inf)
```

### Plot the chain

```
iii = 1:min(npar,4); % plot first 4 chain columns

figure(1); clf
mcmcplot(chain,iii,res);
figure(2); clf
mcmcplot(chain,iii,res,'hist');
for i=iii
  subplot(2,2,i)
  xlim([-10,10]);
  xx = linspace(-10,10);
  yy = cauchypf(xx);
  hold on
  plot(xx,yy)
  hold off
end
```

 

Published with MATLAB® R2018b
