## Supplementary material for "Quantifying non-communicable diseases’ burden in Egypt using State-Space model": S1 File: himmelex.html

MCMC toolbox » Examples » Himmelblau

### Himmelblau exercise 9.9

This is exercise 9.9 from David M. Himmelblau, *Process Analysis by Statistical Methods*, Wiley, 1970.

We model the reactions

```
A + B  (k1)-> C + F
A + C  (k2)-> D + F
A + D  (k3)-> E + F
```

The derivatives can be written as

```
dA/dt = -k1 AB - k2 AC - k3 AD
dB/dt = -k1 AB
dC/dt =  k1 AB - k2 AC
dD/dt =          k2 AC - k3 AD
dE/dt =                  k3 AD
```

The system is written in file himmelode.m and the sum of squares function in himmelss.m.

```
clear model data parama options

data.ydata = [
%  Time (min)     [A] (mole/liter)
            0     0.02090
         4.50     0.01540
         8.67     0.01422
        12.67     0.01335
        17.75     0.01232
        22.67     0.01181
        27.08     0.01139
        32.00     0.01092
        36.00     0.01054
        46.33     0.00978
        57.00     0.009157
        69.00     0.008594
        76.75     0.008395
        90.00     0.007891
       102.00     0.007510
       108.00     0.007370
       147.92     0.006646
       198.00     0.005883
       241.75     0.005322
       270.25     0.004960
       326.25     0.004518
       418.00     0.004075
       501.00     0.003715
    ];
```

Initial concentrations are saved in data to be used in sum of squares function.

```
A0 = 0.02090; B0 = A0/3; C0 = 0; D0 = 0; E0 = 0;
data.y0 = [A0;B0;C0;D0;E0];
```

Refine the first guess for the parameters with fminseacrh and calculate residual variance as an estimate of the model error variance.

```
k00 = [15,1.5,0.3]';
[k0,ss0] = fminsearch(@himmelss,k00,[],data)
mse = ss0/(length(data.ydata)-4);
```

```
k0 =

       14.402
       1.5663
      0.29042


ss0 =

   4.1564e-07
```

```
params = {
    {'k1', k0(1), 0}
    {'k2', k0(2), 0}
    {'k3', k0(3), 0}
    };

model.ssfun = @himmelss;
model.sigma2 = mse;

options.nsimu = 1000;
options.updatesigma = 1;
```

```
[results,chain,s2chain] = mcmcrun(model,data,params,options);
```

```
Sampling these parameters:
name   start [min,max] N(mu,s^2)
k1: 14.4019 [0,Inf] N(0,Inf)
k2: 1.5663 [0,Inf] N(0,Inf)
k3: 0.290424 [0,Inf] N(0,Inf)
```

```
figure(1); clf
mcmcplot(chain,[],results,'chainpanel')
subplot(2,2,4)
mcmcplot(sqrt(s2chain),[],[],'dens',2)
title('error std')
```

Function chainstats lists some statistics, including the estimated Monte Carlo error of the estimates.

```
chainstats(chain,results)
```

```
MCMC statistics, nsimu = 1000

                 mean         std      MC_err         tau      geweke
---------------------------------------------------------------------
        k1     14.415     0.67092    0.053086       9.598     0.98736
        k2     1.5668    0.039761   0.0039085      11.468     0.99983
        k3    0.29106    0.012697   0.0012202      13.761     0.99665
---------------------------------------------------------------------
```

```
figure(2); clf
[t,y] = ode45(@himmelode,linspace(0,600),data.y0,[],mean(chain));
plot(data.ydata(:,1),data.ydata(:,2),'s',t,y,'-')
ylim([0,0.021])
legend({'Aobs','A','B','C','D','E'},'Location','best')
title('Data and fitted model')
```

Published with MATLAB® R2018b
