## Supplementary material for "Quantifying non-communicable diseases’ burden in Egypt using State-Space model": S1 File: himmelode.html

```
function ydot = himmelode(t,y,k)
% Himmelblau 9.9 odefile
A=y(1); B=y(2); C=y(3); D=y(4);
ydot = [
    -k(1)*A*B - k(2)*A*C - k(3)*A*D;
    -k(1)*A*B;
    +k(1)*A*B - k(2)*A*C;
              + k(2)*A*C - k(3)*A*D;
                         + k(3)*A*D;
       ];
```

Published with MATLAB® R2018b
