## Supplementary material for "Quantifying non-communicable diseases’ burden in Egypt using State-Space model": S1 File: himmelss.html

```
function ss = himmelss(k,data)
% sum-of-squares for Himmelblau 9.9

time = data.ydata(:,1);
Aobs = data.ydata(:,2);
y0   = data.y0;

[t,y] = ode45(@himmelode,time,y0,[],k);
Amodel = y(:,1);

ss = sum((Aobs-Amodel).^2);
```

Published with MATLAB® R2018b
