## Supplementary material for "Quantifying non-communicable diseases’ burden in Egypt using State-Space model": S1 File: monodex.html

MCMC toolbox » Examples » Monod model

### MCMC toolbox examples

This example is from P. M. Berthouex and L. C. Brown: *Statistics for Environmental Engineers*, CRC Press, 2002.

We fit the Monod model

to observations

```
x (mg / L COD):  28    55    83    110   138   225   375
y (1 / h):       0.053 0.060 0.112 0.105 0.099 0.122 0.125
```

First clear some variables from possible previous runs.

```
clear data model options
```

Next, create a data structure for the observations and control variables. Typically one could make a structure data that contains fields xdata and ydata.

```
data.xdata = [28    55    83    110   138   225   375]';   % x (mg / L COD)
data.ydata = [0.053 0.060 0.112 0.105 0.099 0.122 0.125]'; % y (1 / h)
```

Here is a plot of the data set.

```
figure(1); clf
plot(data.xdata,data.ydata,'s');
xlim([0 400]); xlabel('x [mg/L COD]'); ylabel('y [1/h]');
```

For the MCMC run we need the sum of squares function. For the plots we shall also need a function that returns the model. Both the model and the sum of squares functions are easy to write as one line anonymous functions using the @ construct.

```
modelfun = @(x,theta) theta(1)*x./(theta(2)+x);
ssfun    = @(theta,data) sum((data.ydata-modelfun(data.xdata,theta)).^2);
```

In this case the initial values for the parameters are easy to guess by looking at the plotted data. As we alredy have the sum-of-squares function, we might as well try to minimize it using fminsearch.

```
[tmin,ssmin]=fminsearch(ssfun,[0.15;100],[],data)
n = length(data.xdata);
p = 2;
mse = ssmin/(n-p) % estimate for the error variance
```

```
tmin =

      0.14542
       49.053


ssmin =

   0.00081677


mse =

   0.00016335
```

The Jacobian matrix of the model function is easy to calculate so we use it to produce estimate of the covariance of theta. This can be used as the initial proposal covariance for the MCMC samples by option options.qcov below.

```
J = [data.xdata./(tmin(2)+data.xdata), ...
     -tmin(1).*data.xdata./(tmin(2)+data.xdata).^2];
tcov = inv(J'*J)*mse
```

```
tcov =

   0.00024471      0.25011
      0.25011       320.84
```

We have to define three structures for inputs of the mcmcrun function: parameter, model, and options. Parameter structure has a special form and it is constructed as Matlab cell array with curly brackets. At least the structure has, for each parameter, the name of the parameter and the initial value of it. Third optional parameter given below is the minimal accepted value. With it we set a positivity constraits for both of the parameters.

```
params = {
    {'theta1', tmin(1), 0}
    {'theta2', tmin(2), 0}
    };
```

In general, each parameter line can have up to 7 elements: 'name', initial\_value, min\_value, max\_value, prior\_mu, prior\_sigma, and targetflag

The model structure holds information about the model. Minimally we need to set ssfun for the sum of squares function and the initial estimate of the error variance sigma2.

```
model.ssfun  = ssfun;
model.sigma2 = mse; % (initial) error variance from residuals of the lsq fit
```

If we want to sample the error variance sigma2 as an extra model parameter, we need to set a prior distribution for it. A convenient choice is the conjugate inverse chi-squared distribution, which allows Gibbs sampling step for sigma2 after every Metropolis-Hastings update for the other parameters. This is acchieved by options.updatesigma=1, below. The default prior is uninformative, but we can set the prior parameters with the following options. Option model.N for the number of observatios is needed for 'updatesigma', if it is not given, the code tries to guess N from the data.

```
model.N = length(data.ydata);  % total number of observations
model.S20 = model.sigma2;      % prior mean for sigma2
model.N0  = 4;                 % prior accuracy for sigma2
```

The options structure has settings for the MCMC run. We need at least the number of simulations in nsimu. Here we also set the option updatesigma to allow automatic sampling and estimation of the error variance. The option qcov sets the initial covariance for the Gaussian proposal density of the MCMC sampler.

```
options.nsimu = 4000;
options.updatesigma = 1;
options.qcov = tcov; % covariance from the initial fit
```

The actual MCMC simulation run is done using the function mcmcrun.

```
[res,chain,s2chain] = mcmcrun(model,data,params,options);
```

```
Sampling these parameters:
name   start [min,max] N(mu,s^2)
theta1: 0.14542 [0,Inf] N(0,Inf)
theta2: 49.053 [0,Inf] N(0,Inf)
```

During the run, a status window is showing the estimated time to the end of the simulation. The simulation can be ended by Cancel button and the chain generated so far is returned.

After the run the we have a structure res that contains some information about the run and a matrix outputs chain and s2chain that contain the actual MCMC chains for the parameters and for the observation error variance.

The chain variable is nsimu × npar matrix and it can be plotted and manipulated with standard Matlab functions. MCMC toolbox function mcmcplot can be used to make some useful chain plots and also to plot 1 and 2 dimensional marginal kernel density estimates of the posterior distributions.

```
figure(2); clf
mcmcplot(chain,[],res,'chainpanel');
```

The 'pairs' options makes pairwise scatterplots of the columns of the chain.

```
figure(3); clf
mcmcplot(chain,[],res,'pairs');
```

If we take square root of the s2chain we get the chain for error standard deviation. Here we use 'hist' option for the histogram of the chain.

```
figure(4); clf
mcmcplot(sqrt(s2chain),[],[],'hist')
title('Error std posterior')

% add prior distribution to the plot, if it was informative
if res.N0>0
  xl = xlim; xx = linspace(xl(1),xl(2));
  hold on
  plot(xx,invchi1pf(xx,res.N0,sqrt(res.S20)))
  hold off
  legend('posterior','prior')
end
```

A point estimate of the model parameters can be calculated as the mean of the chain. Here we plot the fitted model using the posterior means of the parameters.

```
x = linspace(0,400)';
figure(1)
hold on
plot(x,modelfun(x,mean(chain)),'-k')
hold off
legend({'data','model'},'Location','best')
```

Instead of just a point estimate of the fit, we should also study the predictive posterior distribution of the model. The mcmcpred and mcmcpredplot functions can be used for this purpose. By them we can calculate the model fit for a randomly selected subset of the chain and calculate the predictive envelope of the model. The grey areas in the plot correspond to 50%, 90%, 95%, and 99% posterior regions.

```
figure(5); clf
out = mcmcpred(res,chain,[],x,modelfun);
mcmcpredplot(out);
hold on
plot(data.xdata,data.ydata,'s'); % add data points to the plot
xlabel('x [mg/L COD]'); ylabel('y [1/h]');
hold off
title('Predictive envelopes of the model')
```

Published with MATLAB® R2018b
