## Supplementary material for "Quantifying non-communicable diseases’ burden in Egypt using State-Space model": S1 File: normalex50.html

MCMC toolbox » Examples » 50 dimensional Normal distribution

Test I: 50 dimensional Gaussian

```
clear model options params

nsimu = 100000; % how many simulations
npar = 50;

% generate correlated covariance matrix with increasing variances
s = (1:npar)';
ci = inv(cov2cor(covcond(10,ones(npar,1))).*(s*s'));

model.ssfun      = @(x,d) x(:)'*ci*x(:);
options.nsimu    = nsimu;
options.method   = 'am';
options.qcov     = eye(npar)/npar*2.4^2.;
options.adaptint = 1000;
for i=1:npar, params{i} = {sprintf('x_{%d}',i), 0}; end

[res,chain] = mcmcrun(model,[],params,options);
```

```
Sampling these parameters:
name   start [min,max] N(mu,s^2)
x_{1}: 0 [-Inf,Inf] N(0,Inf)
x_{2}: 0 [-Inf,Inf] N(0,Inf)
x_{3}: 0 [-Inf,Inf] N(0,Inf)
x_{4}: 0 [-Inf,Inf] N(0,Inf)
x_{5}: 0 [-Inf,Inf] N(0,Inf)
x_{6}: 0 [-Inf,Inf] N(0,Inf)
x_{7}: 0 [-Inf,Inf] N(0,Inf)
x_{8}: 0 [-Inf,Inf] N(0,Inf)
x_{9}: 0 [-Inf,Inf] N(0,Inf)
x_{10}: 0 [-Inf,Inf] N(0,Inf)
x_{11}: 0 [-Inf,Inf] N(0,Inf)
x_{12}: 0 [-Inf,Inf] N(0,Inf)
x_{13}: 0 [-Inf,Inf] N(0,Inf)
x_{14}: 0 [-Inf,Inf] N(0,Inf)
x_{15}: 0 [-Inf,Inf] N(0,Inf)
x_{16}: 0 [-Inf,Inf] N(0,Inf)
x_{17}: 0 [-Inf,Inf] N(0,Inf)
x_{18}: 0 [-Inf,Inf] N(0,Inf)
x_{19}: 0 [-Inf,Inf] N(0,Inf)
x_{20}: 0 [-Inf,Inf] N(0,Inf)
x_{21}: 0 [-Inf,Inf] N(0,Inf)
x_{22}: 0 [-Inf,Inf] N(0,Inf)
x_{23}: 0 [-Inf,Inf] N(0,Inf)
x_{24}: 0 [-Inf,Inf] N(0,Inf)
x_{25}: 0 [-Inf,Inf] N(0,Inf)
x_{26}: 0 [-Inf,Inf] N(0,Inf)
x_{27}: 0 [-Inf,Inf] N(0,Inf)
x_{28}: 0 [-Inf,Inf] N(0,Inf)
x_{29}: 0 [-Inf,Inf] N(0,Inf)
x_{30}: 0 [-Inf,Inf] N(0,Inf)
x_{31}: 0 [-Inf,Inf] N(0,Inf)
x_{32}: 0 [-Inf,Inf] N(0,Inf)
x_{33}: 0 [-Inf,Inf] N(0,Inf)
x_{34}: 0 [-Inf,Inf] N(0,Inf)
x_{35}: 0 [-Inf,Inf] N(0,Inf)
x_{36}: 0 [-Inf,Inf] N(0,Inf)
x_{37}: 0 [-Inf,Inf] N(0,Inf)
x_{38}: 0 [-Inf,Inf] N(0,Inf)
x_{39}: 0 [-Inf,Inf] N(0,Inf)
x_{40}: 0 [-Inf,Inf] N(0,Inf)
...
```

```
iii = 1:12;
figure(1); clf; mcmcplot(chain,iii,res);
figure(2); clf; mcmcplot(chain,iii,res,'hist',20);
figure(3); clf
cummean = @(x) cumsum(x(:))./(1:length(x))';
cumstd  = @(x) sqrt(cummean(x.^2) - cummean(x).^2);
plot(1:options.nsimu,cummean(chain(:,1)),'-')
hold on
plot(1:options.nsimu,cumstd(chain(:,1)),'-')
plot(1:options.nsimu,cumstd(chain(:,5)),'-')
hold off
grid
legend({'mean of x_1','std of x_1','std of x_5'},'location','best')
title('Convergence for 50 dimensional Gaussian distribution')
```

  

Published with MATLAB® R2018b
