## Supplementary material for "Quantifying non-communicable diseases’ burden in Egypt using State-Space model": S1 File: normalex.html

MCMC toolbox » Examples » Normal distribution

### MCMC toolbox example

In this example, we generate Gaussian target with known covariance matrix. The target distribution has a known form and could be calculated analytically, so this simulation is mainly for testing of the algorithms. We study the correctness of the chain by calculating points that lie inside 50% and 95% probability contours.

```
clear model data params options

nsimu = 10000;  % number of simulations
npar  = 100;     % dimension of the unknown

data.x0 = zeros(1,npar); % mean vector
```

We create the needed prameter structure in a loop

```
for i=1:npar
  %            'name'                     initial
  params{i} = {sprintf('\\theta_{%d}',i), data.x0(i)};
end
```

Create covariance and precision matrises. Function covcond creates a covariance matrix with given condition number and a direction of the first principal axis. It returns the covariance matrix and its inverse, the precision matrix.

```
[Sig,Lam] = covcond(100,ones(npar,1));
```

Store the precision matrix in data structure so we can use it in the ssfun.

```
data.Lam = Lam;
```

The function ssfun for mcmcrun is the quadratic form in the Gaussian distribution.

```
model.ssfun     = @(x,data) (x-data.x0)*data.Lam*(x-data.x0)';
model.N         = 1;
```

For mcmcrun we use scaled version of the known target covariance as the proposal covariance. This scaling is known to produce an optimal proposal.

```
options.nsimu   = nsimu;
options.qcov    = 2.4^2/npar*Sig;
options.method  = 'ram';  % use the (default) DRAM method
options.verbosity = 1;
```

Generate the MCMC chain.

```
[results,chain] = mcmcrun(model,data,params,options);
```

```
Setting nbatch to 1
Sampling these parameters:
name   start [min,max] N(mu,s^2)
\theta_{1}: 0 [-Inf,Inf] N(0,Inf)
\theta_{2}: 0 [-Inf,Inf] N(0,Inf)
\theta_{3}: 0 [-Inf,Inf] N(0,Inf)
\theta_{4}: 0 [-Inf,Inf] N(0,Inf)
\theta_{5}: 0 [-Inf,Inf] N(0,Inf)
\theta_{6}: 0 [-Inf,Inf] N(0,Inf)
\theta_{7}: 0 [-Inf,Inf] N(0,Inf)
\theta_{8}: 0 [-Inf,Inf] N(0,Inf)
\theta_{9}: 0 [-Inf,Inf] N(0,Inf)
\theta_{10}: 0 [-Inf,Inf] N(0,Inf)
\theta_{11}: 0 [-Inf,Inf] N(0,Inf)
\theta_{12}: 0 [-Inf,Inf] N(0,Inf)
\theta_{13}: 0 [-Inf,Inf] N(0,Inf)
\theta_{14}: 0 [-Inf,Inf] N(0,Inf)
\theta_{15}: 0 [-Inf,Inf] N(0,Inf)
\theta_{16}: 0 [-Inf,Inf] N(0,Inf)
\theta_{17}: 0 [-Inf,Inf] N(0,Inf)
\theta_{18}: 0 [-Inf,Inf] N(0,Inf)
\theta_{19}: 0 [-Inf,Inf] N(0,Inf)
\theta_{20}: 0 [-Inf,Inf] N(0,Inf)
\theta_{21}: 0 [-Inf,Inf] N(0,Inf)
\theta_{22}: 0 [-Inf,Inf] N(0,Inf)
\theta_{23}: 0 [-Inf,Inf] N(0,Inf)
\theta_{24}: 0 [-Inf,Inf] N(0,Inf)
\theta_{25}: 0 [-Inf,Inf] N(0,Inf)
\theta_{26}: 0 [-Inf,Inf] N(0,Inf)
\theta_{27}: 0 [-Inf,Inf] N(0,Inf)
\theta_{28}: 0 [-Inf,Inf] N(0,Inf)
\theta_{29}: 0 [-Inf,Inf] N(0,Inf)
\theta_{30}: 0 [-Inf,Inf] N(0,Inf)
\theta_{31}: 0 [-Inf,Inf] N(0,Inf)
\theta_{32}: 0 [-Inf,Inf] N(0,Inf)
\theta_{33}: 0 [-Inf,Inf] N(0,Inf)
\theta_{34}: 0 [-Inf,Inf] N(0,Inf)
\theta_{35}: 0 [-Inf,Inf] N(0,Inf)
\theta_{36}: 0 [-Inf,Inf] N(0,Inf)
\theta_{37}: 0 [-Inf,Inf] N(0,Inf)
\theta_{38}: 0 [-Inf,Inf] N(0,Inf)
\theta_{39}: 0 [-Inf,Inf] N(0,Inf)
\theta_{40}: 0 [-Inf,Inf] N(0,Inf)
...
```

```
figure(1); clf
mcmcplot(chain,[1:4],results.names,'chainpanel')
```

From the generated chain we calculate the relative distances of the chain points from the origin and count the points that are inside given probability limits. Then we plot the first two dimensions of the chain together with the correct probability contours.

The title of the 2d plot shows the rejection rate and the propotion of points inside the ellipsoids. Number tau\*t in the title tells how many seconds it takes to generate 1000 independent samples according to the integrated autocorrelation time (iact) estimate.

```
d    = mahalanobis(chain(:,1:npar),data.x0,Lam,1);
c50  = chiqf(0.50,npar);
c95  = chiqf(0.95,npar);
cc50 = sum(d<c50)./nsimu;
cc95 = sum(d<c95)./nsimu;

figure(2); clf
mcmcplot(chain,[1,2],results.names,'pairs',0)

title(sprintf('Rejected = %.1f%%, c50 = %.1f%%, c95 = %.1f%%, \\tau*t=%.1f', ...
              results.rejected*100, cc50*100, cc95*100, ...
              results.simutime/results.nsimu*1000*mean(iact(chain))))

c50  = chiqf(0.50,2);
c95  = chiqf(0.95,2);

hold on
ellipse(data.x0(1:2),c50*Sig(1:2,1:2),'r--','LineWidth',2);
ellipse(data.x0(1:2),c95*Sig(1:2,1:2),'r-','LineWidth',2);
axis equal
hold off
```

Published with MATLAB® R2018b
