## Supplementary material for "Quantifying non-communicable diseases’ burden in Egypt using State-Space model": S1 File: threemodesex.html

MCMC toolbox » Examples » Three modes example

Metropolis-Hastings MCMC may have troubles if the target distributio has multiple modes. Here we test with a target, which is 4 dimensional mixed Gaussian with 3 distinct modes. By changing the scaling of DRAM, we find all the modes of this example.

```
clear model options params

nsimu = 50000; % how many simulations
npar = 4;
mu1  = [-8,-8,-8,-8];
mu2  = [ 0, 0, 0, 0];
mu3  = [ 8, 8, 8, 8];
sigs = [ 1, 1, 1, 1];
w    = [0.1, 0.3, 0.6]; % 3 mixture weights

model.ssfun= @(x,d) -2*log(w(1)*mvnorpf(x,mu1,sigs)+w(2)*mvnorpf(x,mu2,sigs)+w(3)*mvnorpf(x,mu3,sigs));

% DRAM, first large, then small
options.nsimu    = nsimu;
options.method   = 'dram';
options.qcov     = eye(npar)*5^2;
options.drscale  = 5;
options.adascale = 2.4 / sqrt(npar) * 5;
for i=1:npar, params{i} = {sprintf('x_{%d}',i), 0}; end

[res,chain] = mcmcrun(model,[],params,options);
```

```
Sampling these parameters:
name   start [min,max] N(mu,s^2)
x_{1}: 0 [-Inf,Inf] N(0,Inf)
x_{2}: 0 [-Inf,Inf] N(0,Inf)
x_{3}: 0 [-Inf,Inf] N(0,Inf)
x_{4}: 0 [-Inf,Inf] N(0,Inf)
```

```
figure(1); clf; mcmcplot(chain,[],res,'chainpanel')
figure(2); clf; mcmcplot(chain,[],res,'denspanel')
figure(3); clf
mcmcplot(chain,[1],res,'hist')
xx = linspace(-14,14,200)';
yy = w(1)*norpf(xx,mu1(1),sigs(1)) + w(2)*norpf(xx,mu2(1),sigs(1)) + w(3)*norpf(xx,mu3(1),sigs(1));
hold on
plot(xx,yy,'-')
hold off
xlabel('x_1')
title('3 modes in 4 dimensional Gaussian mixtures')
```

  

Published with MATLAB® R2018b
