## Supplementary material for "Quantifying non-communicable diseases’ burden in Egypt using State-Space model": S1 File: examples.html

xml version="1.0" encoding="utf-8"?


MCMC toolbox for Matlab - Examples


UP 
|
 HOME

### MCMC toolbox for Matlab - Examples

These examples are all Matlab scripts and the web pages are generated using the `publish` function in Matlab. This collection of examples is a part of the mcmcstat source code, in the `examples` sub directory. They use the MCMC toolbox, only.

Monod model
:   Fitting two dimensional Monod model for bacterial growth.

Dose response
:   The classical beetle data is analysed using logistic regression and MCMC.

Algae model
:   Simple dynamic ODE model describes a nutirition food web in a lake.

Chemical kinetics model 1
:   Simple chemical kinetics example with 3 model parameters.

Chemical kinetics model 2
:   This example uses some advanced features of the toolbox. The data is made of two batches and some of the unknowns are local to the batches.

Normal
:   Sample from a fixed multi dimensional Gaussian distribution to test the MCMC algorithm.

Banana
:   Another technical example to check that the method works also with non Gaussian target distributions

These three examples were motivated by the interactive discussion in
https://www.biogeosciences.net/14/4295/2017/bg-14-4295-2017-discussion.html

50 d Gaussian
:   50 dimensional Gaussian target.

Cauchy distribution
:   10 dimensional Cauchy distribution.

3 modes
:   4 dimensional mixed Gaussian with 3 distinct modes.

---

Author: Marko Laine

Created: 2018-11-19 Ma 16:45
