## Supplementary material for "Quantifying non-communicable diseases’ burden in Egypt using State-Space model": S1 File: examples.org

#+TITLE:     MCMC toolbox for Matlab - Examples
#+AUTHOR:    Marko Laine
#+DESCRIPTION: Matlab toolbox for Markov chain Monte Carlo
#+KEYWORDS: MCMC, Matlab, adaptive mcmc
#+LANGUAGE:  en
#+HTML_LINK_UP: ..
# # +HTML_LINK_HOME: http://helios.fmi.fi/~lainema/mcmc/
#+OPTIONS: num:nil toc:nil ^:{} creator:nil timestamp:t author:t
#+HTML_HEAD: <link rel="stylesheet" type="text/css" href="style.css" />
#+MACRO: helploc http://www.mathworks.com/access/helpdesk/help/techdoc/ref/$1.shtml
#+LINK: matlab  http://www.mathworks.com/access/helpdesk/help/techdoc/ref/%s.shtml


These examples are all Matlab scripts and the web pages are generated using the [[matlab:publish][=publish=]] function in Matlab. This collection of examples is a part of the mcmcstat source code, in the =examples= sub directory. They use the MCMC toolbox, only.

- [[file:ex/monodex.html][Monod model]] :: Fitting two dimensional Monod model for bacterial  growth.
- [[file:ex/beetleex.html][Dose response]] :: The classical beetle data is analysed using logistic regression and MCMC.
- [[file:ex/algaeex.html][Algae model]] :: Simple dynamic ODE model describes a nutirition food web in a lake.
- [[file:ex/himmelex.html][Chemical kinetics model 1]] :: Simple chemical kinetics example with 3 model parameters.
- [[file:ex/boxoex.html][Chemical kinetics model 2]] :: This example uses some advanced features of the toolbox. The data is made of two batches and some of the unknowns are local to the batches.
- [[file:ex/normalex.html][Normal]] :: Sample from a fixed multi dimensional Gaussian distribution to test the MCMC algorithm.
- [[file:ex/bananaex.html][Banana]] :: Another technical example to check that the method works also with non Gaussian target distributions


These three examples were motivated by the interactive discussion in 
https://www.biogeosciences.net/14/4295/2017/bg-14-4295-2017-discussion.html

- [[file:ex/normalex50.html][50 d Gaussian]] :: 50 dimensional Gaussian target.
- [[file:ex/cauchyex.html][Cauchy distribution]] :: 10 dimensional Cauchy distribution.
- [[file:ex/threemodesex.html][3 modes]] :: 4 dimensional mixed Gaussian with 3 distinct modes.

#
#+HTML: <hr>

# Local Variables:
# coding: utf-8
# mode: org
# eval: (flyspell-mode 1)
# eval: (visual-line-mode 1)
# eval: (auto-fill-mode -1)
# ispell-dictionary: "english"
# End:
