## Supplementary material for "Quantifying non-communicable diseases’ burden in Egypt using State-Space model": S1 File: index.html

xml version="1.0" encoding="utf-8"?


MCMC toolbox for Matlab


### MCMC toolbox for Matlab

#### Table of Contents

- Introduction
- Examples
- Code
- Downloads
- References

#### Introduction

The MCMCSTAT Matlab package contains a set of Matlab functions for some Bayesian analyses of mathematical models by Markov chain Monte Carlo simulation. This code might be useful to you if you are already familiar with Matlab and want to do MCMC analysis using it.

For a more comprehensive and better documented and maintained software for MCMC, see, e.g. Stan. There are some MCMC functions in Mahtworks own Statistics Toolbox, too.

This toolbox provides tools to generate and analyse Metropolis-Hastings MCMC chains using multivariate Gaussian proposal distribution. The covariance matrix of the proposal distribution can be adapted during the simulation according to adaptive schemes described in the references.

The code can do the following

- Produce MCMC chain for user-written -2\*log(likelihood) and -2\*log(prior) functions. These will be equal to sum-of-squares cost functions when using Gaussian likelihood and prior.
- In case of Gaussian error model, sample the model error variance from the conjugate inverse chi-squared distribution.
- Do plots and statistical analyses based on the chain, such as basic statistics, convergence diagnostics, chain timeseries plots, 2 dimensional clouds of points, kernel densities, and histograms.
- Calculate densities, cumulative distributions, quantiles, and random variates for some useful common statistical distributions without using Mathworks own statistics toolbox.

The code is self consistent, no additional Matlab toolboxes are used. However, a quite recent version of Matlab is needed. The documentation might get better gradually, but in the mean while please look at the following page:

#### Examples

- Examples on using the toolbox for some statistical problems.

#### Code

The main functions in the toolbox are the following.

`mcmcrun.m`
:   Matlab function for the MCMC run. The user provides
    her own Matlab function to calculate the
    "sum-of-squares" function for the likelihood part,
    e.g. a function that calculates minus twice the log
    likelihood, -2log(p(θ;data)). Optionally a prior
    "sum-of-squares" function can also be given,
    returning -2log(p(θ)).
    See the example and help mcmcrun for more details.

`mcmcplot.m`
:   This function makes some useful plots of the
    generated chain, such as chain time series, 2
    dimensional marginal plots, kernel density
    estimates, and histograms. See help mcmcplot.

`mcmcpred.m`
:   For certain types of models is is useful to plot
    predictive envelopes of model functions by sampling
    parameter values from the generated chain. This
    functions calls the model function repeatedly while
    sampling the unknowns from the chain. It calculates
    probability regions with respect to a given "time"
    variable of the model. See the examples.

#### Downloads

The full Matlab code and this documentation are available from GitHub.

To install the toolbox clone the folder `mcmcstat` to a suitable directory and then add Matlab path to that directory.

```
git clone https://github.com/mjlaine/mcmcstat.git
```

Alternatively, you can download the whole repository as a zip file from the GitHub page or directly as
https://github.com/mjlaine/mcmcstat/archive/master.zip

Comments are welcome!

---

Author: Marko Laine

Created: 2018-11-19 Ma 13:33
