## Supplementary material for "Quantifying non-communicable diseases’ burden in Egypt using State-Space model": S1 File: mcmcplot.html

xml version="1.0" encoding="utf-8"?


mcmcplot.m


UP 
|
 HOME

### mcmcplot.m

```
%MCMCPLOT Plot mcmc chain
% mcmcplot(chain,inds,names,plottype, ...)
% plot several plots from mcmc chain
%  chain    - mcmc chain matrix
%  inds     - columns to use, default all columns
%  names    - names of the columns of chain as char array
%  plottype - 'chain', 'chainpanel', 'pairs', 'dens', 'denspanel', 'acf', 'hist'
%   'pairs':  mcmcplot(...,'pairs',smo,rho)
%             SMO is the bandwidth parameter for 2D kernel estimation: smoother
%             density curves with big smo (if smo not defined, no density
%             curves plotted)
%             RHO is the correlation for the kernel estimator (if not
%             defined, calculated from the chain)
%   'dens':   mcmcplot(...,'dens',smo)
%             SMO is the bandwidth for 1D kernel estimation (default: 1)
%   'hist':   mcmcplot(...,'hist',nbin,dens)
%             NBIN is the number of bins used for the histograms
%             DENS ('normal', 'lognor', or 'kernel') adds estimated density
%             on top of the histogram
%   'chain':  mcmcplot(...,'chain',maxpoints)
%             MAXPOINTS max n:o of points in the chain plot
%   'acf':    no optional parameters
```

Created: 2018-11-19 Ma 13:54
