## Supplementary material for "Quantifying non-communicable diseases’ burden in Egypt using State-Space model": S1 File: mcmcplot.org

#+TITLE:     mcmcplot.m
#+AUTHOR:    Marko Laine
#+HTML_LINK_HOME: ..
#+HTML_LINK_UP: ..
#+OPTIONS: num:nil toc:nil ^:{} creator:nil timestamp:t author:nil
#+HTML_HEAD: <link rel="stylesheet" type="text/css" href="style.css" />

#+INCLUDE: "../mcmcplot.m" src matlab :lines "2-24"
