## Supplementary material for "Quantifying non-communicable diseases’ burden in Egypt using State-Space model": S1 File: mcmcrun.html

xml version="1.0" encoding="utf-8"?


mcmcrun.m


UP 
|
 HOME

### mcmcrun.m

```
%MCMCRUN Metropolis-Hastings MCMC simulation for nonlinear Gaussian models
% properties:
%  multiple y-columns, sigma2-sampling, adaptation,
%  Gaussian prior, parameter limits, delayed rejection, dram
%
% [RESULTS,CHAIN,S2CHAIN,SSCHAIN] = MCMCRUN(MODEL,DATA,PARAMS,OPTIONS)
% MODEL   model options structure
%    model.ssfun    -2*log(likelihood) function
%    model.priorfun -2*log(pior) prior function
%    model.sigma2   initial error variance
%    model.N        total number of observations
%    model.S20      prior for sigma2
%    model.N0       prior accuracy for S20
%    model.nbatch   number of datasets
%
%     sum-of-squares function 'model.ssfun' is called as
%     ss = ssfun(par,data) or
%     ss = ssfun(par,data,local)
%     instead of ssfun, you can use model.modelfun as
%     ymodel = modelfun(data{ibatch},theta_local)
%
%     prior function is called as priorfun(par,pri_mu,pri_sig) it
%     defaults to Gaussian prior with infinite variance
%
%     The parameter sigma2 gives the variances of measured components,
%     one for each. If the default options.updatesigma = 0 (see below) is
%     used, sigma2 is fixed, as typically estimated from the fitted residuals.
%     If opions.updatesigma = 1, the variances are sampled as conjugate priors
%     specified by the parameters S20 and N0 of the inverse gamma
%     distribution, with the 'noninformative' defaults
%          S20 = sigma2   (as given by the user)
%          N0  = 1
%     Larger values of N0 limit the samples closer to S20
%     (see,e.g., A.Gelman et all:
%     Bayesian Data Analysis, http://www.stat.columbia.edu/~gelman/book/)
%
% DATA the data, passed directly to ssfun. The structure of DATA is given
%      by the user. Typically, it contains the measurements
%
%      data.xdata
%      data.ydata,
%
%      A possible 'time' variable must be given in the first column of
%      xdata. Note that only data.xdata is needed for model simulations.
%      In addition, DATA may include any user defined structure needed by
%      |modelfun| or |ssfun|
%
% PARAMS  theta structure
%   {  {'par1',initial, min, max, pri_mu, pri_sig, targetflag, localflag}
%      {'par2',initial, min, max, pri_mu, pri_sig, targetflag, localflag}
%      ... }
%
%   'name' and initial are compulsary, other values default to
%   {'name', initial,  -Inf, Inf,  NaN, Inf,  1,  0}
%
% OPTIONS mcmc run options
%    options.nsimu            number of simulations
%    options.qcov             proposal covariance
%    options.method           'dram','am','dr', 'ram' or 'mh'
%    options.adaptint         interval for adaptation, if 'dram' or 'am' used
%                             DEFAULT adaptint = 100
%    options.drscale          scaling for proposal stages of dr
%                             DEFAULT 3 stages, drscale = [5 4 3]
%    options.updatesigma      update error variance. Sigma2 sampled with updatesigma=1
%                             DEFAULT updatesigma=0
%    options.verbosity        level of information printed
%    options.waitbar          use graphical waitbar?
%    options.burnintime       burn in before adaptation starts
%
% Output:
%  RESULTS   structure that contains results and information about
%            the simulations
%  CHAIN, S2CHAIN, SSCHAIN
```

Created: 2018-11-19 Ma 13:55
