## Supplementary material for "Quantifying non-communicable diseases’ burden in Egypt using State-Space model": S1 File: elapsed.m

```
function y=elapsed(t)
%ELAPSED  Time elapsed as a string since t or since last tic


% Marko Laine <>
% $Revision: 1.4 $  $Date: 2012/09/27 11:47:35 $

if nargin<1
   t=toc;
end
secs = t;
mins = floor(secs/60);
secs = floor(secs - 60*mins);
hrs  = floor(mins/60);
mins = mins - hrs*60;
  e=sprintf('%g:%02g:%02g',hrs,mins,secs);
if nargout>0
  y=e;
else
  disp(e)
end
```
