## Supplementary material for "Quantifying non-communicable diseases’ burden in Egypt using State-Space model": S1 File: ellipse.m

```
function [o1,o2]=ellipse(mu,cmat,varargin)
%ELLIPSE Draws a probability contour ellipse
% ellipse(mu,cmat) draws an ellipse centered at mu and with
% covariance matrix cmat.
% h = ellipse(...) plots and returns handle to the ellipse line
% [x,y] = ellipse(...) returns ellipse points (no plot)
% use chiqf(0.9,2)*cmat to get 90% Gaussian probability region
% example:
% plot(x(:,1),x(:,2),'.'); % plot data points
% hold on; ellipse(xmean, chiqf(0.9,2)*xcov); hold off

% Marko Laine <>
% $Revision: 1.7 $  $Date: 2012/09/27 11:47:35 $

if size(cmat)~=[2 2]
  error('covmat must be 2x2');
end
if cmat~=cmat'
  error('covmat must be symmetric');
end

t = linspace(0,2*pi)';
R = chol(cmat);
x = mu(1) + R(1,1).*cos(t); 
y = mu(2) + R(1,2).*cos(t) + R(2,2).*sin(t);

if nargout<2
  hh = plot(x,y,varargin{:});
else
  o1 = x;
  o2 = y;
end

if nargout==1
  o1=hh;
end
```
