## Supplementary material for "Quantifying non-communicable diseases’ burden in Egypt using State-Space model": S1 File: epsfile.m

```
function epsfile(file,stretch,varargin)
%EPSFILE  dumps current image into eps file
% epsfile(filename)
% epsfile(filename,1) sets 'PaperPositionMode' to 'auto'

% $Revision: 1.4 $  $Date: 2014/10/09 10:51:01 $

if length(file)==0
  error 'usage epsfile file'
end
if nargin > 1 & stretch~=0
  set(gcf,'PaperPositionMode','auto');
end
print('-depsc','-noui',varargin{:},file)
```
