## Supplementary material for "Quantifying non-communicable diseases’ burden in Egypt using State-Space model": S1 File: boxoSS.m

```
function ss=boxoSS(theta,data,local)
% sum-of-square function for the boxo example

nbatch = length(data);
% cumulate the sum-of-squares over the data sets (batches)

ss = 0;
for i=1:nbatch
  
  th     = theta(local==0|local==i);
  tspan  = data{i}.ydata(:,1);
  ydata  = data{i}.ydata(:,2:3);
  ymodel = boxoM(data{i},th);

  ss = ss + sum((ydata - ymodel).^2);
  
end
```
