## Supplementary material for "Quantifying non-communicable diseases’ burden in Egypt using State-Space model": S1 File: puboptions.m

```
% publish options
popts = struct();
popts.format = 'html';
popts.outputDir = '../docs/ex/';
popts.imageFormat = 'png';
popts.stylesheet = '../docs/mxdom2simplehtml.xsl';

popts2 = popts;
popts2.evalCode = false;

pub = @(file) publish(file,popts);
pub2 = @(file) publish(file,popts2);
```
