## Supplementary material for "Quantifying non-communicable diseases’ burden in Egypt using State-Space model": S1 File: expdf.m

```
function y=expdf(x,a)
%EXPDF Exponential cumulative distribution function
% expdf(x,a)

% Marko Laine <>
% $Revision: 1.4 $  $Date: 2012/09/27 11:47:35 $

if nargin<2,a=1;end
y=1-exp(-a.*x);
```
