## Supplementary material for "Quantifying non-communicable diseases’ burden in Egypt using State-Space model": S1 File: expqf.m

```
function p=expqf(x,a);
%EXPQF   Inverse of exponential distribution function 

% Marko Laine <>
% $Revision: 1.4 $  $Date: 2012/09/27 11:47:35 $

if nargin<2,a=1;end

p = -log(1-x)./a;
```
