## Supplementary material for "Quantifying non-communicable diseases’ burden in Egypt using State-Space model": S1 File: extpf.m

```
function y=extpf(x,a,b)
%EXTPF Extreme value probability density function

% Marko Laine <>
% $Revision: 1.2 $  $Date: 2012/09/27 11:47:36 $

if nargin<2,a=0;end
if nargin<3,b=1;end

z = (x-a)./b;
y = exp(-z).*exp(-exp(-z))./b;
```
