## Supplementary material for "Quantifying non-communicable diseases’ burden in Egypt using State-Space model": S1 File: extr.m

```
function y=extr(m,n,a,b)
%EXTR Extreme value random numbers

% Marko Laine <>
% $Revision: 1.2 $  $Date: 2012/09/27 11:47:36 $

if nargin<3,a=0;end
if nargin<4,b=1;end

y = -log(-log(rand(m,n))).*b+a;
```
