## Supplementary material for "Quantifying non-communicable diseases’ burden in Egypt using State-Space model": S1 File: fdf.m

```
function p=fdf(x,df1,df2,nc)
%FDF     F cumulative distribution function
% p=FDF(x,df1,df2)   x quantile, df1, df2 degrees of freedoms 

% Marko Laine <>
% $Revision: 1.8 $  $Date: 2012/09/27 11:47:36 $

if nargin<4 | nc <= 0
  if ~isunix && exist('distribs') == 3
    p = distribs('fdf',x,df1,df2);
  else
    p = betainc(x.*df1./(df2+df1.*x),0.5*df1,0.5*df2);
  end
else
   p = distribs('fdfnc',x,df1,df2,nc);
   % ncbeta.f is not working properly!
   if nc >= 54
     p = 0.0;
   end
end
```
