## Supplementary material for "Quantifying non-communicable diseases’ burden in Egypt using State-Space model": S1 File: fillyy.m

```
function out=fillyy(x,y1,y2,col)
%FILLYY  Fills space between lines
% fillyy(x,y1,y2,col) fill space between lines (x,y1) and (x,y2)
%  with color col

% Marko Laine <>
% $Revision: 1.3 $  $Date: 2012/09/27 11:47:36 $

if nargin < 4
   col='red';
end

x  = x(:)';
y1 = y1(:)';
y2 = y2(:)';
n   = length(x);
X = [ x(1),  x,  x(n),  fliplr(x)  ];
Y = [ y1(1), y2, y1(n), fliplr(y1) ];
h=fill(X,Y,col,'Linestyle','none');
if nargout>0
  out=h;
end
```
