## Supplementary material for "Quantifying non-communicable diseases’ burden in Egypt using State-Space model": S1 File: fpf.m

```
function y=fpf(x,df1,df2)
%FPF     F probability density function
% FPF(x,df1,df2) x quantile, df1, df2 degrees of freedoms

% Marko Laine <>
% $Revision: 1.5 $  $Date: 2012/09/27 11:47:36 $

m2 = df1/2;
n2 = df2/2;
mn = df1/df2;
mn2= m2+n2;
c = exp(gammaln(mn2)-gammaln(m2)-gammaln(n2)) * mn^m2;
y = c * (x.^(m2-1)) .* ((1 + mn*x).^(-mn2));
```
