## Supplementary material for "Quantifying non-communicable diseases’ burden in Egypt using State-Space model": S1 File: fqf.m

```
function x=fqf(p,df1,df2)
%FQF     F quantile function
% x=FQF(p,df1,df2), p probability, df1, df2 degrees of freedoms 

% Marko Laine <>
% $Revision: 1.6 $  $Date: 2012/09/27 11:47:36 $

if exist('distribs') == 3 % we have the mex code
  x=distribs('fqf',p,df1,df2);
else
  % non-mex version
  x = (df2./invbetainc(1-p,df2./2,df1./2) - df2)./df1;
end
```
