## Supplementary material for "Quantifying non-communicable diseases’ burden in Egypt using State-Space model": S1 File: fr.m

```
function x=fr(m,n,df1,df2)
%FR  Random numbers from the F distribution
% FR(m,n,df1,df2) m,n shape of the result, df1, df2 dergees of freedom

% Marko Laine <>
% $Revision: 1.2 $  $Date: 2012/09/27 11:47:36 $

x = (chir(m,n,df1)./df1)./(chir(m,n,df2)./df2);
```
