## Supplementary material for "Quantifying non-communicable diseases’ burden in Egypt using State-Space model": S1 File: gammadf.m

```
function y=gammadf(x,a,b)
% GAMMADF Gamma cumulative distribution function

% Marko Laine <>
% $Revision: 1.6 $  $Date: 2013/09/05 14:00:13 $
if nargin<3, b=1; end
%if exist('distribs') == 3 % mex version
%  y = distribs('gammadf',x./b,a);
%else
y = gammainc(x./b,a);
%end
```
