## Supplementary material for "Quantifying non-communicable diseases’ burden in Egypt using State-Space model": S1 File: gammapf.m

```
function y=gammapf(x,a,b)
% GAMMAPF - gamma probability density function
% GAMMAPF(x,a,b) x quantile, a shape, b scale

% huom parametrisointi

% Marko Laine <>
% $Revision: 1.4 $  $Date: 2012/09/27 11:47:36 $

if nargin<2, a=1; end
if nargin<3, b=1; end

%y = b.^(-a) ./ gamma(a) .* x.^(a-1) .* exp(-x./b);
y = exp(-a*log(b)-gammaln(a)+(a-1)*log(x)-x./b);
```
