## Supplementary material for "Quantifying non-communicable diseases’ burden in Egypt using State-Space model": S1 File: gammaqf.m

```
function y=gammaqf(p,a,b)
% GAMMAQF inverse of Gamma distribution function
% GAMMAQF(p,alpha,beta)

% Marko Laine <>
% $Revision: 1.5 $  $Date: 2012/09/27 11:47:36 $
if nargin<3, b=1; end

if ~isunix & exist('distribs') == 3 % mex file version
  y = distribs('gammain',p,a).*b;
  return
end

y=zeros(size(p));
for i=1:prod(size(y))
  y(i) = gammaqf_m(p(i),a,b);
end

function y=gammaqf_m(p,a,b)
% simple m code version using fzero if mex-version is not available

if p<=0, y=0; return; end
if p>=1, y=Inf; return; end
if nargin<3, b=1; end

% initial quess
if p < 0.05
  x0 = exp((gammaln(a) + log(p))/a);  
elseif p > 0.95
  x0 = -log(-p+1) + gammaln(a);
else    
  xg = sqrt(2)*erfinv(2*p-1);
  if xg < -sqrt(a)
    x0=a;
  else
    x0=sqrt(a)*xg+a;
  end
end

% zfun(0) = p, so find x0 such that zfun(x0) < 0
h = 1;
while zfun(x0,p,a,b)>=0
  x0 = x0 + h;
  h  = h*1.2;
end

% use fzero for zero
y = fzero(@zfun,[0,x0],optimset('tolx',1e-6,'display','none'),p,a,b);

function z = zfun(x,p,a,b)
% zero function for gamaqf
z = p-gammainc(x./b,a);
```
