## Supplementary material for "Quantifying non-communicable diseases’ burden in Egypt using State-Space model": S1 File: gammar.m

```
function y=gammar(m,n,a,b)
%GAMMAR random deviates from gamma distribution
%  GAMMAR(M,N,A,B) returns a M*N matrix of random deviates from the Gamma
%  distribution with shape parameter A and scale parameter B:
%
%  p(x|A,B) = B^-A/gamma(A)*x^(A-1)*exp(-x/B)

% Marko Laine <>
% $Revision: 1.6 $  $Date: 2012/09/27 11:47:36 $
if nargin<4, b=1; end

if a<=0 % special case of a<=0
  y = zeros(m,n);
  return;
end

if exist('distribs') == 3 % mex version
  y = distribs('gammar',m,n,a).*b;
else
  y = gammar_mt(m,n,a,b);
end

function y=gammar_mt(m,n,a,b)
%GAMMAR_MT random deviates from gamma distribution
% 
%  GAMMAR_MT(M,N,A,B) returns a M*N matrix of random deviates from the Gamma
%  distribution with shape parameter A and scale parameter B:
%
%  p(x|A,B) = B^-A/gamma(A)*x^(A-1)*exp(-x/B)
%
%  Uses method of Marsaglia and Tsang (2000)

% G. Marsaglia and W. W. Tsang:
% A Simple Method for Generating Gamma Variables,
% ACM Transactions on Mathematical Software, Vol. 26, No. 3,
% September 2000, 363-372.

if nargin < 4, b=1; end
y = zeros(m,n);
for j=1:n
  for i=1:m
    y(i,j) = gammar_mt1(a,b);
  end
end
%
function y=gammar_mt1(a,b)
if a<1
  y = gammar_mt1(1+a,b)*rand(1)^(1/a);
else
  d = a-1/3;
  c = 1/sqrt(9*d);
  while(1)
    while(1)
      x = randn(1);
      v = 1+c*x;
      if v > 0, break, end
    end
    v = v^3;
    u = rand(1);
    if u < 1-0.0331*x^4, break, end
    if log(u) < 0.5*x^2+d*(1-v+log(v)), break, end
  end
  y = b*d*v;
end
```
