## Supplementary material for "Quantifying non-communicable diseases’ burden in Egypt using State-Space model": S1 File: genperms.m

```
function perms=genperms(N)
% PERMS=GENPERMS(N) generate all permutations
% BTW: there is allready PERMS.M in matlab

% Marko Laine <>
% $Revision: 1.3 $  $Date: 2012/09/27 11:47:36 $

tot   = prod(1:N); % total number of permutations
perms = zeros(tot,N);
perm  = 1:N;
perms(1,:) = perm;
for ind = 2:tot;
   i = N-1;
   while perm(i) >= perm(i+1)
      i=i-1;
   end
   j = N;
   while perm(j) <= perm(i)
      j=j-1;
   end
   x=perm(i); perm(i)=perm(j); perm(j)=x; % swap
   i=i+1; j=N;
   while i < j
      x=perm(i); perm(i)=perm(j); perm(j)=x;
      i=i+1;
      j=j-1;
   end
   perms(ind,:)=perm;
end
```
