## Supplementary material for "Quantifying non-communicable diseases’ burden in Egypt using State-Space model": S1 File: geweke.m

```
function [z,p]=geweke(chain,a,b)
%GEWEKE Geweke's MCMC convergence diagnostic
% [z,p] = geweke(chain,a,b)
% Test for equality of the means of the first a% (default 10%) and
% last b% (50%) of a Markov chain.
% See:
% Stephen P. Brooks and Gareth O. Roberts.
% Assessing convergence of Markov chain Monte Carlo algorithms.
% Statistics and Computing, 8:319--335, 1998.

% ML, 2002
% $Revision: 1.3 $  $Date: 2003/05/07 12:22:19 $

[nsimu,npar]=size(chain);

if nargin<3
  a = 0.1;
  b = 0.5;
end

na = floor(a*nsimu);
nb = nsimu-floor(b*nsimu)+1;

if (na+nb)/nsimu >= 1
  error('Error with na and nb');
end

m1 = mean(chain(1:na,:));
m2 = mean(chain(nb:end,:));

%%% Spectral estimates for variance
sa = spectrum0(chain(1:na,:));
sb = spectrum0(chain(nb:end,:));

z = (m1-m2)./(sqrt(sa/na+sb/(nsimu-nb+1)));
p = 2*(1-nordf(abs(z)));
```
