## Supplementary material for "Quantifying non-communicable diseases’ burden in Egypt using State-Space model": S1 File: gewekeplot.m

```
function zz=gewekeplot(chain)
%GEWEKEPLOT Plot Geweke's diagnostic
% gewekeplot(chain) plots Geweke's diagnostic for increasing number of 
% iterations. See geweke.m

% ML, 2002
% $Revision: 1.3 $  $Date: 2003/05/07 12:22:19 $

[nsimu,npar]=size(chain);

n  = 40;
e  = fix(nsimu/2);
l  = fix(e/n);
ii = 1:l:e;

z = zeros(length(ii),npar);
for i=1:length(ii)
  z(i,:) = geweke(chain(ii(i):end,:));
end

for i=1:npar
  h = subplot(npar,1,i);
  plot(ii,z(:,i));
  set(h,'XLim',[1 e]);
  if i==1
    title('Geweke diagnostics');
  end
  if i==npar
    xlabel('first iteration used');
  end
end  

if nargout > 0
  zz=z;
end
```
