## Supplementary material for "Quantifying non-communicable diseases’ burden in Egypt using State-Space model": S1 File: hist2d.m

```
function [y,xx,yy]=hist2d(x,mxmy,nbin,lam,plots)
%HIST2D 2d smoothed histogram calculations
% [z,xout,yout] = hist2d(xy,[xmin xmax ymin ymax],nbin)

% todo: how to set the default smoothing parameter?
% how to set the default nbin?

% Marko Laine <>
% $Revision: 1.6 $  $Date: 2013/07/12 07:34:03 $

if nargin<2 | isempty(mxmy)
  mxmy = [min(x(:,1)),max(x(:,1)),min(x(:,2)),max(x(:,2))];
end

if nargin<3|isempty(nbin)
  n = 10; % default number of intervals
else
  n = nbin;
end
if nargin<4|isempty(lam)
  lam = 25;
end
if nargin<5
  if nargout<1
    plots=1;
  else
    plots=0;
  end
end
xi = linspace(mxmy(1),mxmy(2),n+1);
yi = linspace(mxmy(3),mxmy(4),n+1);
xx = xi(1:end-1)+diff(xi)./2;
yy = yi(1:end-1)+diff(yi)./2;

zz = [];
for i=1:(n-1)
  z = hist(x(find( x(:,1) >= xi(i) & x(:,1) < xi(i+1) ),2),n);
  zz = [zz,z'];
end
z = hist(x(find( x(:,1) >= xi(n) & x(:,1) <= xi(n+1) ),2),n);
zz = [zz,z'];

if lam==0
  C = zz;
else
  C = smooth2d(zz,lam);
end
if nargout > 0
  y = C;
end

% some debugging, if plots > 1
if plots>1
  figure(1);
  xyplot(x);
  for i=1:(n+1)
    hline(yi(i));
    hline([],xi(i));
  end
end
if plots
  % figure(2); mesh(xx,yy,C);
  if plots>1, figure(2); end
  contour(xx,yy,C);
  if plots>1
    figure(3); 
    contour(xx,yy,zz);
  end
end
```
