## Supplementary material for "Quantifying non-communicable diseases’ burden in Egypt using State-Space model": S1 File: histp.m

```
function h=histp(y,nbin,dens,smo)
%HISTP   Histogram scaled as relative frequencies (probabilities)
%
% HISTP(Y) histogram for vector Y.
% HISTP(Y,NBIN) histogram with NBIN bins.
% HISTP(Y,DENS) or HISTP(Y,NBIN,DENS), where DENS is 'normal',
%  'lognor', or 'kernel' adds estimated density on top of the histogram.

% Marko Laine <>
% $Revision: 1.7 $  $Date: 2014/02/17 19:32:19 $

if length(y) ~= prod(size(y))
  error('sorry, histp works only for vectors')
end

my = mean(y); 
sy = std(y);
n  = length(y);

if nargin>1&isstr(nbin)
  if nargin>2
    smo=dens;
  else
    smo=1;
  end
  dens=nbin;
  nbin=[];
elseif nargin<3
  dens='';
  smo=1;
elseif nargin<4
  smo=1;
end

if nargin<2|isempty(nbin)
% nbin=ceil(log2(n)+1);                        % Sturges' formula
% nbin=ceil(range(y)/(3.5*sy*n^(-1/3)));       % Scott
  nbin=ceil(range(y)/(2*iqrange(y)*n^(-1/3))); % Freedman & Diaconis

  % bar(...,'w') does not work with nbin>150
  nbin=min(150,nbin);
end

% scaled histogram
[n,x]=hist(y,nbin); 
dx = x(2)-x(1);
hh = bar(x,n./sum(n)/dx,1,'w');

% add density on top
h2 = [];
switch dens
 case 'normal'
  t = linspace(my-3.5*sy,my+3.5*sy);
  hold on; h2=plot(t,norpf(t,my,sy*sy)); hold off
 case 'lognor'
  t = linspace(x(1)-0.5*dx,x(end)+2*dx);
  hold on; h2=plot(t,lognorpf(t,mean(log(y)),var(log(y)))); hold off
 case 'kernel'
  [yk,xk]=density(y,linspace(x(1)-2*dx,x(end)+2*dx,200),smo);
  hold on; h2=plot(xk,yk); hold off
end

% return handles to the bars and to the density
if nargout>0
  h=[hh;h2];
end
```
