## Supplementary material for "Quantifying non-communicable diseases’ burden in Egypt using State-Space model": S1 File: hline.m

```
function out=hline(h,v)
%HLINE  Adds a horizontal/vertical line to current figure
%  hline(h) adds horizontal line at h
%  hline([],v) adds vertical line at v

% Marko Laine <>
% $Revision: 1.5 $  $Date: 2013/10/08 12:29:01 $

ax=get(gcf,'CurrentAxes');
yl=get(ax,'YLim');
xl=get(ax,'Xlim');

if isempty(h)
   hl=line([v v], yl);
else
   hl=line(xl, [h h]);
end
set(hl,'Color',[0 0 0]);
set(hl,'LineStyle',':');
set(ax,'YLim',yl);
set(ax,'Xlim',xl);

if nargout>0
   out=hl;
end
```
