## Supplementary material for "Quantifying non-communicable diseases’ burden in Egypt using State-Space model": S1 File: hwdiag.m

```
function [B,pB]=hwdiag(chain,ns)
%HWDIAG  Heidelberger-Welch MCMC convergence diagnostics
% plot(hwdiag(chain)) plots the Brownian bridges assosiated with
% the chain columns.
% [B,pB] = hwdiag(chain) returns the p-values of Cramer-von Mises
% statistic for stationarity of the chain components.
%
% See:
% Stephen P. Brooks and Gareth O. Roberts.
% Assessing convergence of Markov chain Monte Carlo algorithms.
% Statistics and Computing, 8:319--335, 1998.

% ns is the length of the the Brownian birdge returned

% ML, 2002
% $Revision: 1.3 $  $Date: 2003/05/07 12:22:19 $

[nsimu,npar] = size(chain);

if nargin<2
  ns=200;
end

y = cumsum(chain);
x = y(nsimu,:)./nsimu;

%% Spectral density estimate for variance
n2 = floor(nsimu/2);
S0 = spectrum0(chain(n2:nsimu,:));

s = linspace(0,1,ns);
B = zeros(ns,npar);
for i=1:ns
  n = floor(nsimu*s(i));
  if n==0
    B(i,:) = zeros(1,npar);
  else
    B(i,:) = (y(n,:)-n.*x)./sqrt(nsimu.*S0);
  end
end

%BB = cramerdf(sum(B.*B)./ns)<0.95;
pB = cramerdf(sum(B.*B)./ns);
```
