## Supplementary material for "Quantifying non-communicable diseases’ burden in Egypt using State-Space model": S1 Table

| | Cardiovascular diseases | Neoplasms | Diabetes and kidney diseases | Chronic respiratory diseases |
|---|---|---|---|---|
| $\alpha_1$ | 839.168 | 599.91 | 654.02 | 645.326 |
| $\alpha_2$ | 684.477 | 564.375 | 693.03 | 689.245 |
| $\alpha_3$ | 712.42 | 569.157 | 641.06 | 719.381 |
| $\alpha_4$ | 735.693 | 504.351 | 703.167 | 712.936 |
| $\alpha_5$ | 765.067 | 545.611 | 664.934 | 718.191 |
| $\theta_1$ | 750.155 | 526.083 | 732.688 | 676.47 |
| $\theta_2$ | 837.19 | 592.724 | 663.706 | 701.252 |
| $\theta_3$ | 806.1 | 516.283 | 665.638 | 699.815 |
| $\theta_4$ | 713.969 | 557.124 | 644.762 | 780.714 |
| $\theta_5$ | 832.533 | 585.031 | 674.688 | 669.255 |
| $\theta_6$ | 808.655 | 525.088 | 772.069 | 631.405 |
| $\theta_7$ | 894.32 | 565.859 | 625.744 | 617.261 |
| $\sigma^2_{1m}$ | 790.484 | 541.741 | 734.326 | 719.643 |
| $\sigma^2_{2m}$ | 778.561 | 506.566 | 704.383 | 601.921 |
| $\sigma^2_{3m}$ | 188.831 | 513.369 | 688.776 | 898.132 |
| $\sigma^2_s$ | 181.407 | 560.031 | 701.27 | 673.709 |
