## Supplementary material for "Quantifying non-communicable diseases’ burden in Egypt using State-Space model": S2 Table

| | **Cardiovascular diseases** | **Neoplasms** | **Diabetes and kidney diseases** | **Chronic respiratory diseases** |
|---|---|---|---|---|
| $\alpha_1$ | 0.99455 | 0.93998 | 0.98648 | 0.91291 |
| $\alpha_2$ | 0.97735 | 0.99568 | 0.93718 | 0.94398 |
| $\alpha_3$ | 0.89017 | 0.96222 | 0.97605 | 0.96428 |
| $\alpha_4$ | 0.93787 | 0.98331 | 0.98744 | 0.96017 |
| $\alpha_5$ | 0.94489 | 0.99257 | 0.97337 | 0.97381 |
| $\theta_1$ | 0.97708 | 0.92936 | 0.9654 | 0.96947 |
| $\theta_2$ | 0.32154 | 0.88951 | 0.38675 | 0.13252 |
| $\theta_3$ | 0.92948 | 0.9758 | 0.96214 | 0.92362 |
| $\theta_4$ | 0.98824 | 0.95128 | 0.98271 | 0.93279 |
| $\theta_5$ | 0.37578 | 0.32454 | 0.9982 | 0.26942 |
| $\theta_6$ | 0.87583 | 0.9664 | 0.98966 | 0.95056 |
| $\theta_7$ | 0.93375 | 0.97592 | 0.90898 | 0.92887 |
| $\sigma^2_{1m}$ | 0.96928 | 0.8278 | 0.90521 | 0.923 |
| $\sigma^2_{2m}$ | 0.99208 | 0.97726 | 0.97545 | 0.97576 |
| $\sigma^2_{3m}$ | 0.99108 | 0.90289 | 0.99545 | 0.96787 |
| $\sigma^2_s$ | 0.97364 | 0.65733 | 0.97242 | 0.92012 |
