## Supplementary material for "Quantifying non-communicable diseases’ burden in Egypt using State-Space model": S3 Table

| Inference method | Number of iterations | Number of particles | Number of seconds |
|---|---|---|---|
| **Particle Filter** | | 100 | 0.159061 |
| | | 1000 | 0.33428 |
| | | 10000 | 25.49978 |
| **Particle Independent Metropolis-Hastings** | 5000 | 100 | 7.547062 |
| | 5000 | 1000 | 18.60891 |
| | 7000 | 1000 | 58.00175 |
| | 9000 | 1000 | 78.243741 |
