## Supplementary material for "Quantifying non-communicable diseases’ burden in Egypt using State-Space model": S4 Table

| **Group of disease** | **Root mean square discrepancy** | **Number of particles** | | | |
|---|---|---|---|---|---|
| | | 100 | 500 | 1000 | 10000 |
| **Cardiovascular diseases** | RMSD1 | 1.74 | 1.36 | 1.84 | 1.65 |
| | RMSD2 | 1.3 | 1.08 | 3.32 | 1.35 |
| | RMSD3 | 2.46 | 1.08 | 2.01 | 1.72 |
| **Neoplasms** | RMSD1 | 1.19 | 2.17 | 2.13 | 1.72 |
| | RMSD2 | 1.64 | 1.94 | 4.79 | 1.45 |
| | RMSD3 | 0.79 | 1.35 | 3.57 | 1.16 |
| **Diabetes and kidney diseases** | RMSD1 | 1.44 | 1.86 | 1.57 | 5.76 |
| | RMSD2 | 1.74 | 1.13 | 1.68 | 3.06 |
| | RMSD3 | 1.54 | 1.24 | 1.37 | 2.05 |
| **Chronic respiratory diseases** | RMSD1 | 1.35 | 3.5 | 3.36 | 1.94 |
| | RMSD2 | 1.17 | 1.64 | 2.19 | 1.55 |
| | RMSD3 | 1.9 | 1.81 | 5.11 | 2.28 |
